# Mechanistic modeling to understand variability in responses to chronic Hepatitis B treatment

**DOI:** 10.1101/2024.10.11.24315300

**Authors:** Solène Granjeon-Noriot, Anne Schneider, Solène Porte, Emmanuel Peyronnet, Germán Gómez, Evgueni Jacob, Nicolas Ratto, Yishu Wang, Pietro Scalfaro, Patrice André, Riad Kahoul, Claudio Monteiro, Lara Bruezière

## Abstract

Chronic hepatitis B virus (HBV) remains the most common serious liver infection globally, accounting for an estimated 820,000 deaths each year. Patient responses to treatment vary widely, due to complex interplay between viral and immune system dynamics. As yet, there is no reliable way to predict response; this is one reason cure rates remain disappointingly low (*<*10%).

We developed a mechanistic model to simulate serum viral markers evolution during two HBV treatment mainstays - the nucleoside analog entecavir (ETV) and pegylated interferon alfa (IFN) - for a variety of patients, and identify and quantify the key processes driving variability in patient responses. Based on a detailed literature review, this model integrates key processes in chronic HBV pathophysiology and drug pharmacokinetics/dynamics and was calibrated on published data only.

Post simulation regression and classification analyses, including a global sensitivity analysis and a random forest, highlighted the importance of HBV replication cycle processes in explaining pre-treatment inter-patient variability in serum viral markers. Post-treatment with entecavir, most of the response variability could be attributed to interactions between the viral replication cycle and immune system processes. Response variability after IFN treatment, however, was more directly related to the drug mechanism of action, which includes direct antiviral effects and immune system modulation. Quantifying these measures may help to inform new drug development with identification of more direct tailored and effective HBV therapy.

**Graphical Abstract:** 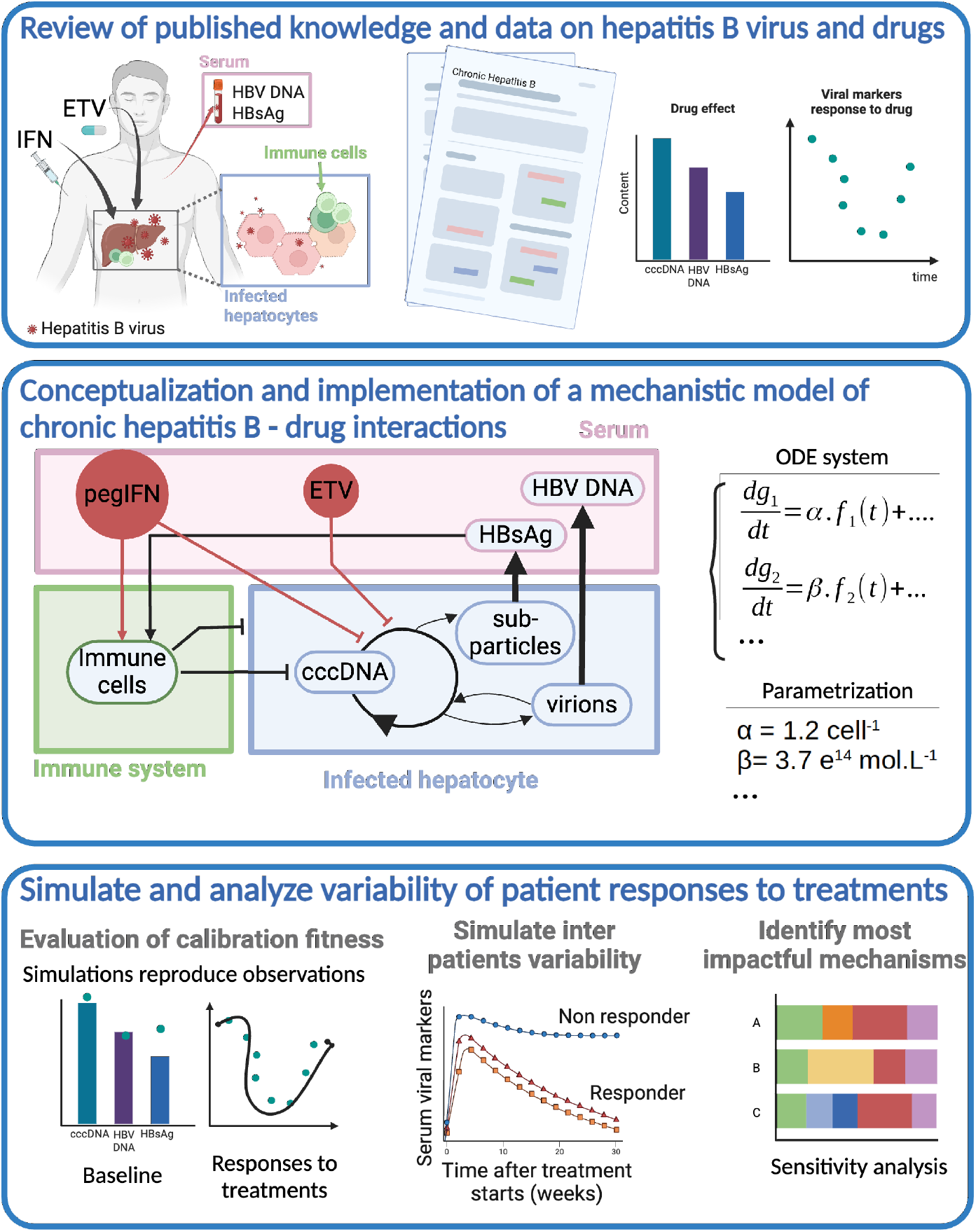

**Highlights:**

- A mechanistic model of chronic hepatitis B disease, accounting for intra-hepatocyte virus replication and an implemented immune response to entecavir and Peginterferon alfa-2a, allows for reproducing the observed variability between patients in terms of measured serum viral markers in response to treatments.
- Statistical analysis of simulated virtual populations helps investigate the mechanisms involved in observed variability between patients for both baseline and responses to treatments.
- Such a mechanistic model offers, via a QSP platform, new perspectives on the exploration of hepatitis B physiopathology, including treatment combinations or hepatitis D co-infection.

## 1. Introduction

Chronic hepatitis B infection (CHB) is a global health concern, affecting over 350 million people worldwide and accounting for a significant proportion of liver cancer cases across the globe. It led to an estimated 1.1 million annual deaths in 2022 (World Health Organization ^1^, Venook et al. ^2^). The hallmark of CHB is delineated by the enduring presence of serum hepatitis B virus (HBV) DNA and surface antigen (HBsAg) for at least six months subsequent to the initial infection (Nicolini et al. ^3^). CHB manifests as a multifaceted and heterogeneous disorder, characterized by a high degree of HBV genotype diversity, broad spectrum of symptom severity and disparate levels of resistance to treatment modalities (Datta et al. ^4^, Kim et al. ^5^). The clinical surveillance of CHB-afflicted patients predominantly entails the assessment of liver fibrosis and inflammation via levels of serum alanine aminotransferase (ALT) and aspartate aminotransferase (AST), markers of the hepatic function, and serum viral markers (SVM). Levels of HBV DNA, HBV RNA, HBeAg and HBsAg are particularly important in determining disease severity before, during and after treatment (Andersson and Chung ^6^, Berg et al. ^7^). The role of HBsAg in disease monitoring has been highlighted throughout the past decade as it is assumed to be a good indicator of the level of hepatic viral covalently closed circular DNA (cccDNA) leading to persistence of the infection (Sonneveld et al. ^8^).

Current therapies involve long term administration of nucleotides or nucleosides analogs (NUC), such as entecavir (ETV) or lamivudine as monotherapies or in combination with immunomodulators (interferons) for a defined duration. Despite high impact on serum virion levels (85-90% of patients reach undetectable HBV DNA levels after a 48-week interferon or entecavir monotherapy) (Yu et al. ^9^, Lai et al. ^10^, Chang et al. ^11^), serum HBsAg clearance is achieved in less than 10% of treated patients (Chan et al. ^12^, Zoulim et al. ^13^, Tangkijvanich et al. ^14^, Lampertico et al. ^15^, Tang et al. ^16^, Fonseca et al. ^17^) denoting the persistence of the infection. There is an urgent need for new therapeutic strategies to increase cure rates. One important strategy is to better understand factors influencing patients’ response to current treatments. Over the past decade, numerous studies have been initiated with the aim of deciphering the multilayered mechanisms driving treatment response. In vitro and animal model experiments along with clinical studies, have highlighted that CHB pathophysiology and its interactions with administered drugs rely on multiscale processes, involving several organs and intricate regulatory interplay between the multiple biological markers. Viral replication processes and mechanisms of absorption and action of drugs occur at different sites and on different time scales (Cangelosi et al. ^18^). Immune response to HBV infection shows a trade-off between stimulation and inhibition of the cytotoxic T lymphocytes (CTL) and natural killer cells (NKC) (Schuch et al. ^19^). Treatment efficacy depends on the stage of disease, and on levels of both the HBV surface antigen (HBsAg) and envelope antigen (HBeAg), found in liver and serum (Lee et al. ^20^). The dynamic interplay between these antigens and viral DNA during therapy is still poorly understood. Thus, despite insights and data provided by these studies, no reliable marker predicting patient treatments response has been identified (Liu et al. ^21^). Correlations have been established between SVM baseline levels and treatment responses, as determined by HBeAg seroconversion or HBsAg clearance (Liem et al. ^22^, Luo et al. ^23^, Ren et al. ^24^). These results were specific to trial conditions such as patients’ HBeAg status, administered drug and dose and cannot be applied to new potential therapies or combinations. Mechanistic modeling’s value in studying inter-patient variability has already been demonstrated in cardiology and oncology settings (Gadkar et al. ^25^, Kumar et al. ^26^). This kind of model is well-suited to help decipher and quantify the processes underlying complex diseases such as HBV. It captures available knowledge and data, from many sources, to build a comprehensive, multi-variables drug-disease model that can be calibrated and evaluated for goodness of fit. Models can be designed to capture HBV pathophysiology and the interactions between HBV replication cycle, immune response and treatment. Mechanistic models of intracellular hepatic and SVM have helped unravel mechanisms behind the immune system response (AsínPrieto et al. ^27^, Murray et al. ^28^, Ciupe et al. ^29^). They have uncovered the dynamics of intra-hepatic cccDNA in association with mitosis processes or liver spatial aspects (Means et al. ^30^, Murray and Goyal ^31^, Cangelosi et al. ^18^), and been used to estimate virus-related parameters such as viral particle half-lives (Nowak et al. ^32^). Models accounting for serum HBV DNA levels, and in some cases number of infected hepatocytes number, were also developed to study NUC and/or therapy effects (Colombatto et al. ^33^, Min et al. ^34^, Wolters et al. ^35^, Dahari et al. ^36^, Perelson and Ribeiro ^37^). Other models have enabled hypothesis testing around novel drug candidate mechanisms of action (Ciupe ^38^). However, none of these models individually capture all SVMs typically monitored in clinical studies, nor do they fully explain inter-patient variability in CHB treatment response.

We developed a comprehensive mechanistic model of CHB pathophysiology and standard of care treatments including Peginterferon-alfa-2a (IFN) and entecavir (ETV), hereafter named MOCHI-B (Model of Chronic Hepatitis B Infection). This model was used to determine processes involved in drug efficacy and to characterize interpatient variability in treatment response. Based on a system of ordinary differential equations (ODEs) MOCHI-B computes the dynamics of SVM (including HBV DNA, HBsAg, HBeAg and HBV RNA) in response to CHB treatments. The model accounts for processes related to the viral replication cycle, host immune system (IS), and pharmacokinetics and pharmacodynamics of ETV and IFN monotherapy treatments. The model was calibrated using published experimental and clinical data. We assessed its ability to reproduce observed SVM levels before and during treatment. The relative contributions of pathophysiological and drug related processes to variability in treatment response were analyzed using a global sensitivity analysis (SA) and a random forest (RF) analysis performed on MOCHI-B parameters. Finally, we studied how biological markers including immune cells, cytokines and intrahepatic viral markers varied for different levels of treatment response.

## 2. Materials and methods

In order to explore the processes involved in CHB inter-patient variability both in terms of pathophysiology and response to treatments, we developed a mechanistic model, MOCHI-B, which includes a model of CHB pathophysiology coupled with two standard of care (SOC) treatments. We present in this article the final version of MOCHI-B, resulting from an iterative process of literature review, modeling hypotheses formulation, implementation, testing, and model development and simplification. MOCHI-B is a knowledge-based mathematical model, built through a set of hypotheses following an extensive literature review allowing to identify main modeled biological entities and processes. The model is applied to sets of virtual populations representing variability in the parameters of MOCHI-B and is calibrated to reproduce observed variability in drug pharmacokinetics and in SVM dynamics under ETV and IFN between patients. Behaviors of the model in terms of inter-patients variability is then explored through sensitivity analysis and random forest.

### 2.1. Model design

#### 2.1.1. Model scope

MOCHI-B was designed to simulate chronic status and responses to ETV and IFN monotherapy treatments of hepatitis B infected virtual patients (VP) with low liver fibrosis levels. Treatment safety, including serum alanine aminotransferase flares, is out of scope of this work.

#### 2.1.2. Model description

MOCHI-B simulates SVMs time-course dynamics (including HBV DNA, HBsAg, HBV RNA and HBeAg) depending on patient’s characteristics and drug regimen (dose, frequency and mode of administration). It also includes intra-hepatic viral replication, host’s immune response and pharmacokinetics (PK) and mechanisms of action of ETV and IFN treatments, as illustrated in Figure 1. Biological entities, including organs, interstitial and intracellular space, where the biological processes occur are modeled as compartments. Comprehensive descriptions of biological phenomena, as reported in the literature, along with the main hypotheses and modeling choices used to build each part of the model, are provided in the Supplementary Data (Appendix A).

**Figure 1.**
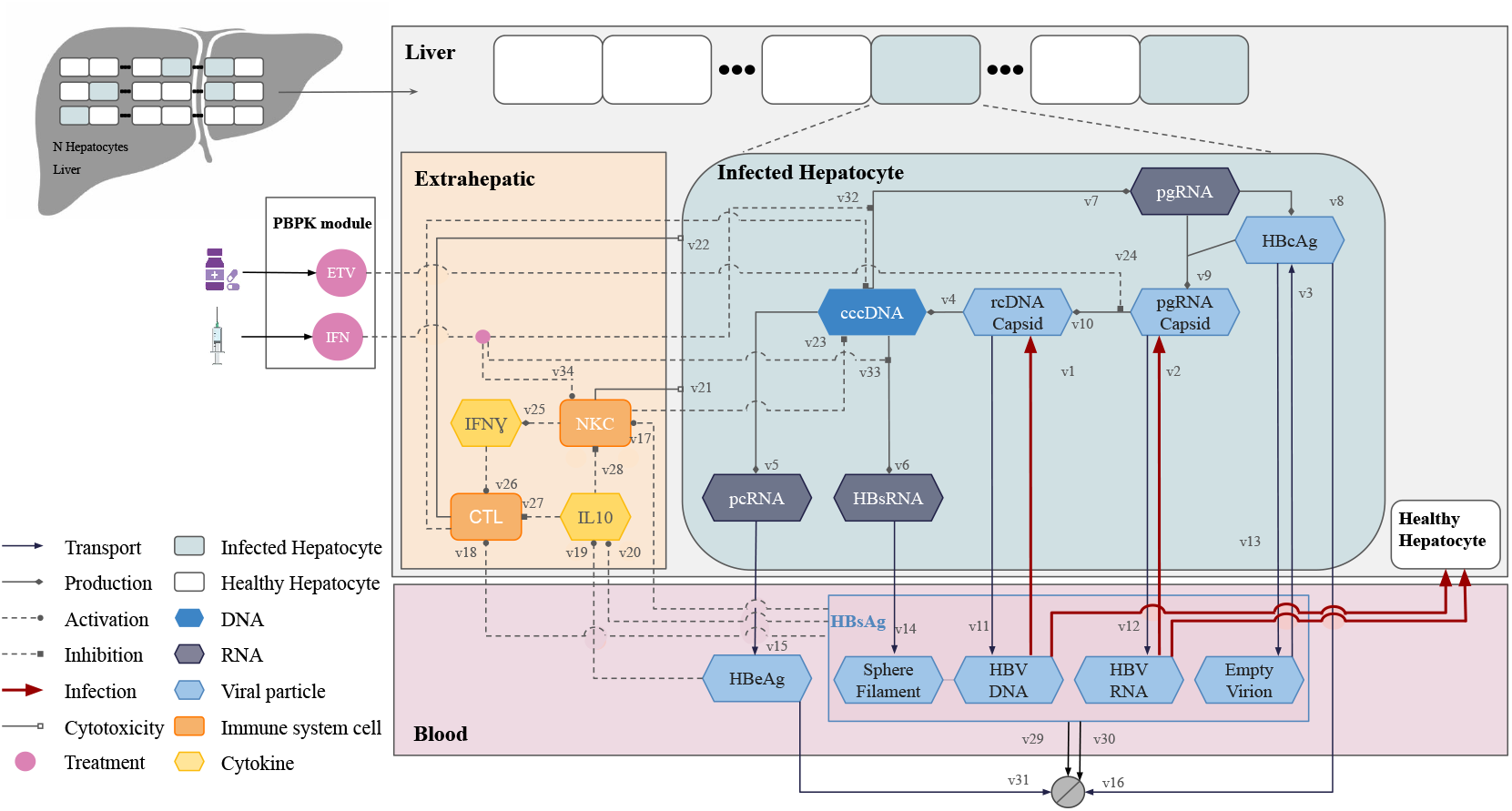
Graphical overview of MOCHI-B, a multi-scale compartmentalized model of chronic hepatitis B disease and treatments with intra-hepatic viral replication (blue box representing infected hepatocytes), host immune response against hepatitis B virus (orange box representing the extra-hepatocyte liver environment), physiologically-based-pharmacokinetic (PBPK) of entecavir (ETV) and pegylated interferon-*α* (IFN) (light gray box representing the PBPK models) and mechanisms of action of both drugs. Abbreviations are as follows: covalently closed circular DNA, cccDNA; cytotoxic T lymphocyte, CTL; hepatitis B envelope antigen, HBeAg; hepatitis B core antigen, HBcAg; hepatitis B surface antigens, HBsAg; hepatitis B surface antigen related RNAs, HBsRNA; interleukin 10, IL10; natural killer cell, NKC; pre-core RNA, pcRNA; pre-genomic RNA, pgRNA; relaxed circular DNA, rcDNA.

HBV replication cycle processes described in the literature (Yuen et al. ^39^) and implemented in the model are described hereafter and illustrated in Figure 1. Infectious virions containing HBV DNA or RNA and empty virions enter healthy and infected hepatocytes [see in figure 1: v1, v2, v3]. Capsids containing relaxed circular DNA (rcDNA) deliver rcDNA, which is then repaired by host cell mechanisms to form covalently closed circular DNA (cccDNA) [v4]. cccDNA serves as template for transcription of viral RNAs including precore RNA (pcRNA) [v5], mRNA encoding for surface proteins (hereafter called HBsRNA) [v6] and pregenomic RNA (pgRNA) [v7]. pgRNA is translated into HBV core antigen (HBcAg) [v8] and assembled into pgRNA capsid [v9]. The encapsulated pgRNA is reverse-transcribed into rcDNA [v10] which either re-enters the hepatocyte nucleus to form cccDNA, thereby amplifying the intrahepatic cccDNA pool, or is secreted in the serum as HBV DNA virions [v11]. pcRNA directs synthesis of the precore protein, a precursor of HBe antigens (HBeAg), which are then secreted into the blood [v15]. HBsRNA are translated into various proteins that are either assembled as subviral particles (sphere and filaments) or used to encapsulate viral capsids. pgRNA capsids, HBcAg, HBsRNA are secreted in the blood as HBV RNA [v12], empty virions [v13] and spheres and filaments [v14], respectively. Serum HBV DNA, HBV RNA and empty virions together with sphere and filaments constitute serum HBsAg. Naked capsids [v16] derived from intrahepatic HBcAg are also secreted. The model also provides dynamics of healthy and infected hepatocyte populations following HBV infection.

Serum viral antigens (HBsAg and HBeAg) interact with the immune system (IS) which is modeled in the liver extra-hepatocytic space. MOCHIB simulates the host’s immune reaction to viral infection and accounts for the following immunological processes: HBsAg triggers an immune response by activating natural killer cells (NKC) [v17] (Waggoner et al. ^40^, Björkström et al. ^41^) and cytotoxic T lymphocytes (CTL) [v18] (Chisari and Ferrari ^42^, Chisari et al. ^43^, Maini et al. ^44^). HBeAg and HBsAg also impair antiviral control via the activation of interleukin-10 (IL-10) production [v19, v20] (Hyodo et al. ^45^, Bertoletti and Gehring ^46^, Inoue and Tanaka ^47^). Immune cells (NKC and CTL) induce apoptosis of infected hepatocytes [v21, v22] and produce cytokines (not modeled explicitly) that lead to cccDNA degradation [v23]. Interferongamma (IFN-*γ*) production by NKC [v25] enhances CTL activation [v26]. Both CTL and NKC are inhibited by IL-10 [v27, v28]. Phagocytosis [v29] enhanced by HBsAg and IFN-*γ* also contributes to the clearance of serum viral particles [v30, v31].

IFN and ETV, respectively administered subcutaneously and orally, are distributed in major organs and eliminated through both renal and nonrenal clearance mechanisms using a classical physiologically based pharmacokinetic (PBPK) model (not detailed here), similar to the one published by Offman and Edginton ^48^. Drugs contained in the liver act through different pathways: IFN inhibits intrahepatic cccDNA transcription into pgRNA and HBsRNA [v32, v33] and activates host immunity (NKC) in the extra-hepatocytic space [v34] (Brunetto and Bonino ^49^, Shuldiner et al. ^50^). ETV interferes with reverse transcription of pgRNA into rcDNA inside capsids by HBV polymerase inhibition [v24] (Langley et al. ^51^).

#### 2.1.3. Model implementation

MOCHI-B was defined as an ODE system with each variable’s dynamic resulting from synthesis, degradation, activation and inhibition rates following established biological and physical laws such as mass-action (Voit et al. ^52^) and Michaelis-Menten (Cornish-Bowden ^53^) laws. Parameters used in these functions were estimated as detailed in the calibration section. The complete set of equations is provided in Tables D.2 and D.3. Definitions and units of parameters and variables are detailed in Tables D.4 and D.5 respectively. MOCHI-B was implemented in Haskell language (Marlow ^54^ and solved within the Nova In Silico platform using sundial solver following the multi-step ODE methods (Hindmarsh et al. ^55^, Städter et al. ^56^).

### 2.2. Model calibration and evaluation

The MOCHI-B model parameter values and ranges were calibrated to ensure that the simulations accurately reproduced average SVM baseline levels and their response to ETV and IFN monotherapy treatments, as well as the variability reported in the literature. Model simulations start with the HBV infection of healthy individuals progressing to a chronic state where serum and intrahepatic viral markers, immune status and populations of infected and healthy cells stabilize to CHB values after approximately 300-500 days. This CHB state is used as the baseline from which treatment simulations are launched. A VP is defined here as a set of parameters that reproduce a realistic and plausible behavior identified in real world data and a virtual cohort is a collection of VPs aiming to reproduce the large variability of experiment outcomes. A virtual population (VPop) is derived from the virtual cohort to reflect the different probability of occurrence of VPs in the observed clinical experiment and account for the different statistical features and experimental measurement (Gadkar et al. ^57^). Specifically, two distinct sets of parameter values derived from two reference patients were calibrated standing respectively for a HBeAg-positive (HBeAg+) patient and a HBeAg-negative (HBeAg-) patient, matching population-mean SVM baselines and response to treatments as reported in published data from Yu et al. ^9^. From these two parameter value sets, ranges were defined for parameters assumed to vary between patients in order to cover inter-patient variability reported in the literature (Cooksley et al. ^58^, Xie et al. ^59^, Lau et al. ^60^, Fried et al. ^61^, Marcellin et al. ^62^, Rijckborst et al. ^63^).

#### 2.2.1. Calibration of the two reference patients

Depending on available published knowledge and data, parameter values were either (i) extracted from scientific articles, (ii) computed from published information and model equations, or (iii) calibrated such that model outputs reproduced observed data.

1. Parameter values extracted from the literature included compartments and PBPK parameters. For example, multiple PK-related parameters were extracted from published models (Gill et al. ^64^, Li and Shah ^65^) and mass of organs and tissues as a function of age were extracted from ICRP ^66^.
2. Parameters related to biological entities homeostasis were derived from model equations to ensure homeostasis in a healthy patient. For instance the basal production rate parameter for IL-10 was computed from the IL-10 degradation rate parameter (*kdeg*) (value suggested by Huhn et al. ^67^) and IL-10 liver basal concentration in healthy adults (*Cbasal*) (Saxena et al. ^68^), assuming a constant basal production and first order degradation as *kdeg × Cbasal*. Also, the number of hepatocytes was calculated based on liver weight and hepatocellularity estimated at 99e6 cells/g in Gazzard et al. ^69^.
3. Remaining parameters for which no direct quantitative information was available were calibrated manually or using an optimization algorithm. Calibration was carried out in several steps, starting with the calibration of each model part separately, followed by the calibration of the integrated model. The calibration procedure consists in minimizing a cost function. The cost function is defined as the weighted sum of the distance between simulated and observed data (see Appendix B for details on published data used for model calibration). The weights account for the userdefined relative importance of each data-point. The cost function minimization is carried out using the CMA-ES algorithm, as described in Palgen et al. ^70^.

The HBV and IS model parameters were calibrated using data from in vitro experiments, in vivo animal models, and clinical studies, with the aim of simulating SVM values that stabilize at reference levels reported in the literature at the start of our simulation. Published data on IFN and ETV pharmacokinetics for humans and animals (serum and liver drug contents following drug administration) were used to calibrate treatments’ pharmacokinetics. Parameters related to the pathophysiology and drug efficacy were then recalibrated so that the model reproduced all observed behaviors, including stabilization to a chronic state and response to ETV and IFN monotherapy treatments.

#### 2.2.2. Reproduction of inter-patient variability

From the two reference patients, a set of parameter ranges were inferred to generate both HBeAg+ and HBeAg-VPs, constituting together a virtual cohort that reproduced observed inter-patient variability while limiting the number of unrealistic simulations: ranges were defined according to literature data when available, or with a range with high probability of including any physiologically possible value (from 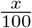 to *x×*100, where *x* is the calibrated value from one of the reference patients). The obtained virtual cohort contained various representative profiles of VPs with a large variety of behaviors and characteristics. The range of parameter values obtained are detailed in Tables D.6 and D.7.

#### 2.2.3. Evaluation of the goodness of fit

The performance of MOCHI-B in simulating observed behaviors and inter-patient variability in terms of drug pharmacokinetics, SVM and intrahepatic viral marker levels before and after treatments was assessed by comparing observations from clinical and in vitro studies with simulations of virtual populations (VPop). Data processing was performed using R Statistical Software (v.4.2.0; R Core Team (2021)) and graphs were produced with the ggplot2 R package (Wilkinson ^71^).

##### CHB disease intensity

To explore MOCHI-B performance to simulate chronic VP and a realistic inter-patient variability for SVM baseline levels, 40,000 VPs were generated using calibrated parameter values and ranges gathered in Tables D.6 and D.7. Simulations were run for 1000 days without treatment starting with a HBV inoculation (1 thousand virions equivalent) at t0. VP presenting low SVM levels (serum HBsAg below 2 log10 IU/mL and HBV DNA and RNA below 2 log10 copies (cp)/mL) were discarded, to select only those with chronic infection. The remaining 37,908 VPs constituted the virtual cohort. Simulated SVM, intrahepatic and host immune related variables ranges at t = 1000 days were compared with data reported in the literature.

##### Drugs pharmacokinetics

VPops used to assess simulations of ETV and IFN pharmacokinetics were designed to reproduce clinical studies published by Yan et al. ^72^ for ETV treatment and Costa et al. ^73^ for IFN treatment. 24 and 100 VPs were generated with summarized patient characteristics (age, body weight, height and gender) matching those of the published studies. The ETV renal clearance parameter and parameters related to IFN pharmacokinetics varied within their calibrated range for ETV and IFN treatments respectively. Simulations were run on the generated VPops considering daily administrations of 0.5 mg p.o. under fasting conditions for 14 days for the ETV VPop and a single injection of 180*µ*g s.c. and a 15-days follow-up for the IFN VPop. Simulated maximum drug concentration in the serum (Cmax), time for which Cmax is reached (tmax) and the area under the curve (AUC, i.e. integral of the drug in serum as a function of time) values for day 14 and from 0-36h for ETV and IFN treatment scenarios respectively were computed from simulated time-course dynamics of drug serum contents and compared to published data.

##### Responses to treatments

MOCHI-B performance to simulate the variety of observed SVM, intrahepatic and immune marker responses to ETV and IFN monotherapies was assessed by running simulations on the virtual cohort of 37,908 chronic VPs. Simulated variability of physiological markers at EOT after ETV (0.5 mg p.o. QD for 48 weeks) and IFN (180 *µ*g QW s.c. for 48 weeks) monotherapies were compared with observations from published clinical trials.

In addition, we evaluated the ability of the parametrized MOCHI-B to reproduce a specific clinical study. Two subsets of 122 and 56 VPs were selected from the virtual cohort of chronic VPs to reproduce results presented by Yu et al. ^9^ for both ETV and IFN monotherapies, respectively. VPs were sampled following the method described by Allen et al. ^74^, using population-level data extracted from Yu et al. ^9^ on SVM levels at baseline and in response to treatments over time, as well as undetectable HBV DNA and RNA rates at EOT as constraints to define VPs probabilities of inclusion. Simulations were run for ETV and IFN monotherapies and compared to observations reported for 5 time points (0, 4, 12, 24 and 48 weeks). Goodness of fit was assessed by computing coverage and precision percentages between simulated and observed data considering a range of two SD centered around the mean values. Coverage is defined as the percentage of observed intervals included within simulated intervals across time, and assesses the model ability to cover the range of observed data. Precision is defined as the percentage of simulated intervals included within observed intervals across time and assesses the model ability to provide results with a reasonable variability. Statistical analyses were performed with Nova In Silico’s R-based package JinkoStats.

## 2.3. Exploration of model behavior

### 2.3.1. Exploration of inter-patient variability in SVM

A global sensitivity analysis (SA) based on the computation of Sobol/Saltelli indices was performed to quantify the relative contributions of the biological processes in the observed inter-patient variability in SVM before and after ETV or IFN treatments. SA process followed the following steps to guarantee correct results interpretation (Saltelli et al. ^75^, Rodriguez-Fernandez et al. ^76^, Friedrich ^77^). (i) Selection of outputs of interest. SVM values at beginning of treatment (BOT) and SVM absolute changes at end of treatment (EOT) after ETV and IFN monotherapies were selected as outputs of interest for the global SA. (ii) Selection of SA input factors. 58 parameters of MOCHI-B (see Table D.5 in Supp. 4 for the whole list of model parameters) were selected as SA input factors, including patient characteristics (n=3), HBV parameters (n=23), IS parameters (n=20) and treatments related parameters (n=3 for ETV and n=9 for IFN). (iii) Fractional factorial design (FFD). To limit the number of simulations, a FFD of size 40,000 was set using a modified Federov algorithm implemented in the AlgDesign R package (see Supp. 3 for details). It was defined for 45, 48 and 54 parameters for SVM at BOT, EOT after ETV and IFN monotherapies respectively, varying on 5 levels within their calibrated ranges, except for the binomial gender parameter. The FFD allows estimating main effects and two-factor interactions assuming that higher-order interaction contributions in the output variances are negligible. Non-chronic patients were removed from the FFD before launching the SA leaving 34,782 and 34,788 simulated VPs for ETV and IFN treatments respectively. (iv) Outputs of interest uncertainty quantification. Uncertainty in the model output value was quantified through the quantification of the variability of simulated SVMs, computing mean, variance, 10th and 90th percentiles for each output of interest (Eck et al. ^78^). (v) Variance-based global sensitivity analysis. The global SA was carried out using the R package multisensi v.1.2.1 (R version 3.6.3) (Bidot et al. ^79^, R package - Cran ^80^) with inputs varying all at the time within their calibrated ranges (refer to section (iii)). Main sensitivity (or first-order) index (MSi), interaction sensitivity index (ISi) and total interaction sensitivity index (TISi) were computed to quantify impact of each input on each model output of interest, as defined in Saltelli et al. ^81^, Sobol ^82^, Monod et al. ^83^, Homma and Saltelli ^84^, Puy et al. ^85^. MSi measures the direct contributions of inputs to the output variance, while ISi and TISi account for contributions of second-order interaction between inputs to the output variance. For each output, the part of the output total variance that is not explained by selected inputs MSi and TISIi is quantified in the residuals (Res).

#### 2.3.2. Exploration of cure status

Random forest (RF) classification were performed using the randomForest R package (v.4.7-1.1) to assess contributions of MOCHI-B parameters - used as inputs of the RF - on VP cure status (cured or uncured) at EOT after ETV and IFN monotherapies as explained by Zhang and Tyson ^86^. The lower limit of detection (LOD) varies in the literature depending on the methods and manufacturers. Therefore, in this study and throughout the article, for the sake of simplicity, the lower limit of detection is set to 0 (corresponding to a perfect detection method). Consequently, patient cure was defined as clearance of the virus, with serum HBV DNA and RNA below 0 log10 cp/mL and HBsAg below 0 log10 IU/mL). For each treatment, a RF algorithm composed of an ensemble of 500 decision trees, testing between 8 or 9 inputs at each node split, was trained on 75% of the virtual cohort selected randomly (Breiman ^87^). The classifier ‘cutoff’ argument of the randomForest R function was set to 0.9 to account for unbalanced categories since simulated number of cured VPs at EOT was much lower than the number of uncured patients (1,283 vs 33,505 for IFN and 1,233 vs 33,549 for ETV). RF prediction capacity to classify patients’ cure status was then tested on the remaining 25% of VPs based on the confusion matrix and the computation of classification metrics (accuracy score, sensitivity and specificity) (Parikh et al. ^88^). For each MOCHI-B parameter, its contributions in predicting cure at EOT for ETV and IFN were expressed with a “parameter importance” computed as the reduction in accuracy resulting from permuting RF inputs (Breiman ^87^). The parameters that, after being permuted, decrease model accuracy the most were deemed to be the most impactful for patients’ cure status at EOT.

#### 2.3.3. Exploration of responding patients categories

Simulated baseline levels of SVM, intrahepatic and immune markers were compared for three categories of VPs. High, medium and low responder categories were defined based on patients’ infection status at EOT and serum HBsAg and HBV DNA relative change at EOT as follows: (1) high responders for cured patients or patients with HBsAg relative decrease at EOT beyond 50% and HBV DNA relative decrease at EOT beyond 40%; (2) medium responders for patients with HBsAg and HBV DNA relative decrease at EOT respectively between 1-50% and 20-40%, (3) low responders for the remaining patients.

## 3. Results

### 3.1. Model design

Knowledge and data were extracted from 206 papers out of 858 initially reviewed papers. The final knowledge-based mathematical model, formulated as a set of ODEs, simulates SVMs of chronic hepatitis B patients in response to ETV and IFN monotherapies. The model incorporates intermediate processes related to HBV pathophysiology, the host’s immune response, and PBPK submodel. The combined MOCHI-B model consists of 16 ODEs and accounts for 20 species distributed across 5 compartments (see model schema in Figure 1, and variables, fluxes, and ODEs lists in Supp. Appendix D). The combined model includes 91 parameters listed in Tables D.2, D.5, with 21 parameters extracted from the literature, 23 related to protocol design, derived from model equations or arbitrarily set, and 47 calibrated.

### 3.2. Model calibration and evaluation

#### 3.2.1. Virtual population reproduces baseline pretreatment variability seen in clinical data

The 37,908 VPs of the virtual cohort displayed stabilized SVM levels at 142 weeks after infection (dynamics not shown). The virtual cohort covered inter-patient variability in terms of SVM, intrahepatic and immune response markers before treatment observed in clinical data. In summary, the virtual cohort structure included both HBeAg-positive (67%) and HBeAg-negative (33%) patients with serum HBV DNA levels ranging from 2.8-9.9 log10 cp/mL and serum HBsAg from 2.0-6.2 log10 IU/mL. Simulated intrahepatic viral markers including cccDNA, pgRNA, pcRNA and rcDNA covered observations reported by Dandri et al. ^89^, Lesmana et al. ^90^, Laras et al. ^91^, Volz et al. ^92^, Wang et al. ^93^ and Wang et al. ^94^. Simulated host immune markers covered a wide-range of values mirroring different levels of host immune activation with both the immunoactive cytokine interferon gamma (IFN-*γ*) ranging from 2.0 to 45.5 pg/mL, matching observations ranging from 0 to 63.5pg/mL (Falasca et al. ^95^) and the immunosuppressive cytokine IL-10 ranging from 81 to 356 pg/mL partly covering observations ranging from 20 to 240 pg/mL (Saxena et al. ^68^, Arababadi et al. ^96^).

#### 3.2.2. Drugs pharmacokinetics data consistently reproduced by model simulations

ETV PBPK drug model satisfyingly reproduced published serum ETV time-course dynamics (Yan et al. ^72^). Simulated serum ETV content sharply increased during the first hours following administration of the drug (0.5mg QD), reaching a mean Cmax of 3.6 pg/mL (CV 9.8%) between 1 and 1.5h (Figure 2 A). In comparison, Yan et al. ^72^ reported a mean Cmax of 4.23 pg/mL (CV 9%) and a tmax ranging between 0.5 and 1h. The simulated and observed mean daily AUC at day 14 were similar (11.27 versus 14.78 ng.h/mL, with CV of 9.2% and 17% respectively).

**Figure 2.**
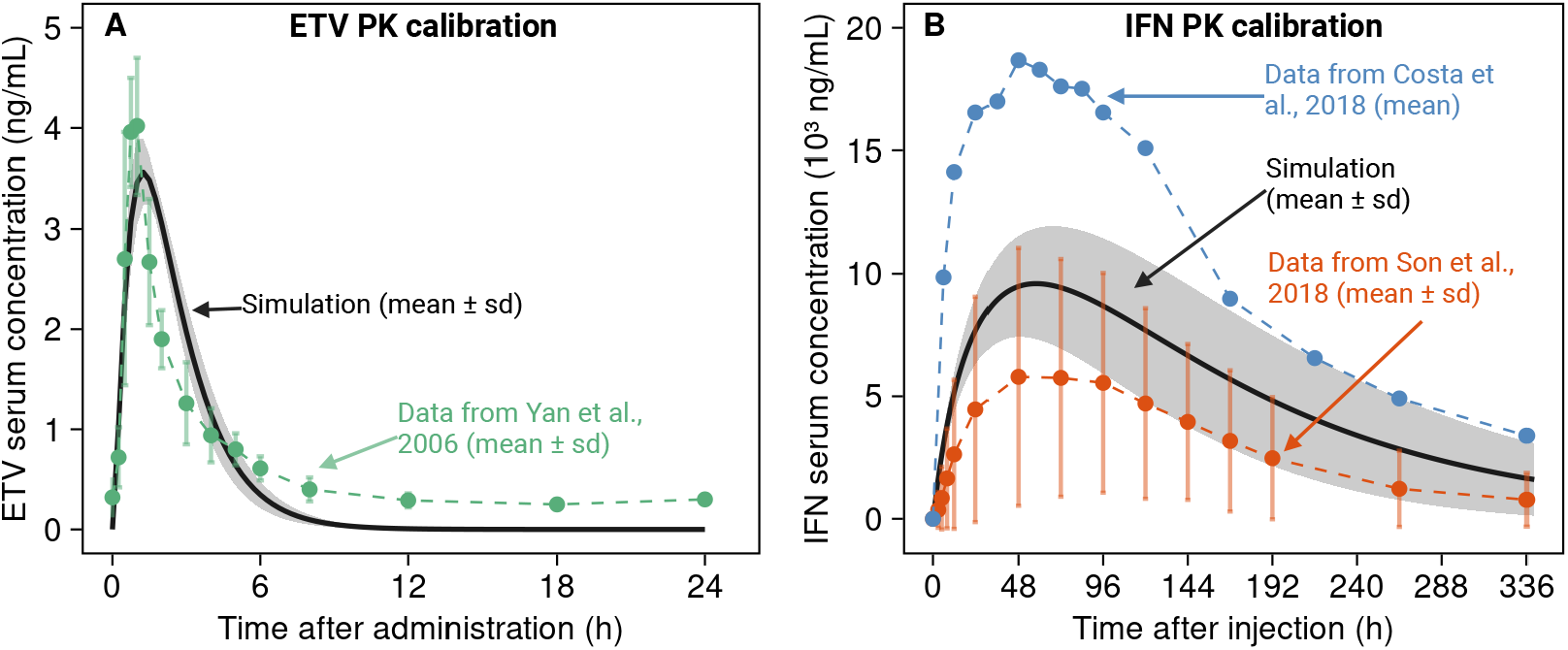
Comparison of the time course of serum drug concentrations as simulated by the model and reported in the literature for (A) entecavir (ETV) on day 14 following 14 daily administrations of the drug (0.5mg QD) and (B) peg-interferon alfa (IFN) after a 180 *µ*g single dose s.c.. Mean simulated values are represented in solid black lines (shaded areas represent*±*SD) and literature mean data are represented in green (Yan et al. ^72^), blue (Costa et al. ^73^) and orange (Son et al. ^97^) (mean*±*SD when available).

Simulations obtained with the IFN PBPK drug model were also consistent with observations from published clinical studies (Costa et al. ^73^ and Son et al. ^97^). Simulated serum IFN content sharply increased following the time of administration (180 *µ*g single dose s.c.) to reach a mean Cmax 9,811 (SD 2,364) pg/mL (range 5,590 to 18,463 pg/mL) at 60 (SD 15) h (range 40 to 110 h) after injection (Figure 2 B). In comparison, Costa et al. ^73^ and Son et al. ^97^ reported mean Cmax of 19,960 (SD 7,066) pg/ml and 7,435 (SD 5,165.7) pg/mL and mean tmax of 54 (SD 18) and 83.4 (SD 46.3) h respectively. Simulated total AUC from administration to 336h was on average equal to 1.93 (SD 0.79) *µ*g.h/mL and varied between 0.94 and 4.66 *µ*g.h/mL, while mean observed values were equal to 1,17 (SD 0.79) *µ*g.h/mL (Son et al. ^97^) and 3.39 (SD 1.56) *µ*g.h/mL (Costa et al. ^73^).

#### 3.2.3. Full range of observed treatment response captured by the virtual cohort

Simulation of ETV and IFN monotherapy treatments for the 37,908 chronic VPs in the virtual cohort generated a wide range of treatment responses, consistent with observations from the literature (Table 1). In particular, the virtual cohort covered the wide spectrum of response to both treatments, from non-responder to functional cure, demonstrating that the calibrated parameter ranges were adapted to simulate observed variability. Based on this virtual cohort, virtual populations can be extracted to reflect realistic proportions of patients and their corresponding treatment responses.

**Table 1:** Patients baseline characteristics by treatment group and HBeAg status in the Vpops designed to reproduce Yu et al. ^9^.

| Characteristics | HBeAg-positive patients |  | HBeAg-negative patients |  |
| --- | --- | --- | --- | --- |
|  | Simulated | Yu et al. <sup>9</sup> | Simulated | Yu et al. <sup>9</sup> |
| **Patients receiving ETV (n=122)** |  |  |  |  |
| Total number of patients | 74 | 72 | 48 | 50 |
| No. male/female | 47/27 | 54/18 | 17/31 | 35/15 |
| Age (yr) (mean±SD) | 51.09±14.08 | 35.4±8.9 | 51.7±13.45 | 47.8±12.9 |
| HBsAg (log10 IU/mL) (mean±SD) | 4.31±0.65 | 4.16±0.72 | 4.07±0.67 | 3.19±0.44 |
| HBV DNA (log10 cp/mL) (mean±SD) | 6.06±1.44 | 7.36±1.00 | 5.33±1.55 | 5.54±1.17 |
| HBV RNA (log10 cp/mL) (mean±SD) | 6.98±1.9 | 7.64±1.37 | 6.2±2.05 | 4.79±1.25 |
| **Patients receiving IFN (n=56)** |  |  |  |  |
| Total number of patients | 51 | 44 | 5 | 12 |
| No. male/female | 30/21 | 27/17 | 3/2 | 11/1 |
| Age (yr) (mean±SD) | 53.76±16.52 | 30.8±7.4 | 58.13±24.54 | 36.1±5.2 |
| HBsAg (log10 IU/mL) (mean±SD) | 3.86±0.72 | 3.77±0.71 | 4.29±0.97 | 3.23±0.57 |
| HBV DNA (log10 cp/mL) (mean±SD) | 5.78±1.77 | 6.97±1.03 | 4.21±1.72 | 5.27±1.46 |
| HBV RNA (log10 cp/mL) (mean±SD) | 6.07±2.13 | 6.78±1.39 | 3.76±0.99 | 4.46±1.65 |

#### 3.2.4. A specific clinical study can be reproduced by a subset of the virtual cohort

Two subpopulations were calibrated by sampling VP from the virtual cohort using the method described by Allen et al. ^74^ in order to reproduce results reported by Yu et al. ^9^ as detailed in the Methods section. Baseline characteristics and simulated SVM responses to ETV and IFN treatments of VPs in these 2 calibrated VPops were consistent with the observed data. In both Vpops, the proportions of males and HBe positive patients, mean age and baseline values of HBV DNA, HBsAg and HBV RNA were similar to patient characteristics reported in Yu et al. ^9^ (Table 1). The efficacy of ETV treatment in reducing serum HBV DNA and HBsAg was slightly underestimated between weeks 4 and 24 compared to observed data (Figure 2). In particular, the percentage of patients undetectable SVM levels were underestimated in the simulations (Figure 3 A1). However, regarding the SVM levels dynamics, the inter-patient variability was well represented and the trend in SVM decrease under ETV was accurately reflected (Figure 3 A2 - A3). Response to IFN monotherapy was also satisfyingly reproduced by MOCHI-B and the Vpop (Figure 3 B2-23), which covered the variability in SVM response observed with coverage ranging between 70.9% and 83.9%, and reproducing the observed trend with precisions ranging from 79.3 to 93.7%. Results for the additional serum viral marker HBV RNA are given in the Supplementary Figure E.6.

**Figure 3.**
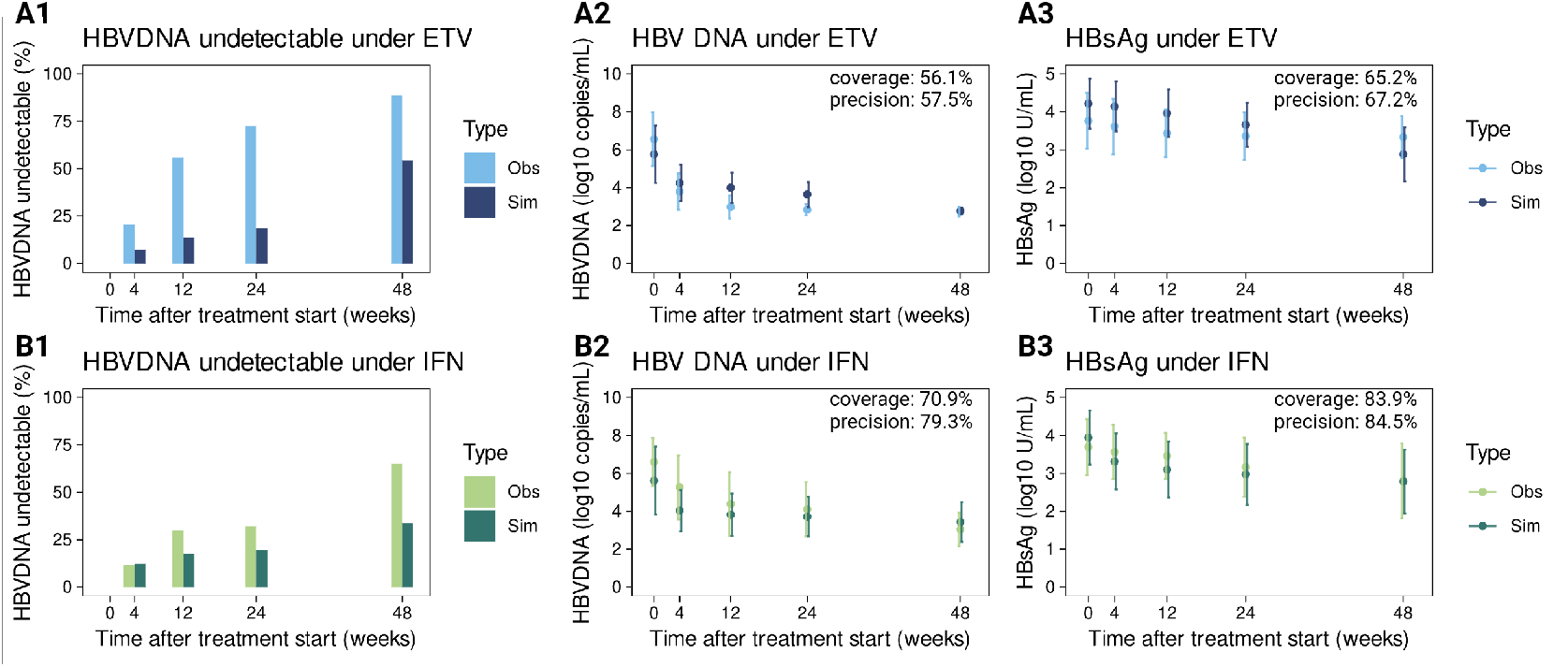
Simulated versus observed HBVDNA and HBsAg levels under ETV and IFN treatments. Simulations are obtained for virtual populations calibrated on Yu et al. ^9^ observations. In all plots, “Sim” (dark color) refers to simulated values obtained with calibrated virtual populations, while “Obs” (light color) refers to data extracted from Yu et al. ^9^. Data obtained under ETV treatment are in blue, and in green for IFN treatment. Percentage of patients with undetectable levels of HBVDNA at different time points are presented under ETV (A1) and IFN (B1) treatments. SVM dynamics (mean *±* sd) under ETV and IFN treatments are presented for HBVDNA (A2 and B2) and HBsAg (A3 and B3). Coverage and precision indicators are computed as explained in the Material and Methods section.

### 3.3. Exploration of model behavior

Contributions of MOCHI-B parameters in explaining simulated variability were assessed for each SVM (HBV DNA, HBsAg, HBeAg and HBV RNA) at BOT and EOT after ETV and IFN monotherapies using GSA (see Figure 4 for results related to HBV DNA and HBsAg, and Figure E.7 for results related to HBV RNA and HBeAg) and RF. Simulated variability at both BOT and EOT aligns with observations from Yu et al. ^9^ (Tables D.6 and D.7).

**Figure 4.**
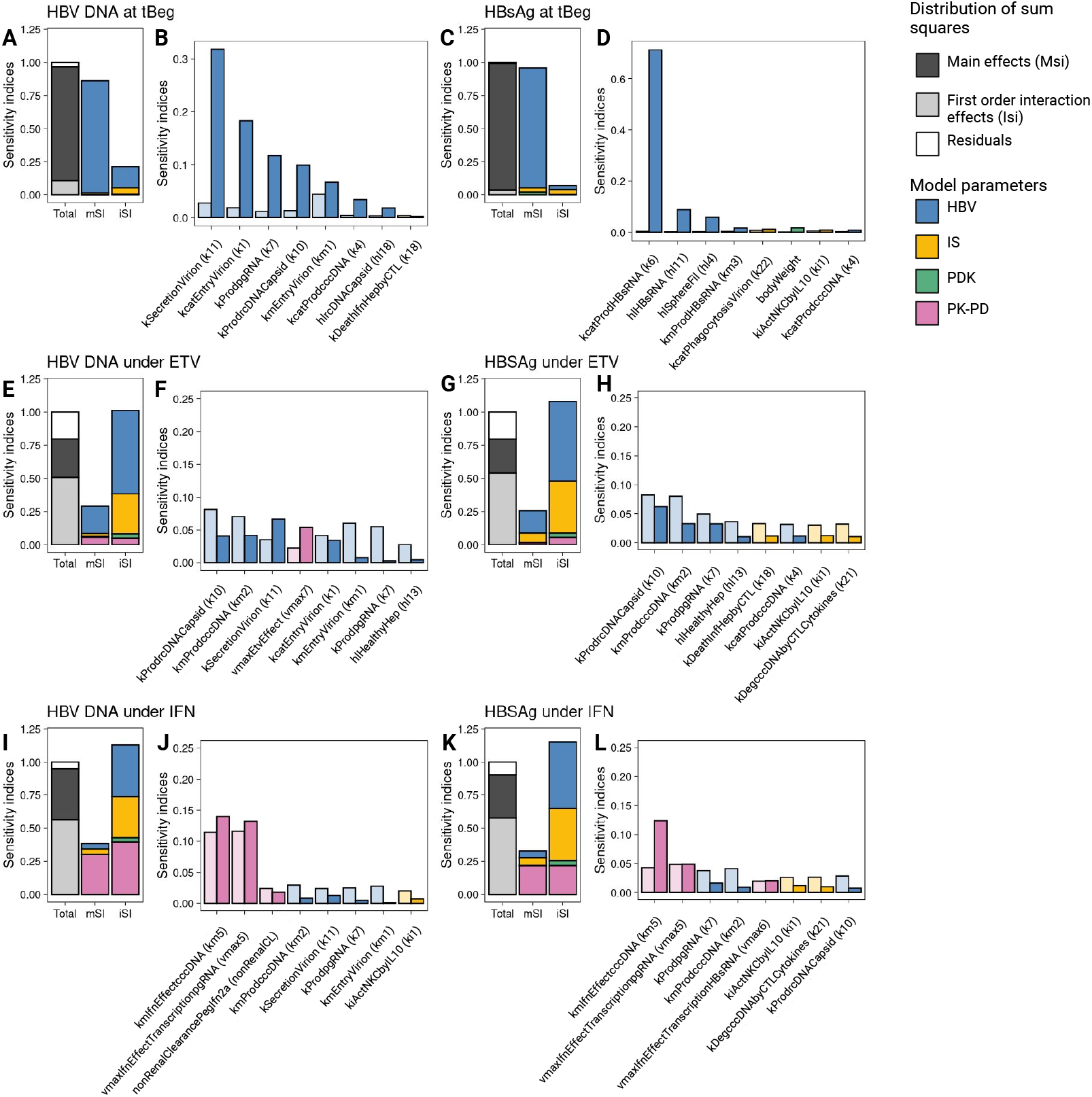
Contributions of MOCHI-B parameters in simulated SVM variability. GSA sensitivity indices for serum HBV DNA and HBsAg values at BOT and decreases between BOT (A to D) and EOT (E to L). The plots A, C, E, G, I and K show the distribution of MSi and Isi and contributions of each parameter type in these indices. HBV submodel parameters are colored in blue, immune system related in yellow, drugs PK-PD in pink, and patients’ known descriptors (PDK) such as age or body weight in green. The plots B, D, F, H, J and L show the effect of the 8 most important parameters for each outcome of interest.

#### Variability at BOT is explained by HBV replication processes

According to GSA results, MOCHI-B parameter main effects (MSi), and in particular MSi of HBV replication cycle related parameters, explained most of the simulated variability of SVMs at BOT (Figure 4 A, C and Figure E.7 A, C). MSi of 6 parameters of the intra-hepatocyte viral replication cycle-namely, kSecretionVirion (k11), kcatEntryVirion (k1), kProdpgRNA (k7), kProdrcDNACapsid (k10), kmEntryVirion (km1) and kmProdcccDNA (km2)-explained most of the variability in HBV DNA (Fig. S4A and B) and HBV RNA (Figure E.7 A, B) values at BOT. In contrast, variability in HBsAg (Figure 4 C, D) and HBeAg values at BOT (Figure E.7 C, D) were mostly explained by the MSi of the parameters directly involved in their respective syntheses, specifically kcatProdHBsRNA (k6), and kProdpcRNA (k5).

#### ETV-treatment responses tied to IS and HBV related parameters

In contrast to SVM values at BOT, variability of SVM decrease at EOT was explained in GSA results by both parameter MSi and their first order interactions (Isi) (Figure 3 E, G, I, K). Main effects of HBV related parameters (such as kProdrcDNACapsid (k10) and kmProdcc-cDNA (km2)) and Isi between parameters related to HBV replication cycle and the IS explained most of the variability under ETV monotherapy (Figure 4 E, F, G, H and Figure E.7 E, F, G, H). HBV and IS related parameters, and in particular mechanisms involved in the intrahepatic viral replication cycle, also appeared to be the most impactful on treatment response in terms of patient cure according to RF results (Figure E.8).

#### IFN-treatment response is tied to first order interactions and relies on both HBV, IS, PK and PD related processes

Regarding responses to IFN monotherapy at EOT (Figure 4 I, K), most of the variability was explained by first order interactions between parameters related to HBV replication cycle, IS or PK and PD of the drug (TIsi between 50 and 73% of the total variability). In contrast to ETV therapy, major parameters’ main effects relied on treatment related parameters (MSi of 0.29, 0.28 and 0.21 for HBV DNA, HBV RNA and HBsAg respectively), and in particular on kmIfnEffectcccDNA (km5) and vmaxIfnEffectTranscriptionpgRNA (vmax5) (Figure 4 J, L). RF results confirmed the importance of processes directly related to IFN mechanism of action and to the viral replication cycle in explaining patient cure (Figure E.8).

##### 3.3.1. Immune and intrahepatic markers predict patient response better than SVM baseline values

Patients’ characteristics associated with treatment response were further explored by classifying VPs into 3 categories: high, medium and low responders (as detailed in the Materials and Methods section) to compare their viral and immune marker baseline levels. 4,838 (2.4%) and 2,491(6.6%) patients displayed high response to ETV and IFN monotherapy treatments respectively. 17,323 (9.7%) and 6,251 (16.5%) VPs were classified as medium responders and the remaining 15,7547 (87.9%) and 29,166 (76.9%) VPs were categorized as low responders under ETV and IFN treatment respectively.

For both patients treated with ETV (results shown in Figure 5) and with IFN (results shown in Figure3), the three categories of treatment responders presented similar serum HBsAg and HBeAg baseline values. Mean and median values of serum HBV DNA and RNA baseline values were slightly lower for high responders compared to medium and low responding patients despite the high overlap between the 3 categories that shows that SVM baseline levels are poor treatment response predictors (Figure 5 C).

**Figure 5.**
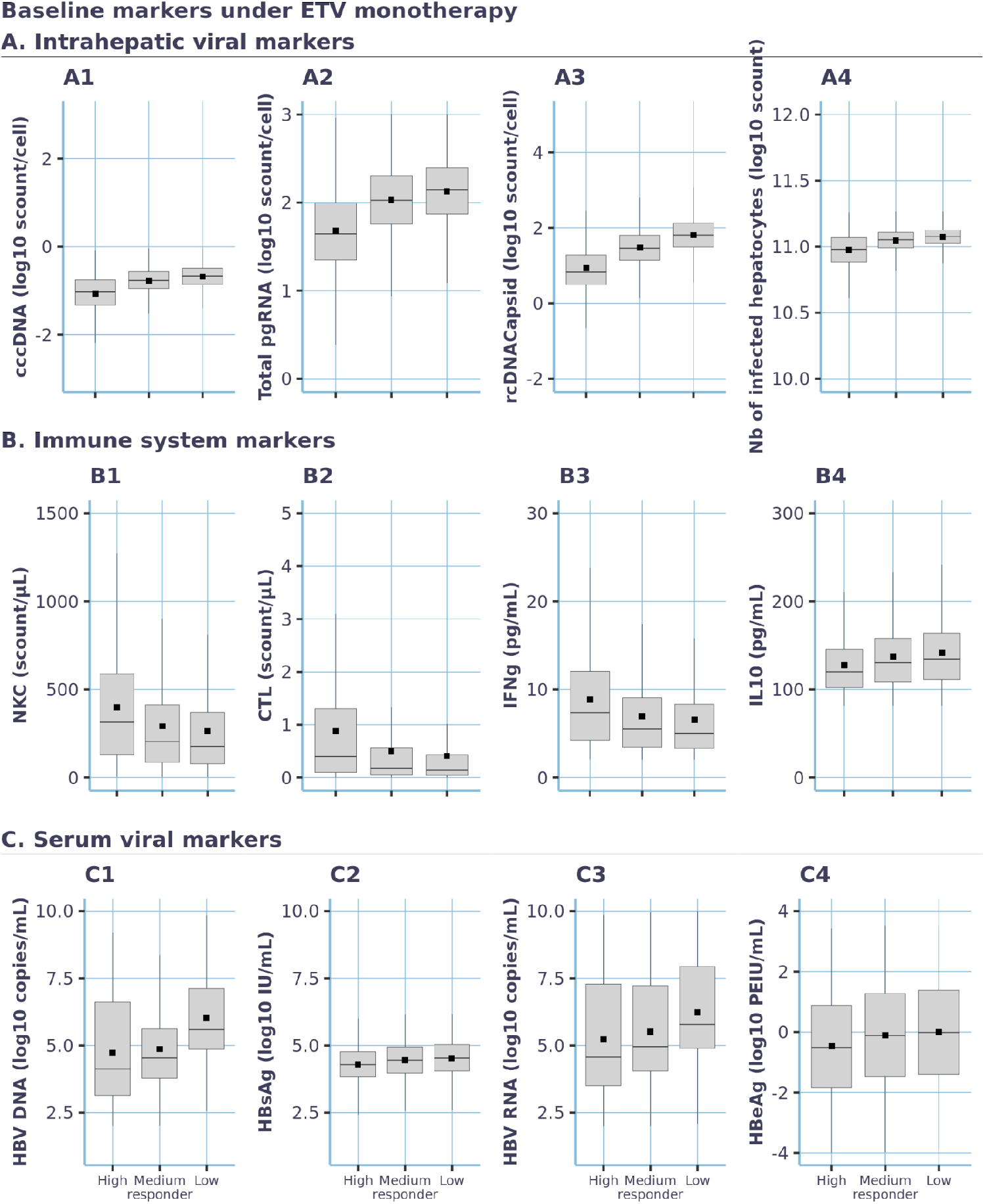
Graphical comparison of intrahepatic, immune system and serum viral markers baseline values among high, medium and low responders under ETV monotherapy treatment. The boxplots display min, 1st quartile, median, 3rd quartile and maximum values. Black squares correspond to mean values. Intrahepatic and serum viral markers are presented in log10.

Differences between responder categories were more noticeable for immune and intrahepatic viral markers levels (Figure 5 A and 5.B for ETV and Figure E.9 A and Figure E.9 B3 for IFN). Low and medium responders presented similar simulated baseline values for immune and intrahepatic viral markers. In contrast, high responders differed from the other VPs categories by lower levels of intrahepatic markers: cccDNA, pgRNA and rcDNA capsid and number of infected cells. In addition, high responder patients had higher basal immune cells (CTL and NKC) and pro-inflammatory cytokine levels (IFN-*γ*) and lower anti-inflammatory cytokine levels (IL-10) (Figure 5 B) suggesting a more efficient immune response at baseline.

Overall these exploratory analyses concur with the former SA and RF results on the importance of intrahepatic viral replication cycle and immune system status and suggest that immune and intrahepatic markers are better predictors of patients response than SVM baseline values under ETV and IFN treatments.

## 4. Discussion and conclusion

Despite the existence of several commercialized drugs, chronic HBV remains a major health issue. Extensive research has been conducted during the last decade to study host and viral markers. The levels of these markers, measured before or few weeks after treatment start, could correlate with treatment efficacy [ref], however there is no consensus on which markers are the best predictors of efficacy. We developed a multi-scale mechanistic model of chronic hepatitis B in response to two SOCs based on public biological and clinical data, in order to better understand biological and pharmacological processes leading to the observed inter-patient variability. The multi-scale model incorporates immune response and intrahepatic viral replication processes, simulates dynamics of all SVMs usually monitored by clinicians, including HBsAg and HBV DNA, and reproduces the variability of the responses to two SOCs as described in the literature. A sensibility analysis (SA) performed on main SVM values before and after ETV and IFN monotherapies highlights the importance of viral and immune-related processes in explaining patients’ response to treatments.

Assumptions about the distributions of input factors have a large impact on GSA results (Rodriguez-Fernandez et al. ^76^). The step-by-step process leading to the definition of parameter ranges prior to a SA, as well as the choice of parameters that may vary between individuals are rarely detailed in modeling papers. Here, we tried to justify with available knowledge and data all parameter values and ranges. Calibrated ranges of parameters values lead to simulations consistent with observations reported in a wide set of publications at different scales (from intrahepatic variables to SVM dynamics). Simulated variability was compared to real data prior to performing the GSA. Some biological processes are hardly measurable in vivo for humans, requiring the use of data from other species or from different biological contexts. This can be a limitation of parameter range estimations, as also observed in previous studies of HBV modeling (AsínPrieto et al. ^27^). In particular, quantitative data on intra-hepatocyte variables related to the virus replication cycle or local immune responses are, to our knowledge, inexistant in published literature for humans under treatments. Model validation might be performed if quantitative data become accessible to confirm the fitting between simulations and observations.

Model results suggest a high contribution of virus-related parameters in explaining disease intensity prior treatment and, to a lesser extent, response to ETV and IFN treatments. Wide ranges were set for intrahepatic virus-related parameters such that the virtual cohort reproduced observed inter-patient variability in SVM levels prior treatment, which concurs with the diversity of HBV (10 genotypes and over 40 variants) leading to different virological features including replication, viral mRNA and DNA expression and virion and antigen secretion capacity (Sunbul ^98^, Tong and Revill ^99^, Campos-Valdez et al. ^100^) as well as different treatment response (Baldick et al. ^101^, Wong et al. ^102^, Campos-Valdez et al. ^100^, Khatun et al. ^103^). Host immune system-related parameters had a lower contribution on disease intensity prior treatment, which can be explained by the fact that parameter ranges were selected to simulate chronic patients only, hence reducing the broad spectrum of anti-HBV immunity found in response to HBV infection (Boni et al. ^104^).

Further, both the global SA and RF results showed that parameters linked to the viral inhibition of the immune response or explaining interactions between viral and immune response had a strong impact on patient’s response to IFN and ETV treatment, which concurs with the notion that restoring a functional host’s immune response is crucial to achieve functional cure (Ezzikouri et al. ^105^, Yang et al. ^106^, Chang et al. ^107^). Besides, stronger basal antiviral immunity, defined in our model by higher levels of immune cells and lower levels of IL-10 and intrahepatic viral markers, were also associated with better treatment response in agreement with Chan et al. ^108^ where patients with lower cccDNA and total intrahepatic HBV DNA baseline level had a higher sustained virological response after treatment. In contrast, although baseline SVM levels were on average lower in high responders than in low responders, in agreement with clinical observations (Boni et al. ^104^, Zeuzem et al. ^109^), the high overlap of simulated SVM levels between high and low responders suggests that SVM are poor predictors of treatment response.

The model was built to account for Chronic HBV in the context of a phase 2 clinical trial, with durations of treatment limited to 48 weeks. Due to this context and the lack of individual patients data, predictions regarding long-term treatment efficacy carry a high degree of uncertainty. Besides, while MOCHI-B simulations satisfyingly reproduced observed inter-patient variability in chronic state prior treatment and SVM dynamics under ETV and IFN monotherapies, the assumption of a system in equilibrium prior treatment is up for discussion since chronic HBV is known to naturally evolve with distinct levels of serum ALT, immune response, serum and intrahepatic viral load, HBeAg status and liver fibrosis (Sandhu et al. ^110^, Fanning et al. ^111^). In this model, the natural evolution of the disease through its various stages (immune-tolerant phase during which patients are HBeAg+, immune-active phase during which patients can become HBeAg-, immune-control and immune-clearance phases) was not accounted for and no mechanistic difference was introduced between HBeAg+ and HBeAg-patients, which discrepancies were accounted for by distinct parameter values only. This strategy enabled the model to implicitly capture the various disease stages with a wide-spectrum of SVM levels and anti-HBV immunity, but can introduce uncertainty in the estimation of treatment efficacy for long treatment periods. Additional modeling effort would be required to better understand differences in terms of disease intensity and treatment response between these populations (Liu et al. ^112^ Na et al. ^113^).

The current model accounts for both innate and adaptive anti-HBV cellular immunity, the importance of which has been widely demonstrated (Biron et al. ^114^, Tsai et al. ^115^, Das and Maini ^116^, Peppa et al. ^117^), as well as for a dual role of HBs and HBe antigens activating not only the immune response but also anti-inflammatory processes leading to an impaired immune response (Revill and Yuan ^118^, Schuch et al. ^19^, Yuen et al. ^39^). However, it doesn’t fully capture the contribution of regulatory T cells (Stoop et al. ^119^) and T cell exhaustion and dysfunction (Fisicaro et al. ^120^) to the impaired anti-HBV immune response. The contribution of B cells and anti-HBV antibodies has attracted increasing attention and it has been suggested that induction of an anti-HBs antibody response could be essential to terminate chronic HBV infection (Ma et al. ^121^). The current model does not account for the humoral immune response; adding this capability would enable the investigation of HBeAg seroconversion with the development of anti-HBe antibodies. As new quantitative data become available, the model could be updated to more comprehensively capture recent knowledge, and thereby further enhance its utility in exploring how to restore a functional anti-HBV host immune response.

Finally, integration of cccDNA within the host genome has not been investigated in the current model due to conflicting evidence at time of model development. Highly sensitive detection methods now allow better characterization of pathogenic aspects of HBV integration (Pollicino and Caminiti ^122^). Recent studies suggest that integrated HBV DNA could act as a stable source of viral RNA and proteins, favoring virus persistence and silencing of host anti-HBV immunity. In particular, integrated HBV DNA could be a major source of HBsAg in HBeAg-negative patients (Wooddell et al. ^123^, Freitas et al. ^124^), which would challenge HBsAg as marker of intrahepatic persistence of infectious cccDNA. Integrated HBV DNA could also contribute to CHB progression to hepatocarcinoma (Pollicino and Caminiti ^122^). Based on novel findings, Tu et al. ^125^ propose a hypothetical knowledge model of HBV DNA integration dynamics which could be tested after refining the model. In a recently published model (El Messaoudi et al. ^126^), integrated HBV DNA was partially accounted for via a second population of productive infected cells containing lower amounts of cccDNA and integrated HBV DNA.

This mechanistic model of HBV treatment and associated analyses are a first step toward better understanding the biological processes driving patient response heterogeneity in chronic hepatitis B, and toward improved therapeutic strategies. Given its mechanistic nature, MOCHI-B can be refined to integrate fast evolving CHB knowledge, in order to increase its utility in the field of CHB research. The model can be used to identify and test new targets and treatment protocols, study patients longitudinally and test hypotheses ahead of confirmatory trials in patients. Future in silico studies would thus allow exploring the contribution of any baseline markers on response to the treatment, in particular those which are difficult to observe in clinical practice.

## Data Availability

The model structure, documentation, and supporting data for this study are accessible in the supplementary material and can also be requested
from the lead contact, C.M., via the jinko.ai platform. The code is likewise available on the jinko.ai platform upon request from the lead contact, C.M.

## Resource availability

### Lead contact

For further information, resource requests, or access to the jink ō platform, please direct inquiries to the lead contact, Claudio Monteiro.

### Data and code availability

The model structure, documentation, and supporting data for this study are accessible in the supplementary material and can also be requested from the lead contact, C.M., via the jinko.ai platform. The code is likewise available on the jinko.ai platform upon request from the lead contact, C.M.

## Acknowledgments

We thank Enyo Pharma for its support of this project. We also thank Bastien Martin, Hippolyte Darré, Simon Arsène, Emmanuelle Bechet, Giulio Foresto, Eliott Tixier, Samuel Laheux, and Ben Illigens for their contributions to the development of this model.

## Author contributions

Conceptualization: S.G.-N., A.S., S.P, E.P., N.R., Y.W. C.M., L.B.; Methodology: S.G.-N., A.S., S.P., E.P., N.R., Y.W., C.M., L.B.; Validation: P.S., P.A.; Formal Analysis: G.G., E.J., R.K., C.M., L.B.; Investigation: S.G.-N., A.S., S.P., E.P., N.R., Y.W., C.M., L.B.; Data Curation: G.G., E.J., R.K.; Writing – Original Draft: S.G.-N., A.S.; Supervision: C.M., L.B.; Writing – Review & Editing: All authors.

## List of abbreviations

BOT: Beginning of treatment
cccDNA: Covalently closed circular DNA
CHB: Chronic Hepatitis B
CTL: Cytotoxic T lymphocyte
EOT: End of treatment
ETV: Entecavir
HBcAg: Hepatitis B core antigen
HBeAg: Hepatitis B envelope antigen
HBsAg: Hepatitis B surface antigens
HBsRNA: Hepatitis B surface antigen related RNAs
HBV: Hepatitis B virus
IFN: Peginterferon-alfa-2a
IL10: Interleukin 10
MOCHI-B: Model of Chronic Hepatitis Infection B
NKC: Natural killer cell
NUC: Nucleotide/side analog
pcRNA: Pre-core RNA
pgRNA: Pre-genomic RNA
rcDNA: Relaxed circular DNA
RF: Random forest
SA: Sensitivity analysis
SOC: Standards of care
SVM: Serum viral markers
VP: Virtual patient
VPOP: Virtual population

## Appendix A. Model description – Literature review & modeling hypotheses

### Appendix A.1. HBV replication cycle

#### Appendix A.1.1. Literature review

HBV replication cycle has been largely reviewed during the past decades (Ko et al. ^127^, Inoue and Tanaka ^47^, Urban et al. ^128^, Lamontagne et al. ^129^, Yuen et al. ^39^, Liang et al. ^130^, Zoulim and Durantel ^131^, Asin-Prieto et al. ^132^, Tang et al. ^133^). Briefly, infectious HBV DNA containing virions transported in blood may enter hepatocytes by binding to surface NTCP receptors followed by endocytosis. Relaxed circular DNA (rcDNA) and HBV polymerase containing virions are uncoated from their HBV surface antigen (HBsAg), HBV envelope antigen (HBeAg) and HBV core antigen (HBcAg) in the hepatocyte cytoplasm and rcDNA and HBV polymerase may enter the hepatocyte nucleus. There, a part of the genetic viral material can integrate the host DNA, and viral DNA is circularized into covalently closed circular DNA (cccDNA), which encodes for all viral RNAs. In the cytoplasm, pregenomic RNA (pgRNA) serves as the messenger for core proteins (HBcAg) and viral polymerase that are assembled with a pgRNA copy into a capsid. The encapsulated pgRNA is then inversely transcripted into double stranded linear DNA (dslDNA) (Cao et al., 2014). dslDNA contained in these capsids may then be recycled and reenter the nucleus of hepatocytes to be relaxed in cccDNA and increase cccDNA content in the nucleus. Proteins, capsids, virions and other subviral particles such as spheres and filaments are created at different steps of the intra hepatocyte replication cycle and secreted in the serum. Precore proteins, encoded by pcRNA, and surface proteins, encoded by different RNAs, enter the secretory pathway through the Golgi and Endoplasmic Reticulum (ER) compartments and are excreted as Hepatitis B envelope antigen (HBeAg), and subviral particles (spheres and filaments) respectively. In parallel, viral capsids containing HBV DNA or HBV RNA may be transported to the Golgi and ER compartments for maturation and covered with surface proteins, before being excreted in the extracellular compartment. Virions secreted in blood may infect any hepatocyte. The number of healthy and infected hepatocytes relies on the ability of circulating virions to infect healthy hepatocytes, of the immune system to eliminate infected cells, of the healthy and infected hepatocytes to divide, and on the vertical transfer of cccDNA during mitosis (Allweiss et al. ^134^, Lutgehetmann et al. ^135^). Previous studies showed slightly reduced mitosis rate for infected compared to healthy hepatocytes (Allweiss et al. ^134^), and the loss of a fraction of cccDNA during mitosis of infected hepatocytes (Lutgehetmann et al. ^135^, Tu and Urban ^136^).

#### Appendix A.1.2. Specific modeling hypotheses

The HBV submodel is designed to account for most biological entities and processes detailed in the previous section (Figure 1), with simplifying assumptions detailed thereafter. The liver is modeled as a pool of healthy and infected hepatocytes, whose relative proportions may evolve over time. HBV replication cycle is assumed to be identical for all infected hepatocytes. The time scale at the cell level is negligible compared to the time scale considered for simulations (months, years) which leads to consider the infection state of infected cells as identical and homogeneous within the liver. At the hepatocyte level, organelles are not represented and the replication cycle is modeled in one compartment equivalent to an hepatocyte. In particular, nucleus, Golgi and ER compartments are not modeled, nor is DNA integration in the host genome. Besides, only virions containing cccDNA and rcDNA forms of DNA are represented, virions containing dsDNA are not modeled nor are naked capsids whose secretion is modeled by the consumption of HBcAg proteins within hepatocytes. rcDNA virions may either be amplified or excreted. The model does not distinguish between viral spheres and filaments, but instead represents a simplified synthesis and secretion of HBsAg subviral particles. Virions and subviral particles are assumed to be directly secreted in the blood compartment in an independent manner (Garcia et al. ^137^) and the secretion regulation is assumed to be driven by the transcription step. For the sake of simplicity, regulation of gene transcription by HBx protein, HNF-4*α* activities or enhancers are not modeled. Regarding cell renewal, we assume that the death rate is independent of the infected state of the cell. Cell proliferation is modeled to balance cell death by mitosis (i.e. the higher a cell-population decrease is, the higher its regeneration is) respecting the current ratio of healthy and infected cells and assuming that infected hepatocytes prolifer 2.6 times slower than healthy hepatocytes as reported by Allweiss et al. ^134^. The model also accounts for a fraction of intrahepatic cccDNA that can be lost during mitosis of infected cells. All related variables, parameters and equations are described in Tables D.2, D.3, D.4 and D.5.

### A.2. Appendix Immune response Appendix

#### A.2.1. Literature review

The role of the immune system in pathogenesis of CHB has been largely reviewed in the past years (Bertoletti and Gehring ^46^, Ferrari ^138^, Tan et al. ^139^, Revill and Yuan ^118^, Chisari and Ferrari ^42^, Das and Maini ^116^, Busca and Kumar ^140^). A brief description of the immune response to HBV infection and main factors leading to development to chronicity is proposed thereafter.

Early after infection, HBV acts as a stealth virus with HBV DNA and HBV antigens undetectable in both serum and liver for 4-7 weeks post infection (Ferrari ^138^). As a consequence, early HBV infection induces a poor innate intracellular response (Wieland et al. ^141^, Fletcher et al. ^142^, Wieland and Chisari ^143^) and natural killer cells (NKC) activation is often delayed (Dunn 2009). Following active viral replication, the presence of HBV antigens and infected hepatocytes induces activation of immune effector cells including NKC and T lymphocytes, and within 6-12 weeks (Dunn et al. ^144^), 95% of adult patients develop an immune response efficiently acting for the resolution of the infection and detected as acute infection. NKCs play a crucial role in antiviral immune defense (Waggoner et al. ^40^, Björkström et al. ^41^). They are innate immune cells activated by stress signals on infected hepatocytes and liver dendritic cells or by direct recognition of viral components (Bertoletti and Gehring ^46^). They actively participate in the resolution of the infection via direct cytotoxicity and production of antiviral and proinflammatory cytokines including IFN-*γ* and TNF*α* (Chen et al. ^145^, Kakimi et al. ^146^) causing inhibition of HBV replication and recruitment of immune cells. NKCs also play a crucial role in antiviral immunity by bridging innate and adaptive response (Biron et al. ^114^). Regarding adaptive response, major effector cells involved in response to viruses are T lymphocytes. As such, control of acute HBV infection relies on a robust CD4+ Type 1 and CD8+ HBV-multispecific polyclonal T cell response (Chisari and Ferrari ^42^, Chisari et al. ^43^, Maini et al. ^44^). Activation of HBV-specific T lymphocytes in lymph nodes draining the site of infection is mediated by dendritic cells (DCs) previously activated by HBV virions and viral proteins in the liver (Bertoletti and Ferrari ^147^, Li et al. ^148^) and pro-inflammatory cytokines produced by activated innate immune cells (Brunetto and Bonino ^49^). CD8+ cytotoxic lymphocytes (CTL) play a key role in clearance of HBV as well as infected cells via cytolysis of HBV-infected cells and secretion of cytokines such as IFN-*γ* and TNF-*α* (Guidotti and Chisari ^149^). CTLs are also responsible for hepatocellular injury (Ando et al. ^150^, Tsui et al. ^151^). CD4+ helper T cells enhance activation of the humoral response, which appears 10-12 weeks after HBV infection (Fong 1994) and yields to the production of antibodies mediating a protective immunity to prevent a reinfection by blocking the binding ability of virus on target cells (Rehermann and Nascimbeni ^152^, Huang et al. ^153^). Upon resolution of the infection, regulatory mechanisms appear to prevent excessive liver damage and inflammation inhibiting immune cells’ cytolitic activity as well as cytokine secretion (Ferrari ^138^). In particular, immunosuppressive cytokines such as IL-10 can inhibit both innate and adaptive immunity (Das and Maini ^116^).

In contrast to adult patients, over 95% of neonates and 20-30% of children (aged 1-5 years) develop CHB disease following HBV infection (Beasley ^154^). CHB is a heterogeneous disease that can vary greatly in the level of viral replication, liver disease and immune responses (Bertoletti and Gehring ^46^). Studies have shown that HBV is able to escape from surveillance of the host innate immune system (Busca and Kumar ^140^, Ma et al. ^155^). Secretory HBV proteins (HBcAg, HBsAg, HBeAg) have been reported to impair antiviral control notably through suppression of TLR expression (Wu et al. ^156^, Visvanathan et al. ^157^) and induction of higher levels of regulatory T cells (Treg) (Loggi et al. ^158^) and IL-10 (Hyodo et al. ^45^, Bertoletti and Gehring ^46^, Inoue and Tanaka ^47^) which in turns suppresses adaptive immune response. Usually, chronic patients lack the acute phase immune response and subsequently fail to prime an adequate adaptive response (Bertoletti and Ferrari ^147^, Revill and Yuan ^118^). Additionally, studies suggest that in CHB infection NK cells are functionally impaired (de Martino et al. ^159^) and that CD4+ and CD8+ T cell responses are characterized by weak or undetectable virus-specific T cell levels (Sobao et al. ^160^, Webster et al. ^161^, Chang et al. ^162^). Therefore, it is likely that viral chronicity alters the repertoire of HBV-specific immunity to a level that makes its functional restoration very complex (Bertoletti and Gehring ^46^). The immunological features of the liver might also contribute to the maintenance of immunological tolerance present in chronic HBV infection (Bertoletti and Gehring ^46^). Indeed, the liver facilitates tolerance rather than immunoreactivity, which protects the host from antigenic overload of dietary components and drugs derived from the gut (Bogdanos et al. ^163^).

#### Appendix A.2.2. Specific modeling hypotheses

The immune system (IS) submodel is designed to model the IS response to CHB infection and accounts for cytolysis of infected hepatocytes and degradation of viral components by immune cells and cytokines. The immune system submodel relies on the following assumptions and modeling hypotheses. Regarding innate immunity, NKC are the only cell-type modeled. NKC are activated by viral antigens (serum HBsAg) and IFN treatment and inhibited by IL-10. They produce proinflammatory cytokines, modeled as IFN-*γ*, which in turn enhances T cells’ activation. Regarding adaptive immune response, the model focuses on HBV-specific CTL. CTL are activated by HBsAg and IFN-*γ* and inhibited by IL-10. NKC and CTL both act through cytolytic (lysis of infected cells) and non cytolytic pathways to clear HBV infection (degradation of cccDNA induced by IFN-*γ* signaling leading to a decrease in the pool of cccDNA in the infected hepatocyte). In addition, phagocytosis is modeled to account for viral particle degradation in the serum. The action of Kupffer cells and other phagocytizing cells degrading viral particles is modeled via a phagocytosis reaction enhanced by HBV antigens (serum HBsAg) and pro-inflammatory cytokines (IFN-*γ*) and inhibited by IL-10. In order to model HBV ability to impair host immunity, we model a production of the anti-inflammatory cytokine IL-10 enhanced by viral antigens (serum HBsAg and HBeAg). Antigen presenting cells, regulatory T cells, humoral response and anti-HBV antibody production are not modeled implicitly nor are lymphatic circulation of immune cells and draining lymph nodes. Instead the immune response is modeled in the liver.

#### Appendix A.3. Drugs pharmacokinetics

Entecavir (ETV) and peg-interferon alfa 2a (IFN) pharmacokinetics models were based on a standard PBPK model, accounting for drug administration, distribution, metabolism and excretion assuming a physiologically realistic compartmental structure, as described in Jones et al. ^164^. Implemented PK compartments, mechanisms and variables are schematized Figure E.6. ETV was considered to be administered as a solution therefore no dissolution mechanisms were implemented. As ETV is orally administered, it transits in the different segments of the gut and is absorbed into enterocytes of the small intestine. Distribution kinetics were assumed to be perfusion-limited in all organs except the gut where it was described as permeability-limited since active transports for ETV were found to be negligible (Hsueh et al. ^165^). ETV goes from the enterocyte to the vascular and interstitial part of the liver through the portal vein. Non renal clearance was neglected (Hsueh et al. ^165^). Specific hypotheses were made for the modeling of IFN PK, as previously published in Offman and Edginton ^48^. In particular, regarding administration, the skin compartment was subdivided into depot, interstitial (to account for sub cutaneous administration) and vascular. Regarding distribution, for each organ, a fraction of the drug (computed based on the vascular reflection coefficient of each organ) is transported from plasma to the interstitial compartment where, the drug then transits into lymphatic supply (lymphatic reflection coefficient) where it is carried into arterial supply for distribution into other organs (Adipose, Heart, Lung, Brain, Bone, Skin, Kidney, Liver, Muscle, Gut, Pancreas and spleen) according to the plasma flow rate supplying each organ (perfusion-limited kinetics). IFN clearance was modeled via renal and non-renal pathways (in both interstitial and vascular compartments)(Shuldiner et al. ^50^).

#### A.4. Appendix Drugs’ mechanisms of action Appendix

##### A.4.1. Literature review

ETV is a deoxyguanosine analogue. ETV treatment inhibits the HBV polymerase (Langley et al. ^51^) and consequently prevents capsid maturation necessary for the viral replication cycle. ETV triphosphate (ETV-TP) displays activity against all three synthetic activities of the HBV polymerase, i.e., the unique protein-linked priming activity, RNA-directed first-strand DNA synthesis or reverse transcription, and second-strand DNA-directed DNA synthesis (Langley et al. ^51^).

IFN is an antiviral drug with multiple mechanisms of action including immunomodulatory activities and to a lesser extent direct antiviral and antiproliferative effects (Boortalary et al. ^166^, Borden et al. ^167^). IFN induces a non-virus-specific immune response in both infected and noninfected cells through upregulation of many IFN-stimulated genes (ISGs) that encode antiviral effector proteins (Sadler and Williams ^168^). IFN also interacts with adaptive and innate immune responses to promote dendritic cell maturation, stimulate natural-killer cell activation, proliferation and antiviral potential (Micco et al. ^169^), enhance memory T cell proliferation and prevent T cell apoptosis (Boni et al. ^170^).

Several studies have also suggested that IFN could interfere with several steps of viral replication, notably by blocking core particle formation, accelerating the degradation of core particles and pregenomic RNA (Brunetto and Bonino ^49^, Wieland et al. ^171^, Xu et al. ^172^, Li et al. ^173^), and repressing HBV cccDNA transcriptional activity by epigenetic modification (Belloni et al. ^174^, Brunetto and Bonino ^49^). Some reports have proposed an additional direct action on cccDNA degradation [maybe a publication from Xia (Interferons induce degradation of HBV cccDNA] with a significant decrease in cccDNA levels after IFN treatment (Chuaypen et al. ^175^, Mu et al. ^176^). However, this direct impact on cccDNA is subject for debate with studies reporting that cccDNA remained at similar levels in IFN-treatment hepatocytes (Niu et al. ^177^, Shen et al. ^178^, Mutz et al. ^179^).

##### Appendix A.4.2. Modeling hypotheses

ETV effect was modeled by the inhibition of the reaction producing rcDNA capsids from pgRNA capsids as viral polymerase is needed for the retro-transcription to occur. Specifically, this effect was modeled by an inhibition term in the reaction of rcDNA capsid formation with a parameter defining the maximal effect of ETV and a Hill function dependent on ETV concentration in the liver. To model IFN immunomodulatory impact, the model accounted for an additional activation rate of innate immune cells by IFN treatment. Regarding IFN direct antiviral effect, we assumed that IFN’s main impact was to repress HBV cccDNA transcriptional activity into pgRNA and others mRNA species (preS/S RNA) in infected hepatocytes (Brunetto and Bonino ^49^). These effects were modeled by an inhibition term in the viral RNAs (pgRNA and HBsRNA) production reaction.

## Appendix B. Model implementation and parametrization

### B.1. Appendix Literature data used to calibrate the model

A literature review was conducted using online electronic databases including Pubmed and Google Scholar. Data were extracted from original publications when available or digitalized from figures using WebPlotDigitizer Rohatgi A. ^180^.

In order to calibrate CHB pathophysiology submodels (HBV viral replication and immune response), the literature was screened to gather information and data related to viral replication following HBV infection in vitro and in vivo and chronic patients evolution of serum and intra-hepatic viral markers and immune markers over time. Notably, data on pathophysiology of HBV infection were obtained from observations in humans on viral incubation periods and SVM time-course following HBV infection World Health Organization website, Ganem and Prince ^181^ and Trépo et al. ^182^. Data on CHB disease were obtained from quantification of SVM concentrations including HBV DNA, HB-sAg and HBeAg (Ganem and Prince ^181^, Luckenbaugh et al. ^183^, Wang et al. ^93^, Gao et al. ^184^) and intrahepatic HBV viral components including HBV covalently closed circular DNA (cccDNA), pregenomic RNA (pgRNA), precore RNA (pcRNA) and relaxed circular DNA (rcDNA) levels of per cell (Laras et al. ^91^, Volz et al. ^92^, Dandri et al. ^89^, Wang et al. ^185^) in treatment-naive HBe-antigen-positive (HBeAg+) or negative (HBeAg-) CHB patients. Data on immune system response to HBV infection were obtained from quantification of immune markers (immune cells, cytokines) in healthy and HBV infected humans (Huhn et al. ^67^, Saxena et al. ^68^, Arababadi et al. ^96^).

PK of ETV was based on a study by Yan et al. ^72^ where ETV plasma concentrations were measured in 18 healthy subjects administered with ETV 0.1, 0.5 or 1.0 mg QD (tablets) daily for 14 consecutive days and on a study by Chen et al. ^186^ where plasma, liver and kidney concentrations were measured in 5 rats administered with intragastric ETV 0.09 mg/kg daily during 10 days. The time-course of IFN PK was extracted from a study by Costa (2018) where serum IFN concentrations were measured in 31 healthy subjects administered with a single dose of IFN 180 *µ*g s.c. and additional information on IFN PK including maximal and trough concentrations as well as serum IFN accumulation following regular weekly IFN administrations were obtained from the FDA and EMA IFN drug description.

Regarding efficacy of ETV and IFN monotherapies, the literature was screened for published clinical studies reporting SVM baseline levels and SVM evolution over time in HBeAg+ or HBeAg-CHB patients treated with either an ETV monotherapy (0.5 mg QD p.o. for at least 48 weeks) or IFN monotherapy (180 *µ*g QW s.c. for 48 weeks) and summarized or population-level data were extracted. An overview of these clinical studies is provided in Table 1.

## Appendix C. Statistical analyses

### Appendix C.1. Fractional factorial design

A fractional factorial design was defined using the AlgDesign (R package - Cran ^187^) which implements a modified Federov algorithm. The Federov algorithm consists in starting from a large design that maximizes the determinant of the information matrix. The final design is obtained iteratively by selecting a candidate design that maximizes the amount of change of information in the initial design. Because the computational cost of the above algorithm became prohibitive for a large number of parameters, the Federov algorithm was modified by randomly drawing a subset of nCand candidate samples at each iteration. The obtained design may represent a local optimum. Therefore, the whole procedure was repeated nRepeats times and the best design across all repetitions was kept.

## Appendix D. Table of model parameters, variables, fluxes

**Table D.2:**
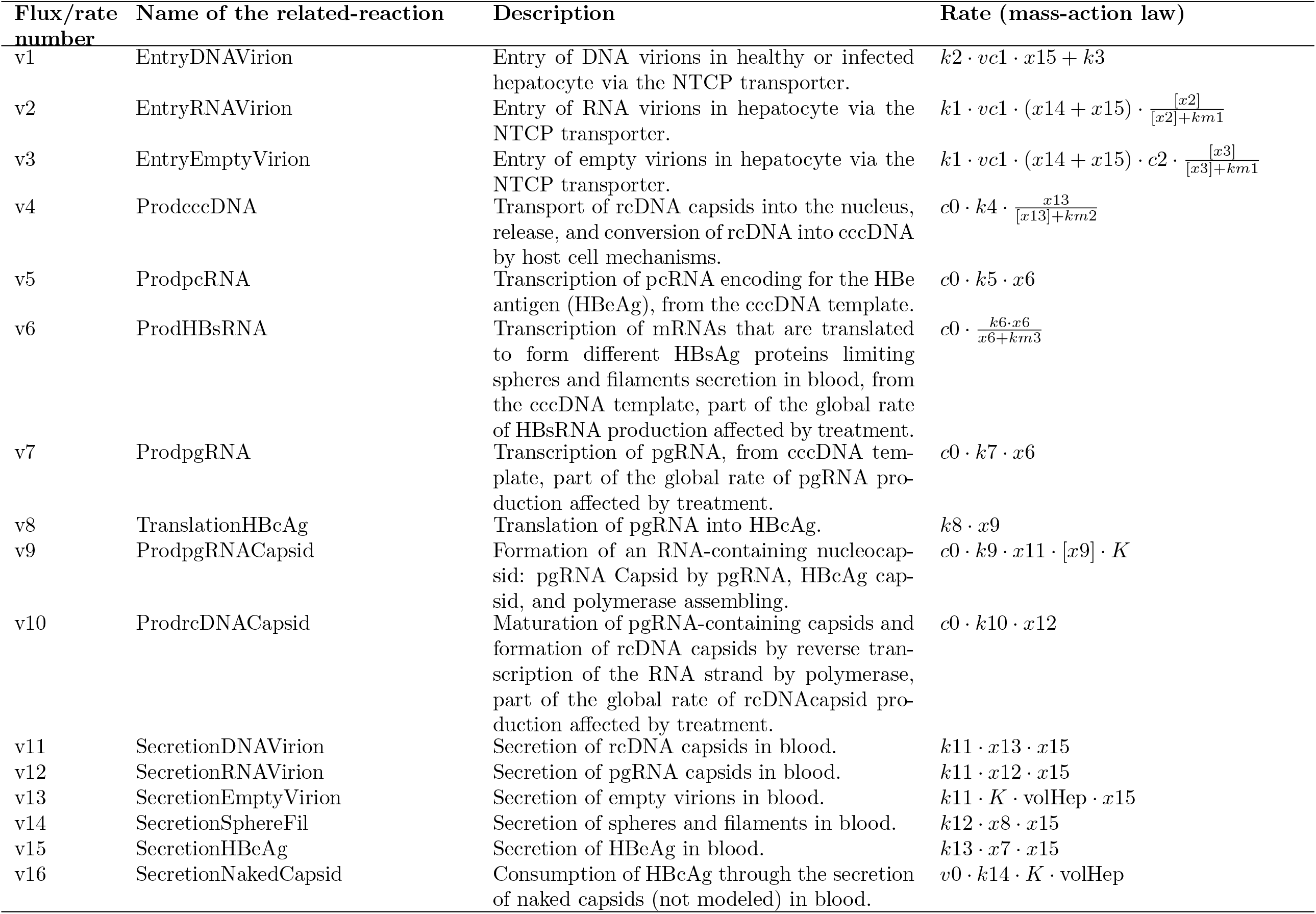
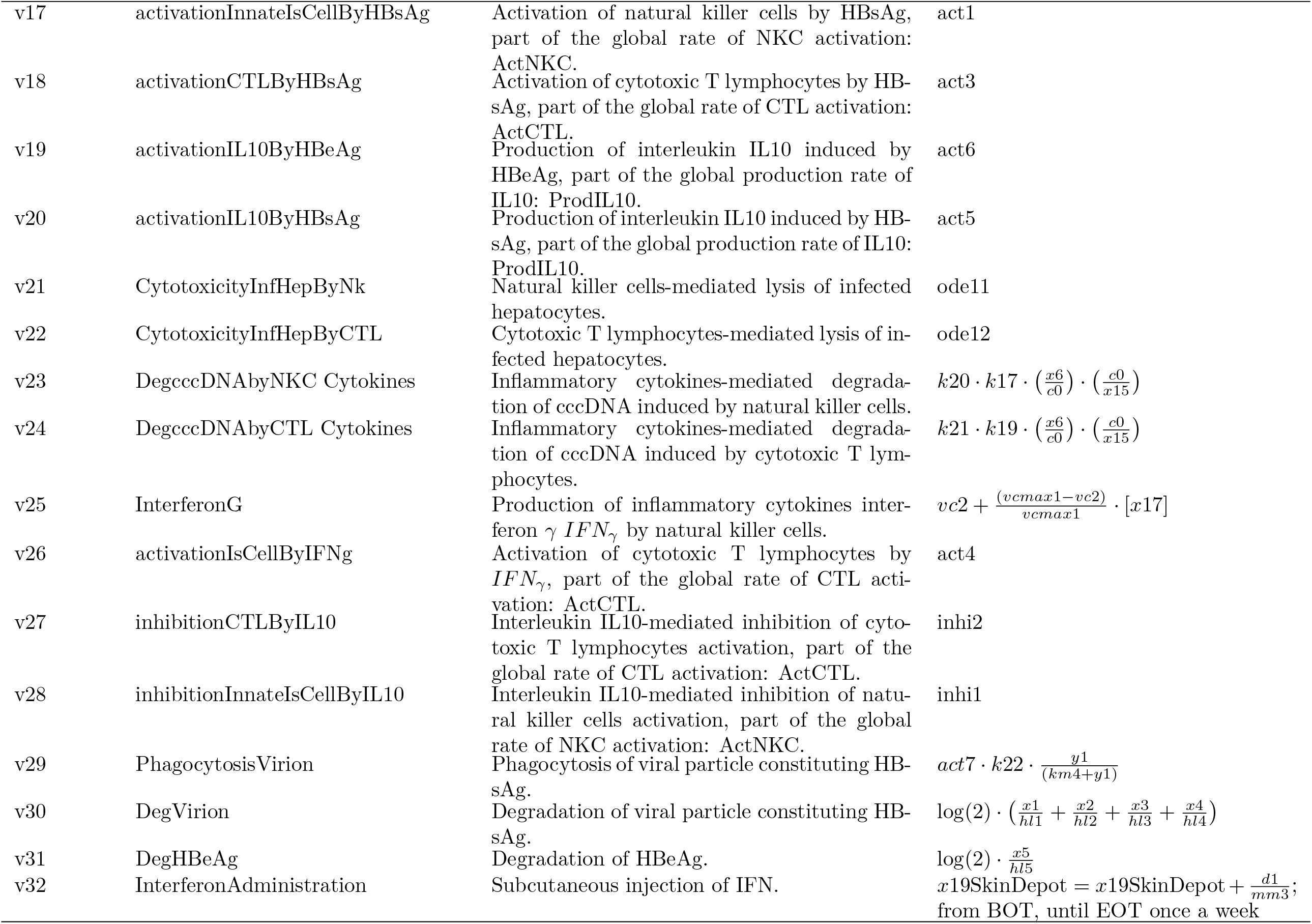
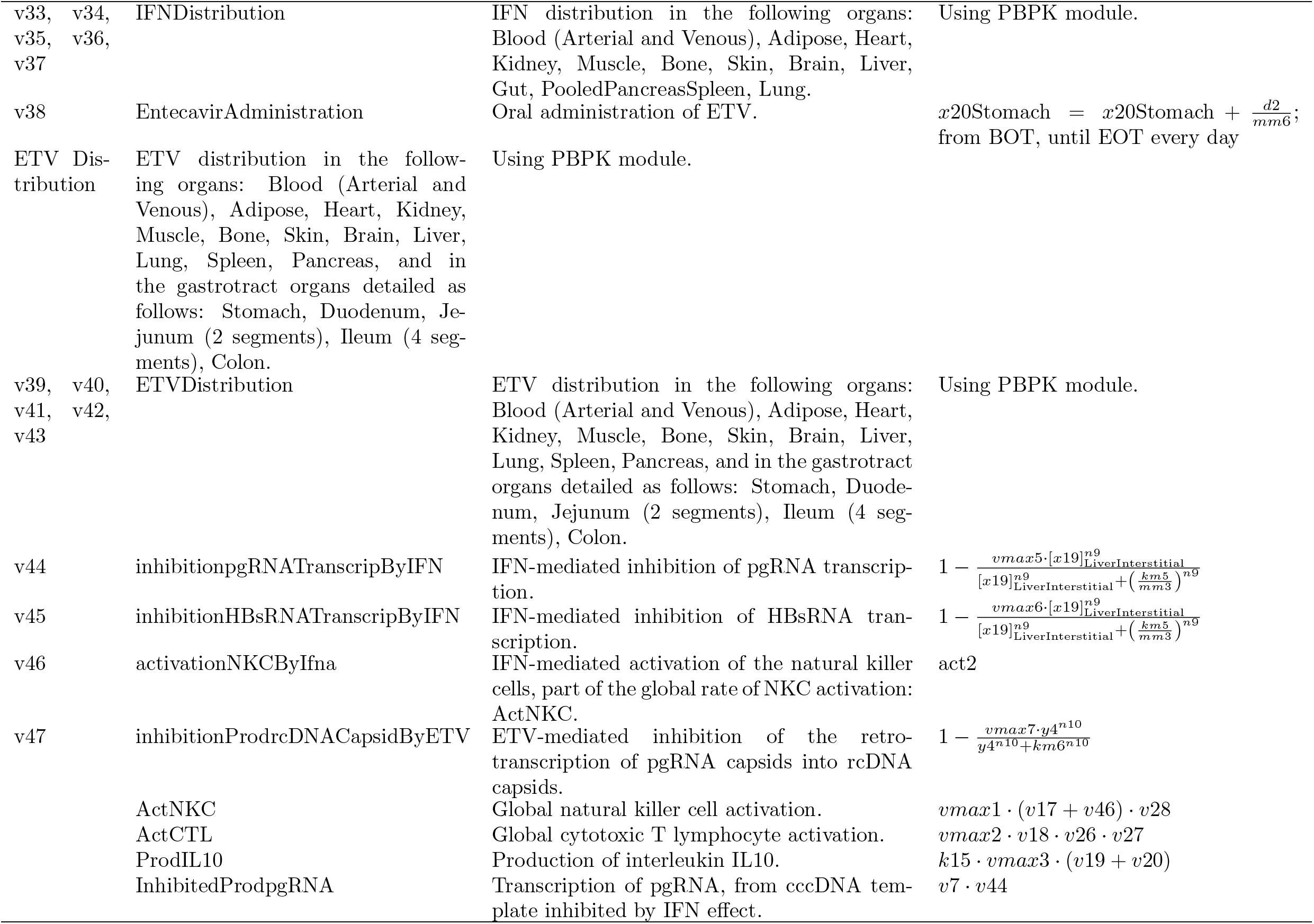
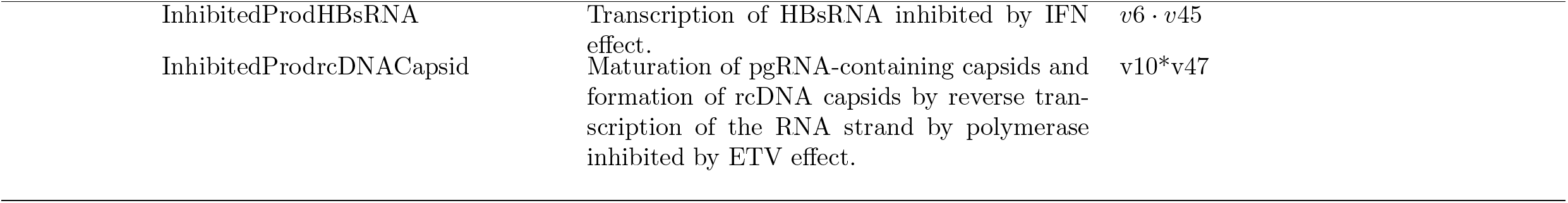
Table of model parameters, variables, fluxes.

**Table D.3:** Table of model odes.

| ODE number | Name of the related ODE | Description | Rate law |
| --- | --- | --- | --- |
| ode1 | infectionInfHep | Healthy cells are infected upon virion entry | $dx15/dt = k3$ |
| ode2 | infectionHealthyHep | Healthy cells are infected upon virion entry | $dx14/dt = -k3$ |
| ode3 | infectionMediatedDilutionrcDNA Capsid | Dilution of rcDNACapsid upon infection of healthy cells | $\frac{dx13}{dt} = (1 - x13) * k3/x15$ |
| ode4 | proliferationInfHep | Proliferation of infected cells to return to the basal amount respecting the current ratio of healthy and infected cells | $dx15/dt = k23 * nb1 * ((nb1 - x15 - x14)/nb1) * y7/r3$ |
| ode5 | proliferationHealthyHep | Proliferation of healthy cells to return to the basal amount respecting the current ratio of healthy and infected cells | $dx14/dt = k23 * nb1 * ((nb1 - x15 - x14)/nb1) * y8$ |
| ode6 | naturalMitosisMediated DilutioncccDNA | Dilution of cccDNA upon mitosis induced by natural cell death | $dx6/dt = -(1 + f1) * x6 * (y9 * y7)/x15$ |
| ode7 | naturalDeinfectionHealthy Hep | Deinfection of infected hepatocytes by loss of cccDNA related to natural death-induced proliferation | $dx14/dt = k24$ |
| ode8 | naturalDeinfectionInfHep | Deinfection of infected hepatocytes by loss of cccDNA related to natural death-induced proliferation | $dx15/dt = -k24$ |
| ode9 | infectionMediatedDilution ODEs | Dilution of intrahepatic entities upon infection of new cells. Intrahepatic entities: cccDNA, pgRNA, pcRNA, HBsRNA, HBcAg, HBVpol, pgRNACapsid. | $d[entity]/dt = -[entity] * k3/x15$ |
| ode10 | naturalMitosisMediated Dilution-ODEs | Dilution of intrahepatic entities upon mitosis induced by natural cell death. Intrahepatic entities: pgRNA, pcRNA, HBsRNA, HBcAg, HBVpol, pgRNACapsid, rcDNACapsid. | $d[entity]/dt = -([entity] * y9 * y7/x15)$ |
| ode11 | cytotoxicityInfHepByNk | NK cell mediated cytotoxicity | $dx_{15}/dt = -k_{17}$ |
| ode12 | cytotoxicityInfHepByCTL | CTL mediated cytotoxicity | $dx_{15}/dt = -k_{19}$ |
| ode13 | iSMitosisMediatedDilution cccDNA | Dilution of cccDNA upon mitosis induced by immune system killing | $dx_6/dt = -(1 + f_1) * x_6 * (k_{17} + k_{19}) * y_7/x_{15}$ |
| ode14 | iSMediatedDeinfection HealthyHep | Deinfection of infected hepatocytes by loss of cccDNA related to is-mediated killing-induced proliferation | $dx_{14}/dt = k_{25}$ |
| ode15 | iSMediatedDeinfection InfHep | Deinfection of infected hepatocytes by loss of cccDNA related to is-mediated killing-induced proliferation | $dx_{15}/dt = -k_{25}$ |
| ode16 | isMitosisDilutionODEs | Dilution of intrahpetic entities upon mitosis induced by immune system killing. Intra-hepatic entities: pgRNA, pcRNA, HBsRNA, HBcAg, HBVpol, pgRNACapsid, rcDNACapsid. | $d[entity]/dt = -([entity] * (k_{17} + k_{19}) * y_7/x_{15})$ |

**Table D.4:** Summary of variables implemented in MOCHI-B. The variables name column refers to variable names used in Table S1 to detail the rate of each flux under a mass-action law.

| Variable | Variable name | Variable definition | Conventions |
| --- | --- | --- | --- |
| x1 | DNAHBV | HBV DNA virion in the serum. | variable in its compartment. $[x]_{Organ}$ : drug molar concentration in the Organ compartment $x_{Organ}$ : drug quantity in the Organ compartment |
| x2 | RNAHBV | HBV RNA virion in the serum. |  |
| x3 | EmptyHBV | HBV empty virion in the serum. |  |
| x4 | SphereFil | HBV sphere and filaments in the serum. |  |
| x5 | HBeAg | HBe Antigen in the serum. |  |
| x6 | cccDNA | Intrahepatic cccDNA. |  |
| x7 | pcRNA | Intrahepatic pre-core RNA. |  |
| x8 | HBsRNA | Intrahepatic HBsRNA. |  |
| x9 | pgRNA | Intrahepatic pregenomic RNA. |  |
| x10 | HBcAg | Intrahepatic protein HBcAg. |  |
| x11 | HBVpol | Intrahepatic HBV polymerase. |  |
| x12 | pgRNACapsid | RNA-containing capsid in the hepatocyte. |  |
| x13 | rcDNACapsid | DNA-containing capsid in the hepatocyte. |  |
| x14 | Nhealthy | Number of healthy hepatocytes in the liver. |  |
| x15 | Ninfected | Number of infected hepatocytes in the liver. |  |
| x16 | CTL | Activated cytotoxic CD8+ T cells in the liver. |  |
| x17 | NKC | Activated Natural Killer cells in the liver. |  |
| x18 | IL10 | Interleukin-10 in liver vascular. |  |
| x19 | PegIfn2a | Pegylated interferon alpha (treatment). |  |
| x20 | ETV | Entecavir (treatment). |  |

**Table D.5:**
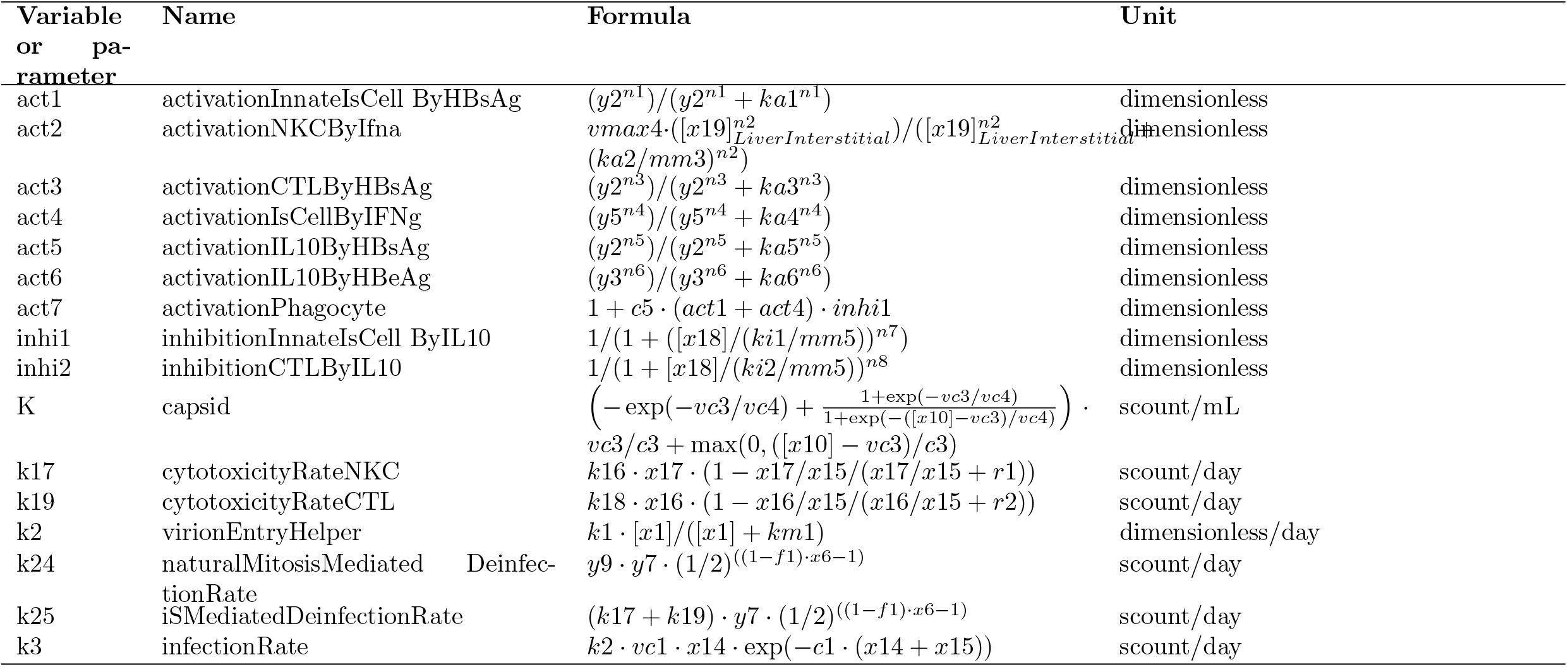
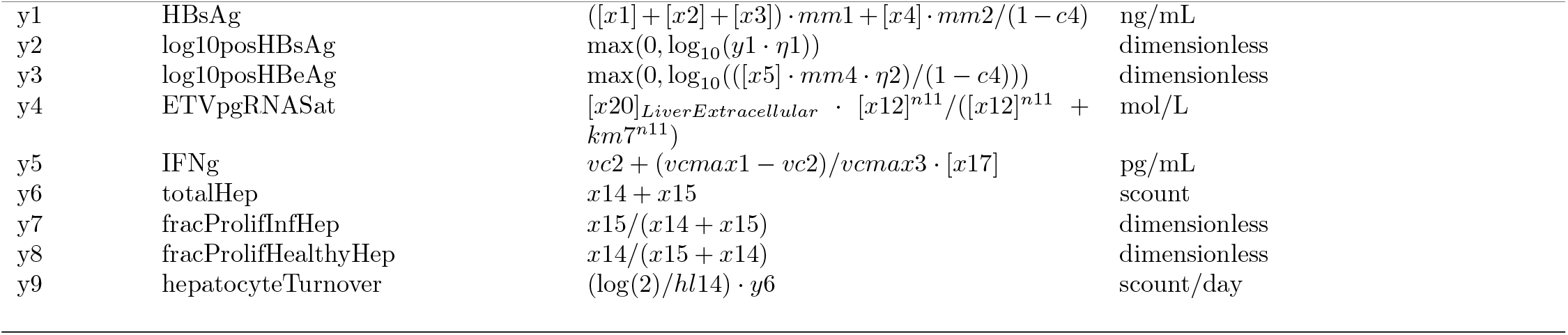
Summary of the intermediate variables and parameters implemented in MOCHI-B. The variables name column refers to variable names used in Table S1 to detail the rate of each flux under a mass-action law.

**Table D.6:**
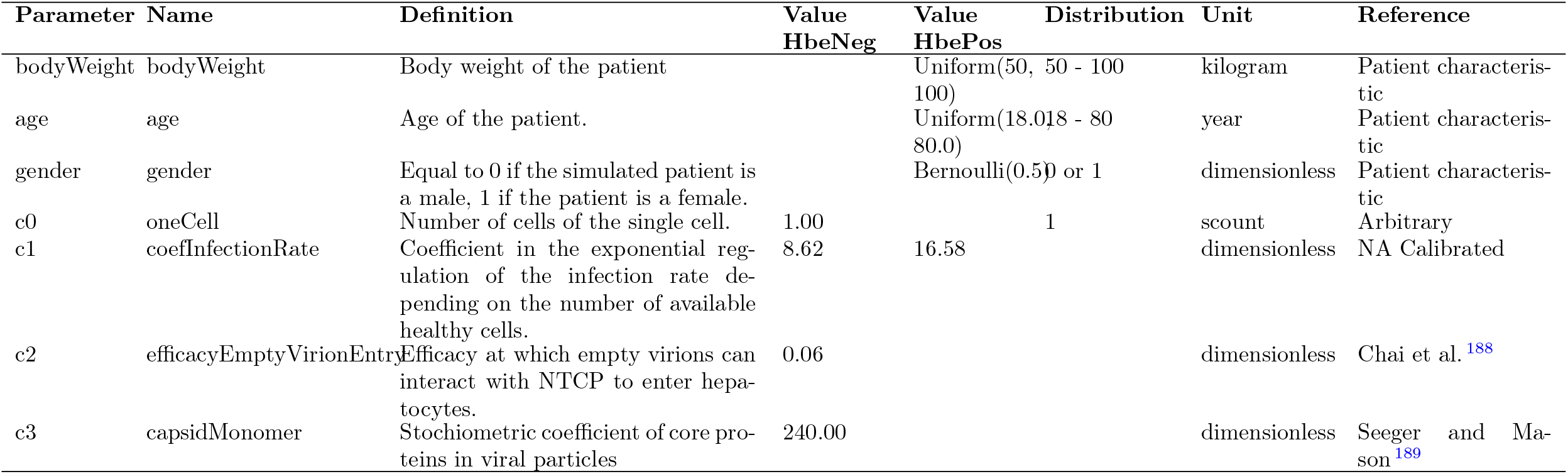
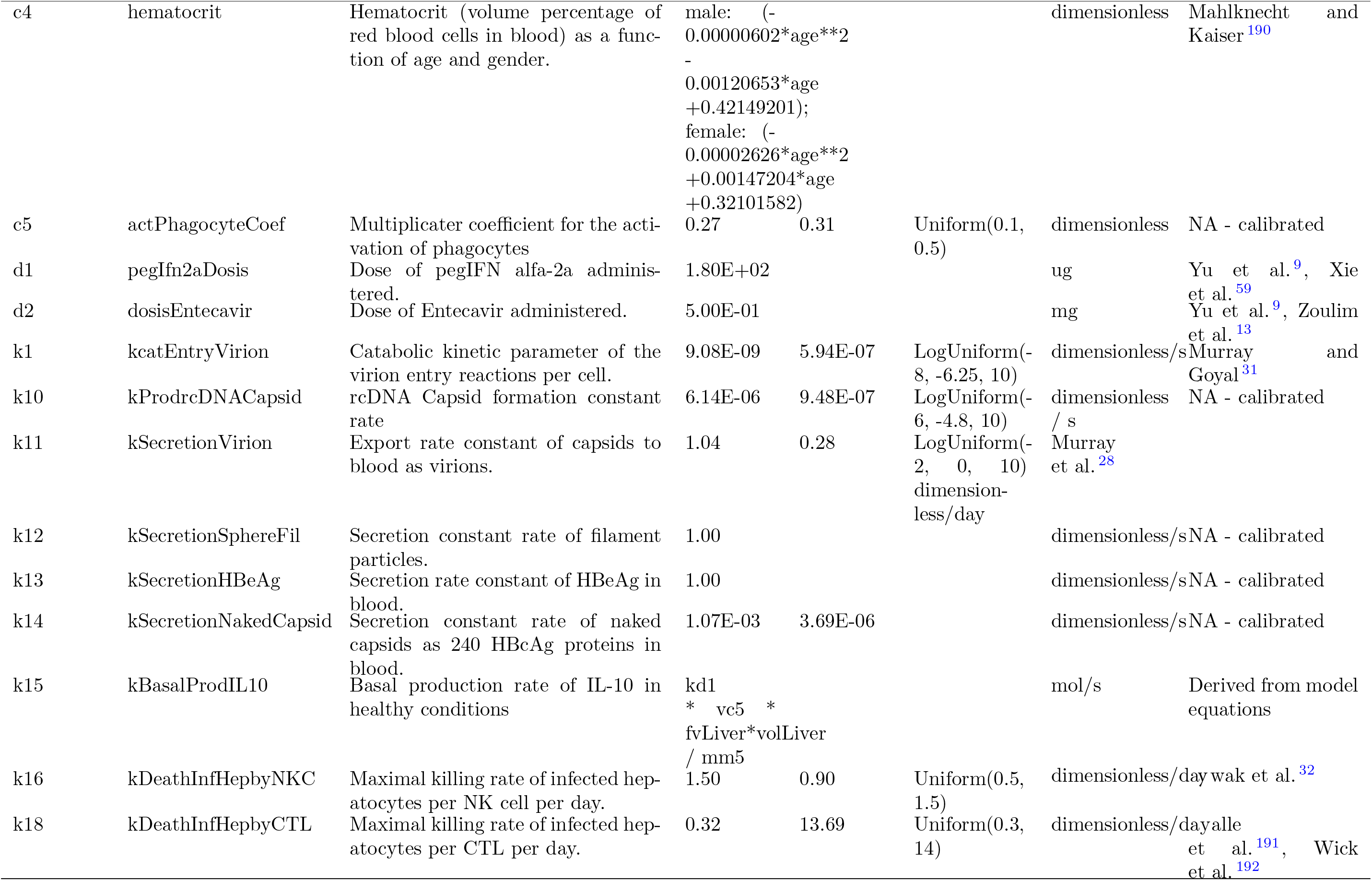
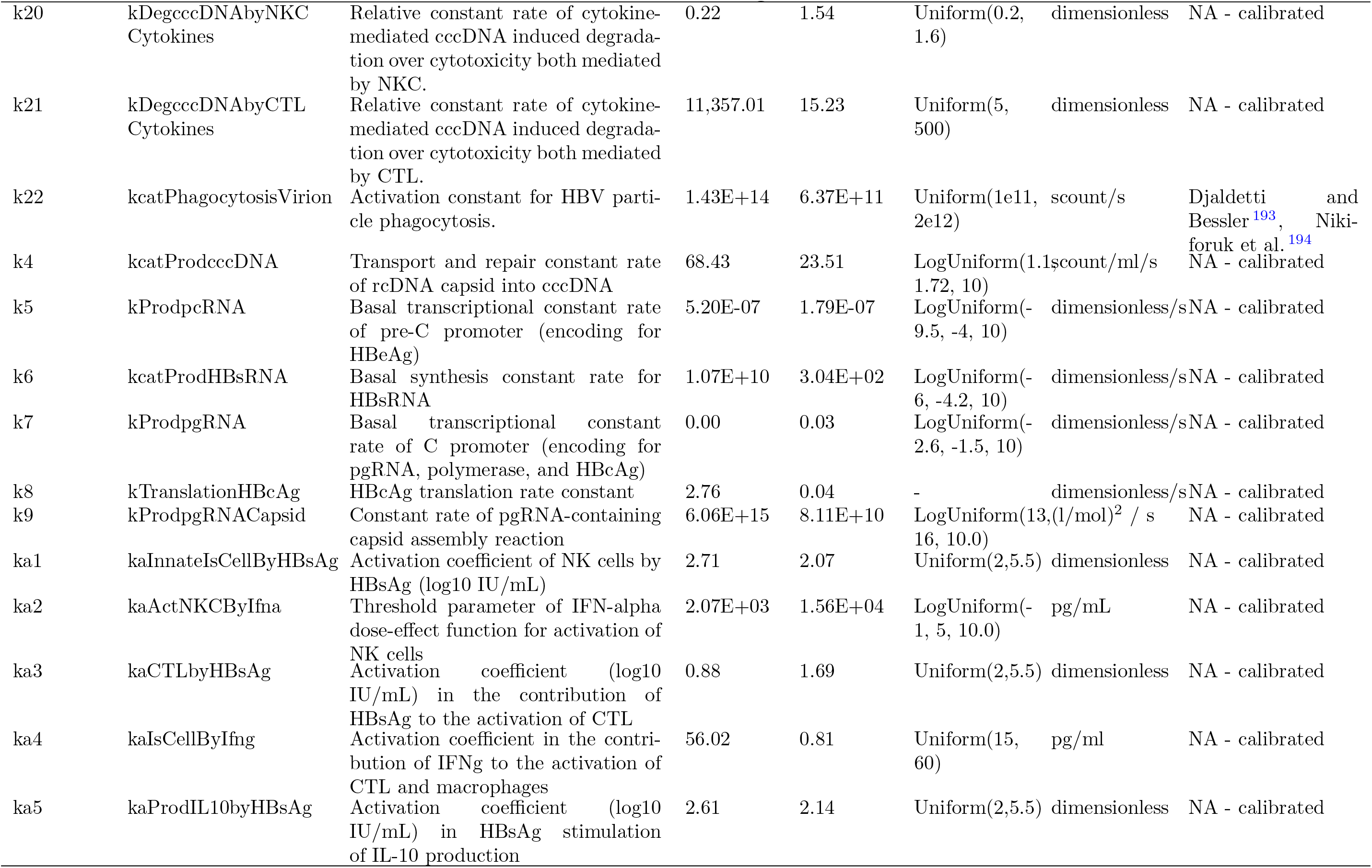
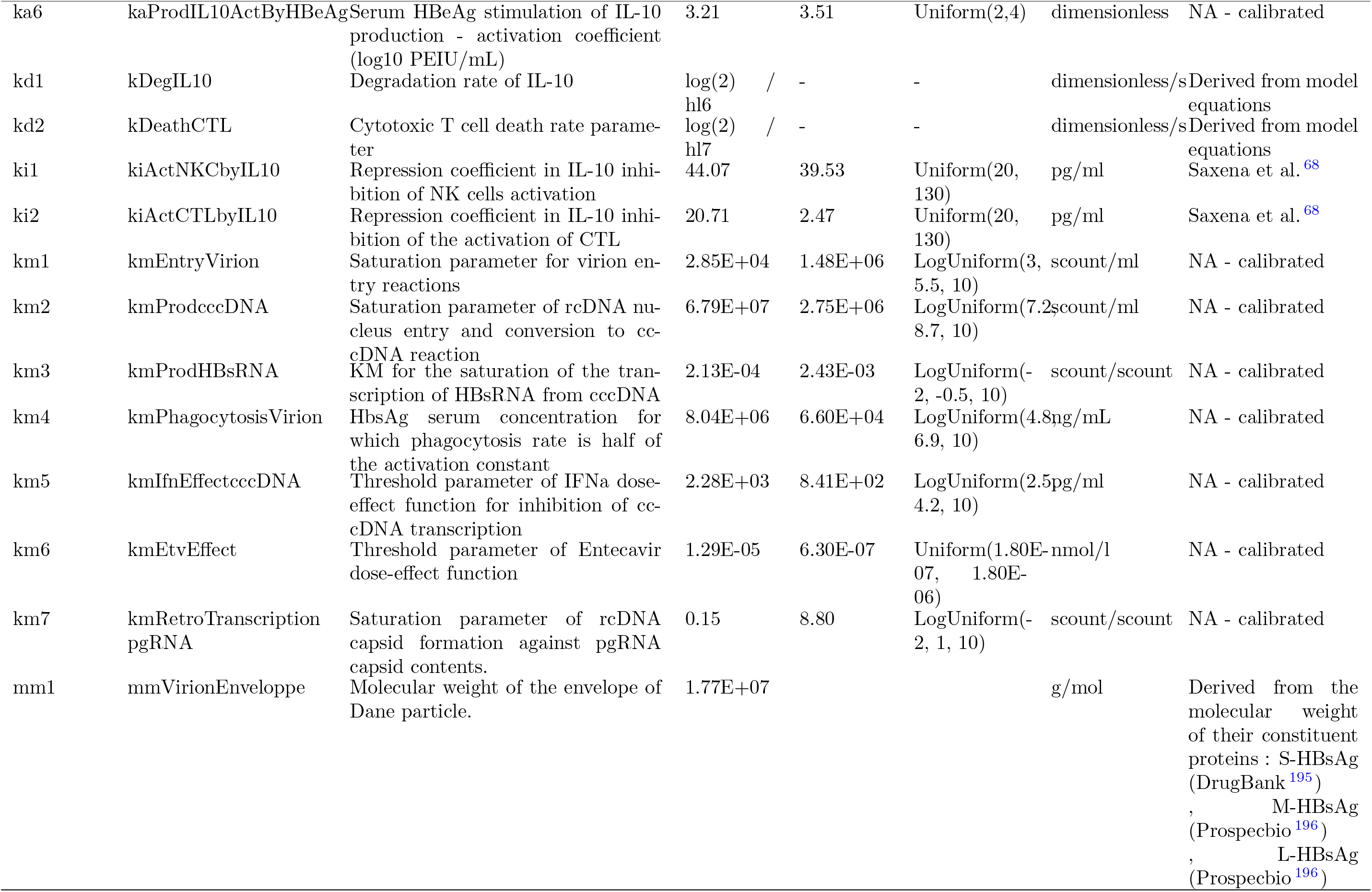
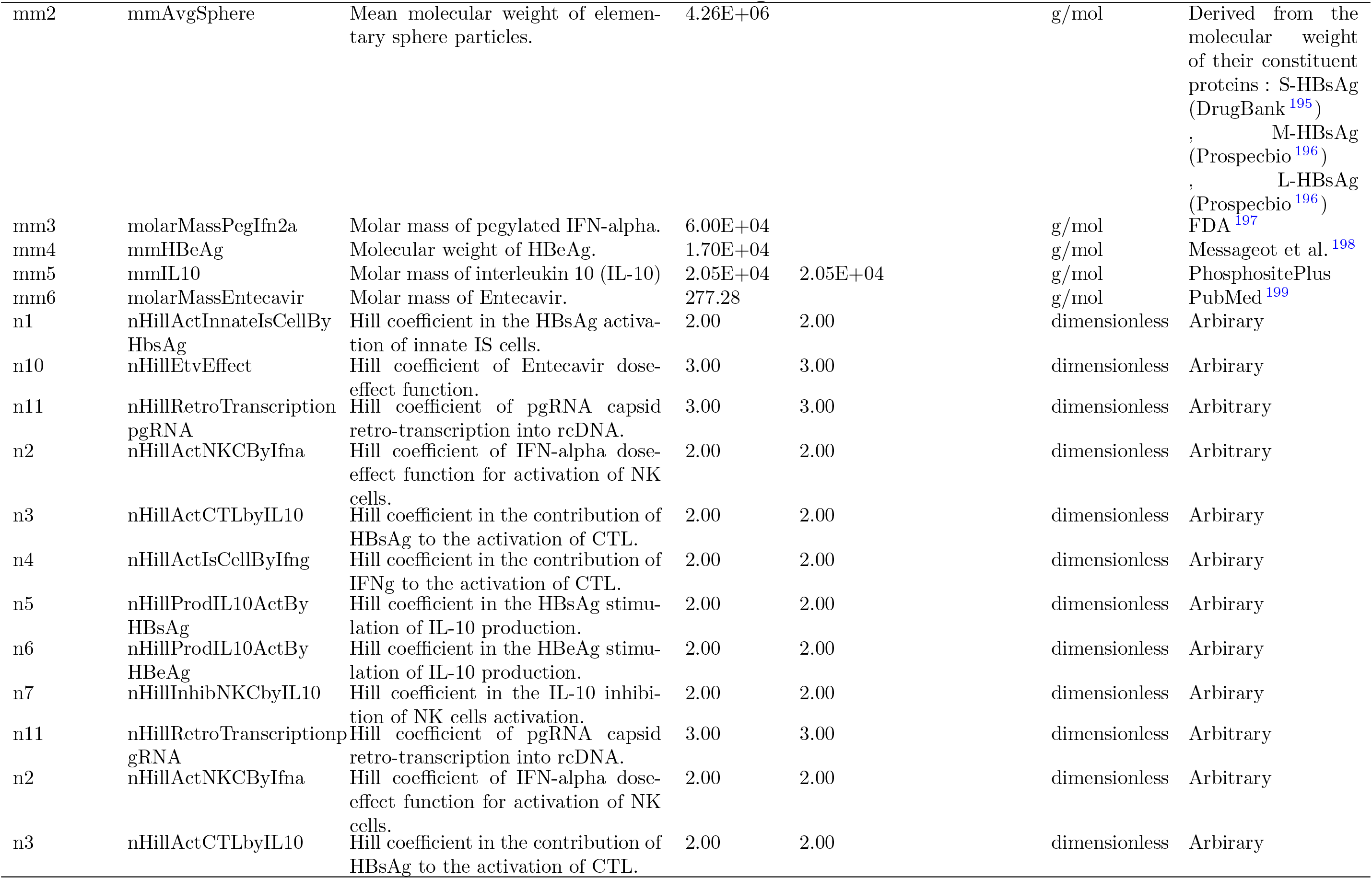
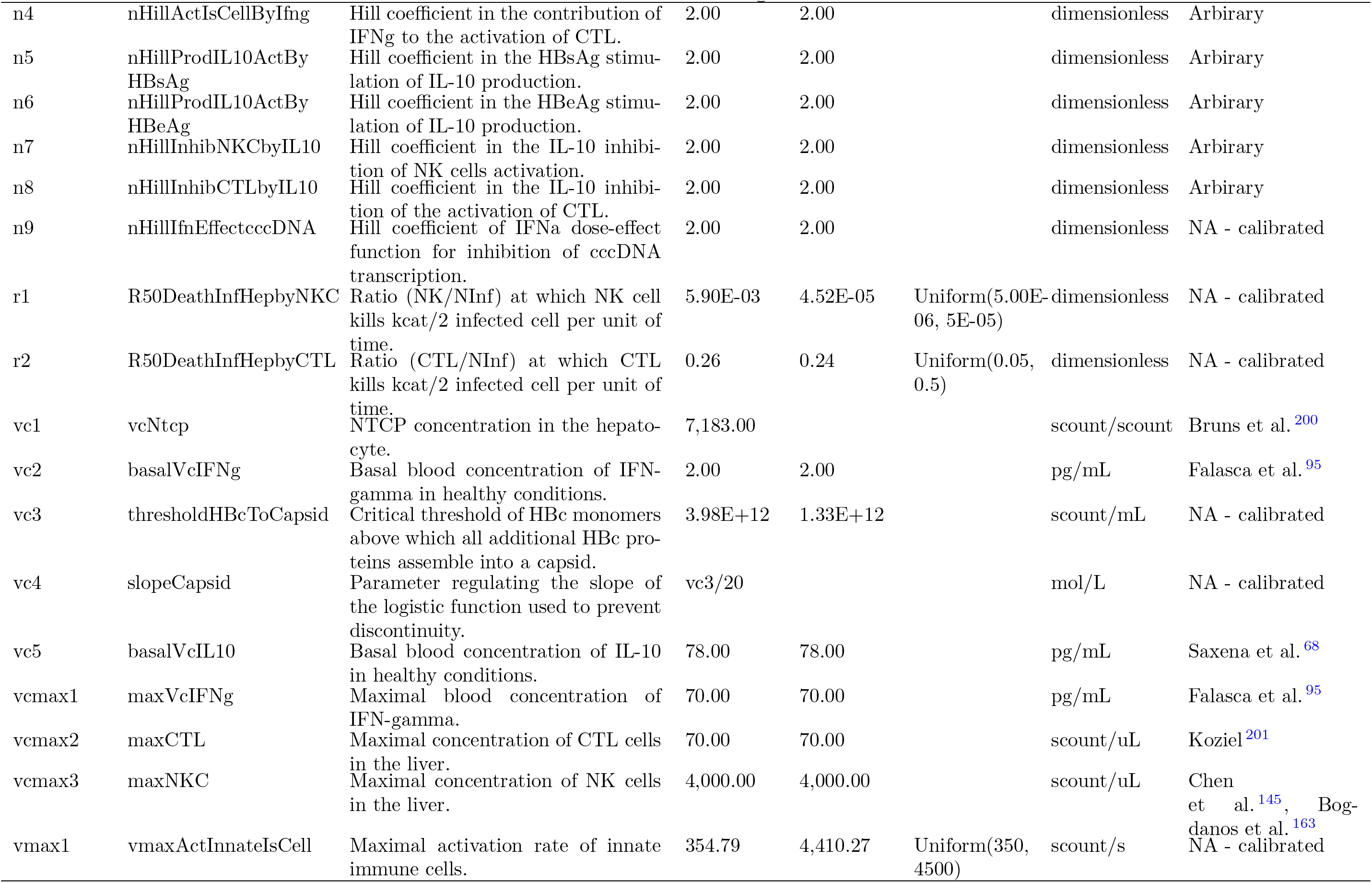
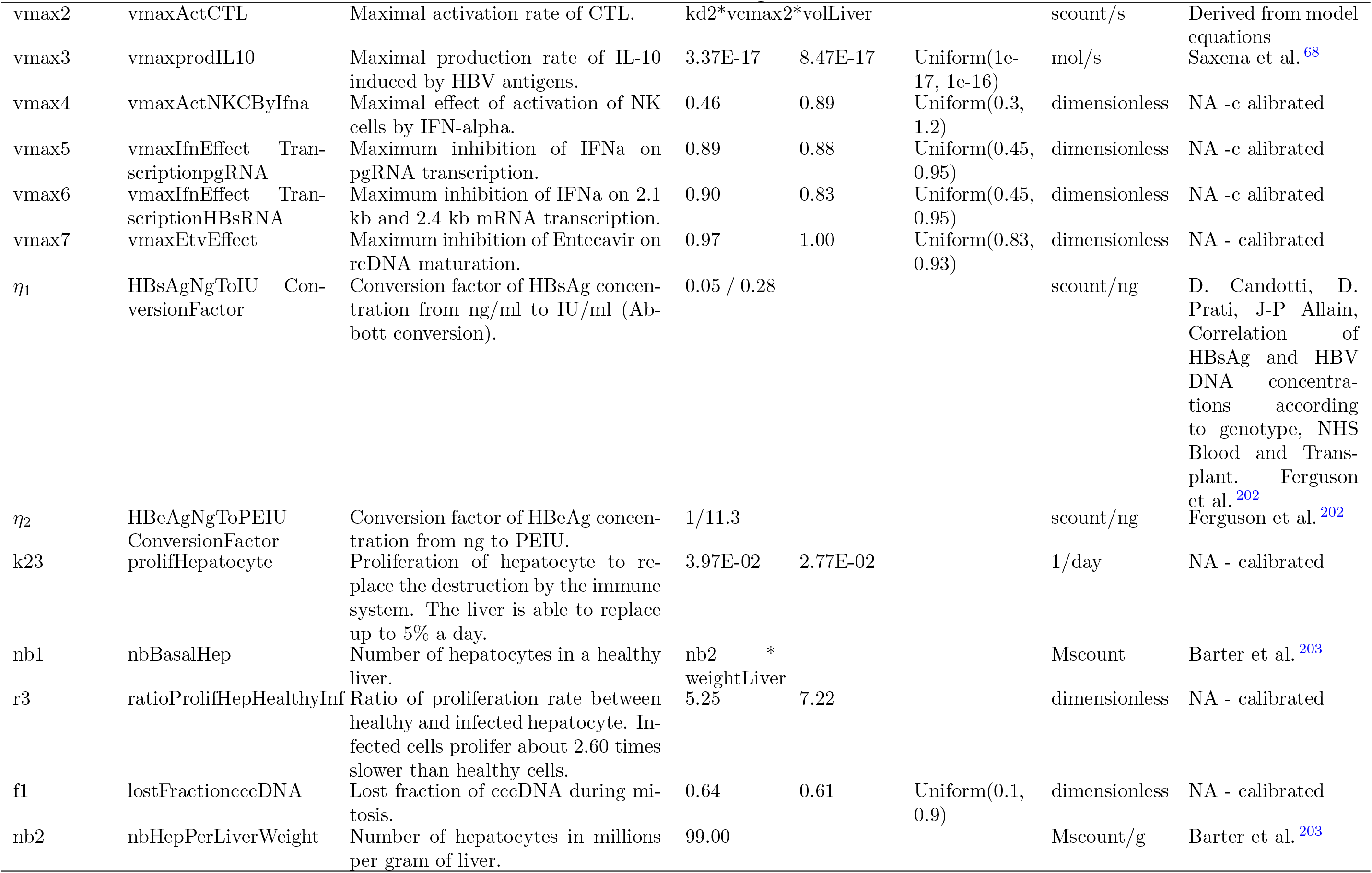
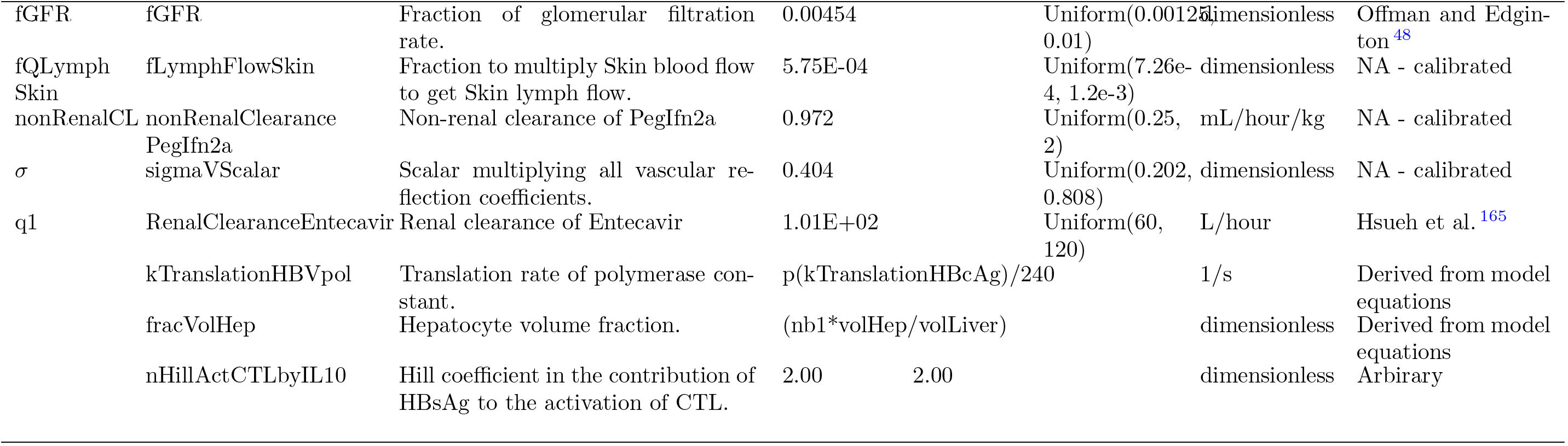
Model parameters list. Value or range column provides the final calibrated value or ranges. The reference column indicates published study used to calibrate each parameter.

**Table D.7:**
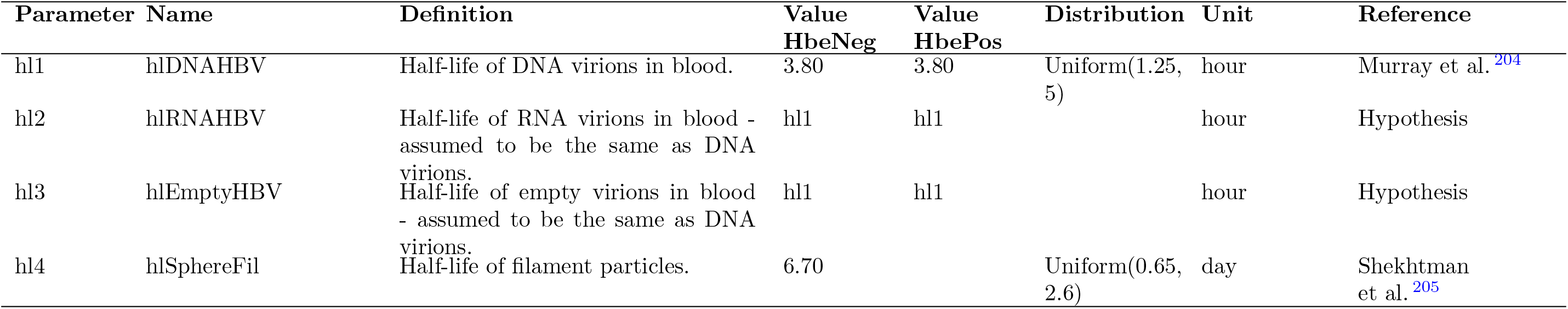
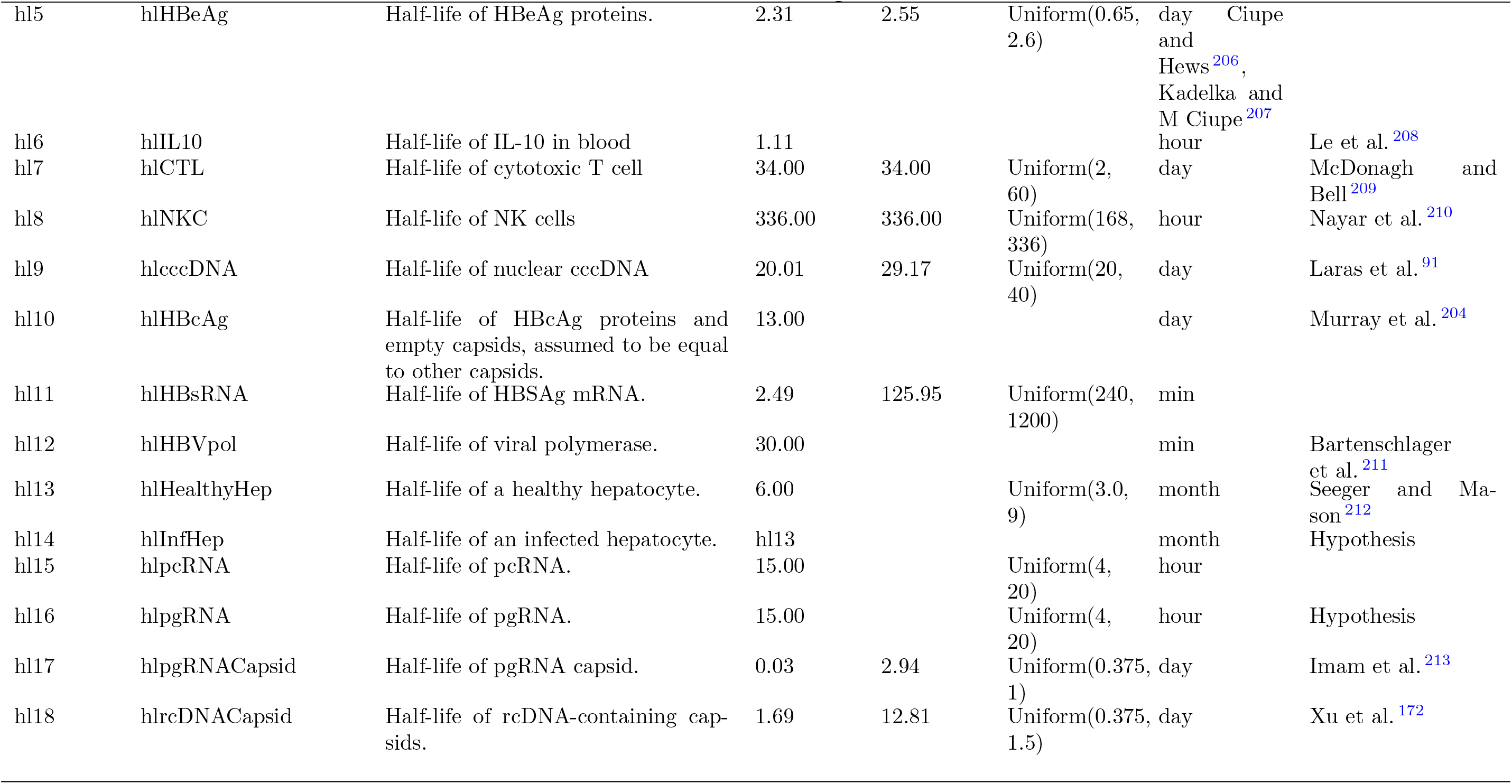
Model degradation rates list. Value or range column provides the final calibrated value or ranges. The reference column indicates published study used to calibrate each parameter. The following half-lifes account for the degradation of all the associated entities in the model. Degradation rates are given by the formula : *kdeg* = *log*(2)*/hl*

## Appendix E. Additional figures

**Figure E.6:**
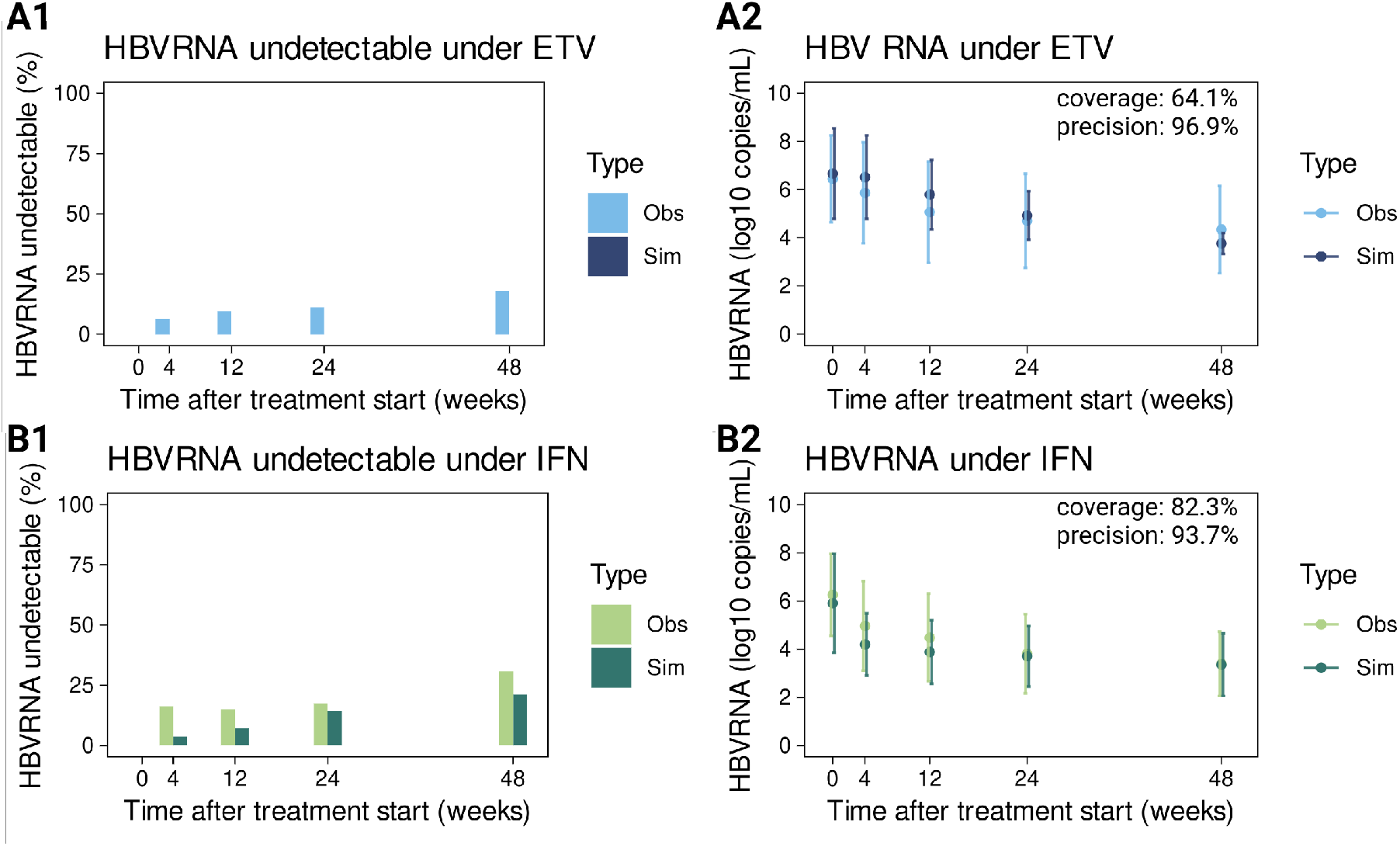
Simulated versus observed HBVRNA levels under ETV and IFN treatments. Simulations are obtained for virtual populations calibrated on Yu et al. ^9^ observations. In all plots, “Sim” (dark color) refers to simulated values obtained with calibrated virtual populations, while “Obs” (light color) refers to data extracted from Yu et al. ^9^. Data obtained under ETV treatment are in blue, while green is used for IFN treatment. Percentage of patients with undetectable levels of HBVRNA at different time points are presented under ETV (A1) and IFN (B1) treatments. Serum HBV RNA dynamics (mean ± sd) under ETV and IFN treatments are presented in A2 and B2. Coverage and precision indicators are computed as explained in the Material and Methods section.

**Figure E.7:**
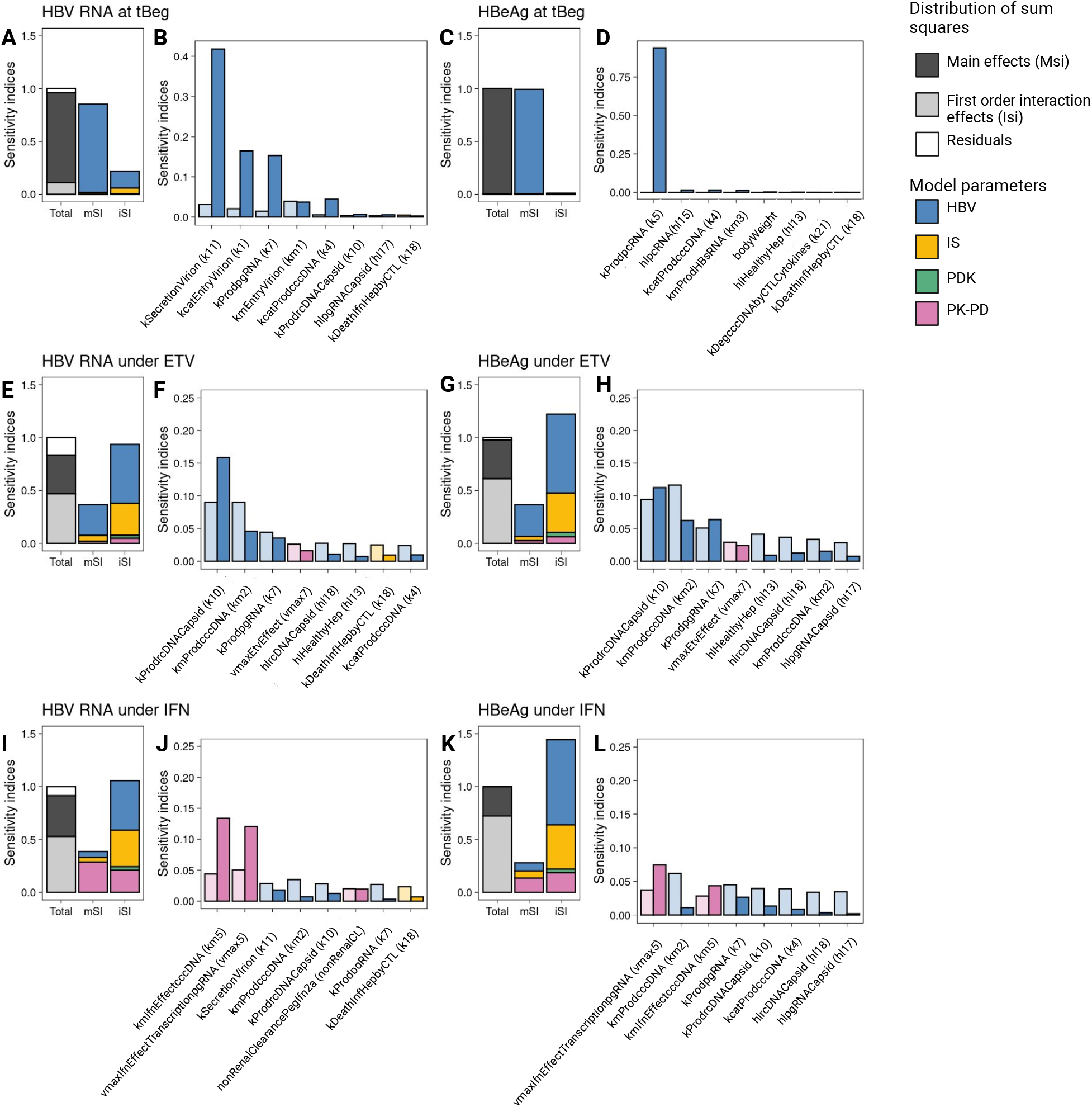
Contributions of MOCHI-B parameters in simulated SVM variability. GSA sensitivity indices for serum HBV RNA and HBeAg values at BOT and decreases between BOT (A to D) and EOT (E to L). Left plots (A, C, E, G, I and K) show the distribution of MSi and Isi and contributions of each parameter type in these indices. HBV submodel parameters are colored in blue, immune system related in yellow, drugs PK-PD in pink, and patients’ known descriptors (PDK) such as age or body weight in green. Right plots (B, D, F, H, J, L) show the effect of the 8 most important parameters for each outcome of interest.

**Figure E.8:**
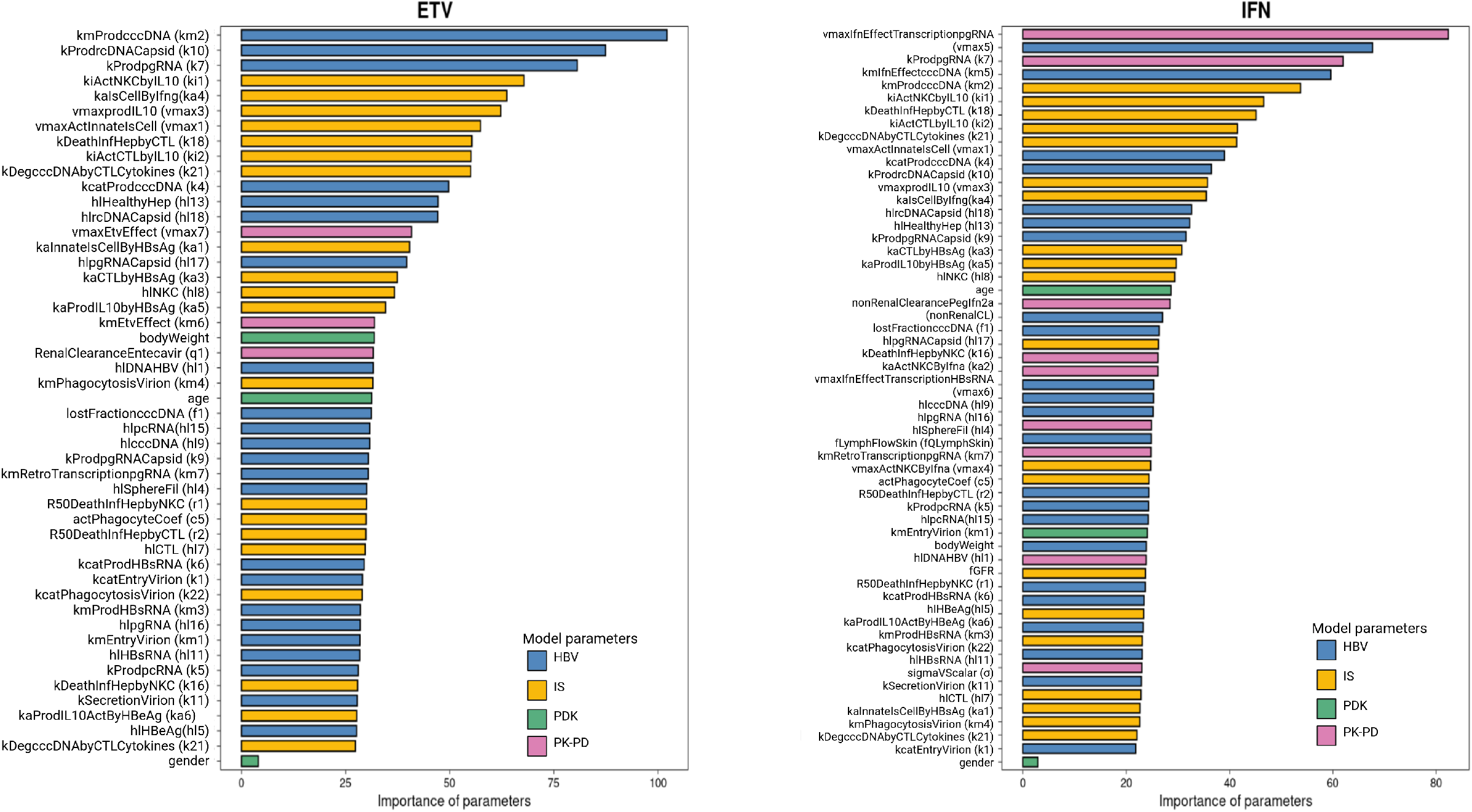
Importance of MOCHI-B parameters in explaining simulated patient response status (cure or uncured) according to random forest analysis. HBV submodel parameters are colored in blue, immune system related in yellow, drugs PK-PD in pink, and patients’ known descriptors (PDK) such as age or body weight in green. The RF model predicts VP cure classification with sensitivities of 0.93 and 0.95 and specificities of 0.83 and 0.70 for ETV and IFN respectively.

**Figure E.9:**
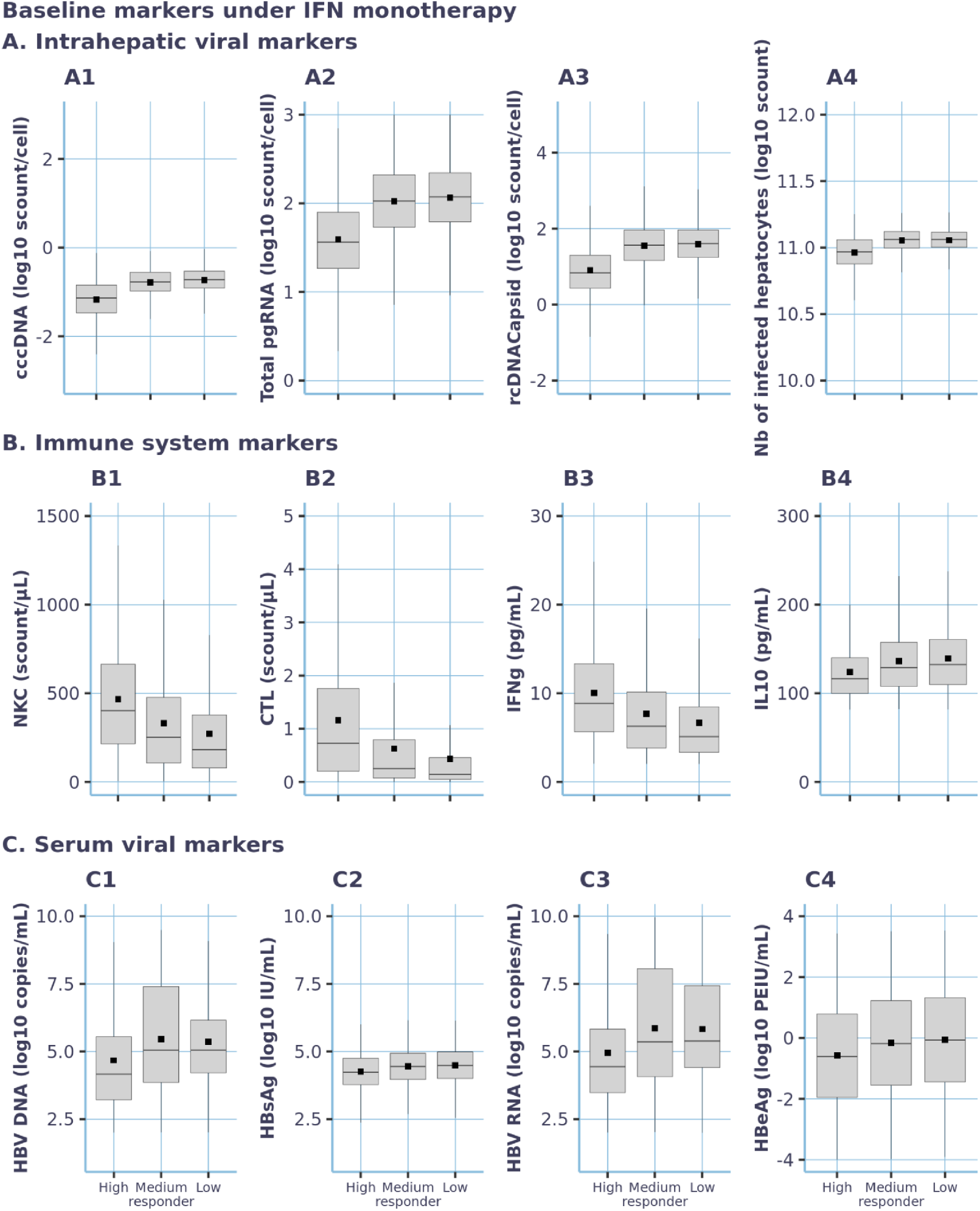
Graphical comparison of intrahepatic, immune system and serum viral markers baseline values among high, medium and low responders under IFN monotherapy treatment. The boxplots display min, 1st quartile, median, 3rd quartile and maximum values. Black squares correspond to mean values. Intrahepatic and serum viral markers are presented in log10.

## References

[1] World Health Organization, Hepatitis b, https://www.who.int/news-room/fact-sheets/detail/hepatitis-b, 2024. Consulted on September 2024.

[2] A. P. Venook, C. Papandreou, J. Furuse, L. Ladrón de Guevara, The incidence and epidemiology of hepatocellular carcinoma: a global and regional perspective, The oncologist 15 (2010) 5–13.

[3] L. A. Nicolini, A. Orsi, P. Tatarelli, C. Viscoli, G. Icardi, L. Sticchi, A global view to hbv chronic infection: evolving strategies for diagnosis, treatment and prevention in immunocompetent individuals, International Journal of Environmental Research and Public Health 16 (2019) 3307.

[4] S. Datta, S. Chatterjee, V. Veer, R. Chakravarty, Molecular biology of the hepatitis b virus for clinicians, Journal of clinical and experimental hepatology 2 (2012) 353–365.

[5] B. K. Kim, P. A. Revill, S. H. Ahn, Hbv genotypes: relevance to natural history, pathogenesis and treatment of chronic hepatitis b, Antiviral therapy 16 (2011) 1169–1186.

[6] K. L. Andersson, R. T. Chung, Monitoring during and after antiviral therapy for hepatitis b, Hepatology 49 (2009) S166–S173.

[7] T. Berg, M. Brunetto, S. Buchholz, M. Buti, C. Hly, K. Chang, M. Dandri, G. Dusheiko, J. Feld, C. Ferrari, et al., 2019 easl-aasld hbv treatment endpoints conference faculty. guidance for design and endpoints of clinical trials in chronic hepatitis b-report from the 2019 easl-aasld hbv treatment endpoints conference., JOURNAL OF HEPATOLOGY (2019).

[8] M. Sonneveld, R. Zoutendijk, H. Janssen, Hepatitis b surface antigen monitoring and management of chronic hepatitis b, Journal of viral hepatitis 18 (2011) 449– 457.

[9] X.-q. Yu, M.-j. Wang, D.-m. Yu, P.-z. Chen, M.-y. Zhu, W. Huang, Y. Han, Q.-m. Gong, X.-x. Zhang, Comparison of serum hepatitis b virus rna levels and quasispecies evolution patterns between entecavir and pegylated-interferon mono-treatment in chronic hepatitis b patients, Journal of Clinical Microbiology 58 (2020) 10–1128.

[10] C.-L. Lai, D. Shouval, A. S. Lok, T.-T. Chang, H. Cheinquer, Z. Goodman, D. DeHertogh, R. Wilber, R. C. Zink, A. Cross, et al., Entecavir versus lamivudine for patients with hbeag-negative chronic hepatitis b, New England Journal of Medicine 354 (2006) 1011– 1020.

[11] T.-T. Chang, R. G. Gish, R. De Man, A. Gadano, J. Sollano, Y.-C. Chao, A. S. Lok, K.-H. Han, Z. Goodman, J. Zhu, et al., A comparison of entecavir and lamivudine for hbeag-positive chronic hepatitis b, New England Journal of Medicine 354 (2006) 1001–1010.

[12] H.-Y. Chan, V.-S. Wong, A.-L. Chim, H.-Y. Chan, G.-H. Wong, J.-Y. Sung, Serum hbsag quantification to predict response to peginterferon therapy of e antigen positive chronic hepatitis b, Alimentary pharmacology & therapeutics 32 (2010) 1323–1331.

[13] F. Zoulim, G. Carosi, S. Greenbloom, W. Mazur, T. Nguyen, L. Jeffers, M. Brunetto, S. Yu, C. Llamoso, Quantification of hbsag in nucleos (t) ide-naive patients treated for chronic hepatitis b with entecavir with or without tenofovir in the be-low study, Journal of hepatology 62 (2015) 56–63.

[14] P. Tangkijvanich, S. Chittmittraprap, K. Poovorawan, U. Limothai, A. Khlaiphuengsin, N. Chuaypen, N. Wisedopas, Y. Poovorawan, A randomized clinical trial of peginterferon alpha-2b with or without entecavir in patients with hb eag-negative chronic hepatitis b: Role of host and viral factors associated with treatment response, Journal of viral hepatitis 23 (2016) 427–438.

[15] P. Lampertico, M. Viganó, G. G. Di Costanzo, E. Sagnelli, M. Fasano, V. Di Marco, S. Boninsegna, P. Farci, S. Fargion, T. Giuberti, et al., Randomised study comparing 48 and 96 weeks peginterferon α-2a therapy in genotype d hbeag-negative chronic hepatitis b, Gut 62 (2013) 290–298.

[16] C.-M. Tang, T. O. Yau, J. Yu, Management of chronic hepatitis b infection: current treatment guidelines, challenges, and new developments, World journal of gastroenterology: WJG 20 (2014) 6262.

[17] M. A. Fonseca, J. Z. J. Ling, O. Al-Siyabi, V. Co-Tanko, E. Chan, S. G. Lim, The efficacy of hepatitis b treatments in achieving hbsag seroclearance: a systematic review and meta-analysis, Journal of Viral Hepatitis 27 (2020) 650–662.

[18] Q. Cangelosi, S. A. Means, H. Ho, A multi-scale spatial model of hepatitis-b viral dynamics, PLoS One 12 (2017) e0188209.

[19] A. Schuch, A. Hoh, R. Thimme, The role of natural killer cells and cd8+ t cells in hepatitis b virus infection, Frontiers in immunology 5 (2014) 258.

[20] J. M. Lee, S. H. Ahn, H. S. Kim, H. Park, H. Y. Chang, D. Y. Kim, S. G. Hwang, K. S. Rim, C. Y. Chon, K.-H. Han, et al., Quantitative hepatitis b surface antigen and hepatitis b e antigen titers in prediction of treatment response to entecavir, Hepatology 53 (2011) 1486–1493.

[21] S. Liu, Y. Wu, R. Deng, R. Fan, J. Peng, W. Li, X. Liang, J. Hou, J. Sun, B. Zhou, et al., Methodology-dependent performance of serum hbv rna in predicting treatment outcomes in chronic hepatitis b patients, Antiviral Research 189 (2021) 105037.

[22] K. S. Liem, M. J. van Campenhout, Q. Xie, W. P. Brouwer, H. Chi, X. Qi, L. Chen, F. Tabak, B. E. Hansen, H. L. Janssen, Low hepatitis b surface antigen and hbv dna levels predict response to the addition of pegylated interferon to entecavir in hepatitis b e antigen positive chronic hepatitis b, Alimentary Pharmacology & Therapeutics 49 (2019) 448–456.

[23] H. Luo, X.-X. Zhang, L.-H. Cao, N. Tan, Q. Kang, H.-L. Xi, M. Yu, X.-Y. Xu, Serum hepatitis b virus rna is a predictor of hbeag seroconversion and virological response with entecavir treatment in chronic hepatitis b patients, World Journal of Gastroenterology 25 (2019) 719.

[24] P. Ren, H. Li, Y. Huang, J. Jiang, S. Guo, Z. Cao, C. Zhang, T. Zhou, Q. Gan, S. Zhao, et al., A simple-to-use tool for predicting response to peginterferon in hbv dna suppressed chronic hepatitis b patients in china, Antiviral Research 194 (2021) 105163.

[25] K. Gadkar, N. Budha, A. Baruch, J. Davis, P. Fielder, S. Ramanujan, A mechanistic systems pharmacology model for prediction of ldl cholesterol lowering by pcsk9 antagonism in human dyslipidemic populations, CPT: pharmacometrics & systems pharmacology 3 (2014) 1–9.

[26] R. Kumar, K. Thiagarajan, L. Jagannathan, L. Liu, K. Mayawala, D. de Alwis, B. Topp, Beyond the single average tumor: Understanding io combinations using a clinical qsp model that incorporates heterogeneity in patient response, CPT: pharmacometrics & systems pharmacology 10 (2021) 684–695.

[27] E. Asín-Prieto, Z. P. Parra-Guillen, J. D. G. Mantilla, J. Vandenbossche, K. Stuyckens, X. W. de Trixhe, J. J. Perez-Ruixo, I. F. Troconiz, A quantitative systems pharmacology model for acute viral hepatitis b, Computational and Structural Biotechnology Journal 19 (2021) 4997–5007.

[28] J. M. Murray, S. F. Wieland, R. H. Purcell, F. V. Chisari, Dynamics of hepatitis b virus clearance in chimpanzees, Proceedings of the National Academy of Sciences 102 (2005) 17780–17785.

[29] S. M. Ciupe, R. M. Ribeiro, P. W. Nelson, G. Dusheiko, A. S. Perelson, The role of cells refractory to productive infection in acute hepatitis b viral dynamics, Proceedings of the National Academy of Sciences 104 (2007) 5050–5055.

[30] S. Means, M. A. Ali, H. Ho, J. Heffernan, Mathematical modeling for hepatitis b virus: Would spatial effects play a role and how to model it?, Frontiers in Physiology 11 (2020) 146.

[31] J. M. Murray, A. Goyal, In silico single cell dynamics of hepatitis b virus infection and clearance, Journal of Theoretical Biology 366 (2015) 91–102.

[32] M. A. Nowak, S. Bonhoeffer, A. M. Hill, R. Boehme, H. C. Thomas, H. McDade, Viral dynamics in hepatitis b virus infection., Proceedings of the National Academy of Sciences 93 (1996) 4398–4402.

[33] P. Colombatto, L. Civitano, R. Bizzarri, F. Oliveri, S. Choudhury, R. Gieschke, F. Bonino, M. R. Brunetto, P. A. 2a HBeAg-Negative Chronic Hepatitis B Study Group, A multiphase model of the dynamics of hbv infection in hbeag-negative patients during pegylated interferon-α2a, lamivudine and combination therapy, Antiviral Therapy 11 (2006) 197–212.

[34] L. Min, W. Li, Y. Su, Y. Kuang, A mathematical model of the dynamics for anti-hbv infection treatment with peginterferon alfa-2a, in: 2008 International Conference on Communications, Circuits and Systems, IEEE, 2008, pp. 1295–1298.

[35] L. Wolters, B. Hansen, H. Niesters, D. DeHertogh, R. de Man, Viral dynamics during and after entecavir therapy in patients with chronic hepatitis bj hepatol, Hepatology (2002).

[36] H. Dahari, E. Shudo, R. M. Ribeiro, A. S. Perelson, Modeling complex decay profiles of hepatitis b virus during antiviral therapy, Hepatology 49 (2009) 32–38.

[37] A. S. Perelson, R. M. Ribeiro, Hepatitis b virus kinetics and mathematical modeling, in: Seminars in Liver Disease, volume 24 Suppl1, Published in 2004 by Thieme Medical Publishers, Inc., 333 Seventh Avenue …, 2004, pp. 11–16.

[38] S. M. Ciupe, Modeling the dynamics of hepatitis b infection, immunity, and drug therapy, Immunological Reviews 285 (2018) 38–54.

[39] M.-F. Yuen, D.-S. Chen, G. M. Dusheiko, H. L. Janssen, D. T. Lau, S. A. Locarnini, M. G. Peters, C.-L. Lai, Hepatitis b virus infection, Nature reviews Disease primers 4 (2018) 1–20.

[40] S. N. Waggoner, S. D. Reighard, I. E. Gyurova, S. A. Cranert, S. E. Mahl, E. P. Karmele, J. P. McNally, M. T. Moran, T. R. Brooks, F. Yaqoob, et al., Roles of natural killer cells in antiviral immunity, Current opinion in virology 16 (2016) 15–23.

[41] N.K. Björkström, B. Strunz, H.-G. Ljunggren, Natural killer cells in antiviral immunity, Nature Reviews Immunology 22 (2022) 112–123.

[42] F. V. Chisari, C. Ferrari, Hepatitis b virus immunopathogenesis, Annual review of immunology 13 (1995) 29–60.

[43] F. V. Chisari, et al., Cytotoxic t cells and viral hepatitis., The Journal of clinical investigation 99 (1997) 1472–1477.

[44] M. K. Maini, C. Boni, G. S. Ogg, A. S. King, S. Reignat, C. K. Lee, J. R. Larrubia, G. J. Webster, A. J. McMichael, C. Ferrari, et al., Direct ex vivo analysis of hepatitis b virus-specific cd8+ t cells associated with the control of infection, Gastroenterology 117 (1999) 1386–1396.

[45] N. Hyodo, I. Nakamura, M. Imawari, Hepatitis b core antigen stimulates interleukin-10 secretion by both t cells and monocytes from peripheral blood of patients with chronic hepatitis b virus infection, Clinical & Experimental Immunology 135 (2004) 462–466.

[46] A. Bertoletti, A. J. Gehring, The immune response during hepatitis b virus infection, Journal of General Virology 87 (2006) 1439–1449.

[47] T. Inoue, Y. Tanaka, Hepatitis b virus and its sexually transmitted infection-an update, Microbial cell 3 (2016) 420.

[48] E. Offman, A. N. Edginton, A pbpk workflow for firstin-human dose selection of a subcutaneously administered pegylated peptide, Journal of pharmacokinetics and pharmacodynamics 42 (2015) 135–150.

[49] M. R. Brunetto, F. Bonino, Interferon therapy of chronic hepatitis b, Intervirology 57 (2014) 163–170.

[50] S. R. Shuldiner, L. Gong, A. J. Muir, R. B. Altman, T. E. Klein, Pharmgkb summary: peginterferon-α pathway, Pharmacogenetics and genomics 25 (2015) 465–474.

[51] D. R. Langley, A. W. Walsh, C. J. Baldick, B. J. Eggers, R. E. Rose, S. M. Levine, A. J. Kapur, R. J. Colonno, D. J. Tenney, Inhibition of hepatitis b virus polymerase by entecavir, Journal of virology 81 (2007) 3992–4001.

[52] E. O. Voit, H. A. Martens, S. W. Omholt, 150 years of the mass action law, PLoS computational biology 11 (2015) e1004012.

[53] A. Cornish-Bowden, One hundred years of michaelis– menten kinetics, Perspectives in Science 4 (2015) 3–9.

[54] S. Marlow, Parallel and concurrent programming in Haskell: Techniques for multicore and multithreaded programming, “ O’Reilly Media, Inc.”, 2013.

[55] A. C. Hindmarsh, P. N. Brown, K. E. Grant, S. L. Lee, R. Serban, D. E. Shumaker, C. S. Woodward, Sundials: Suite of nonlinear and differential/algebraic equation solvers, ACM Transactions on Mathematical misc (TOMS) 31 (2005) 363–396.

[56] P. Städter, Y. Schälte, L. Schmiester, J. Hasenauer, P. L. Stapor, Benchmarking of numerical integration methods for ode models of biological systems, Scientific reports 11 (2021) 2696.

[57] K. Gadkar, D. Kirouac, D. Mager, P. H. van der Graaf, S. Ramanujan, A six-stage workflow for robust application of systems pharmacology, CPT: pharmacometrics & systems pharmacology 5 (2016) 235–249.

[58] W. G. Cooksley, T. Piratvisuth, S.-D. Lee, V. Mahachai, Y.-C. Chao, T. Tanwandee, A. Chutaputti, W. Y. Chang, F. Zahm, N. Pluck, Peginterferon α-2a (40 kda): an advance in the treatment of hepatitis b e antigen-positive chronic hepatitis b, Journal of viral hepatitis 10 (2003) 298–305.

[59] Q. Xie, H. Zhou, X. Bai, S. Wu, J.-J. Chen, J. Sheng, Y. Xie, C. Chen, H. L.-Y. Chan, M. Zhao, A randomized, open-label clinical study of combined pegylated interferon alfa-2a (40kd) and entecavir treatment for hepatitis b “e” antigen–positive chronic hepatitis b, Clinical infectious diseases 59 (2014) 1714–1723.

[60] G. K. Lau, T. Piratvisuth, K. X. Luo, P. Marcellin, S. Thongsawat, G. Cooksley, E. Gane, M. W. Fried, W. C. Chow, S. W. Paik, et al., Peginterferon alfa-2a, lamivudine, and the combination for hbeag-positive chronic hepatitis b, New England Journal of Medicine 352 (2005) 2682–2695.

[61] M. W. Fried, T. Piratvisuth, G. K. Lau, P. Marcellin, W.-C. Chow, G. Cooksley, K.-X. Luo, S. W. Paik, Y.-F. Liaw, P. Button, et al., Hbeag and hepatitis b virus dna as outcome predictors during therapy with peginterferon alfa-2a for hbeag-positive chronic hepatitis b, Hepatology 47 (2008) 428–434.

[62] P. Marcellin, G. K. Lau, F. Bonino, P. Farci, S. Hadziyannis, R. Jin, Z.-M. Lu, T. Piratvisuth, G. Germanidis, C. Yurdaydin, et al., Peginterferon alfa-2a alone, lamivudine alone, and the two in combination in patients with hbeag-negative chronic hepatitis b, New England Journal of Medicine 351 (2004) 1206–1217.

[63] V. Rijckborst, B. E. Hansen, Y. Cakaloglu, P. Ferenci, F. Tabak, M. Akdogan, K. Simon, U. S. Akarca, R. Flisiak, E. Verhey, et al., Early on-treatment prediction of response to peginterferon alfa-2a for hbeag-negative chronic hepatitis b using hbsag and hbv dna levels, Hepatology 52 (2010) 454–461.

[64] K. L. Gill, I. Gardner, L. Li, M. Jamei, A bottomup whole-body physiologically based pharmacokinetic model to mechanistically predict tissue distribution and the rate of subcutaneous absorption of therapeutic proteins, The AAPS journal 18 (2016) 156–170.

[65] Z. Li, D. K. Shah, Two-pore physiologically based pharmacokinetic model with de novo derived parameters for predicting plasma pk of different size protein therapeutics, Journal of pharmacokinetics and pharmacodynamics 46 (2019) 305–318.

[66] ICRP, Icrp publication 89: Basic anatomical and physiological data for use in radiological protection reference values. icrp publication 89, 2002.

[67] R. D. Huhn, E. Radwanski, J. Gallo, M. B. Affrime, R. Sabo, G. Gonyo, A. Monge, D. L. Cutler, Pharmacodynamics of subcutaneous recombinant human interleukin-10 in healthy volunteers, Clinical Pharmacology & Therapeutics 62 (1997) 171–180.

[68] R. Saxena, Y. K. Chawla, I. Verma, J. Kaur, Association of interleukin-10 with hepatitis b virus (hbv) mediated disease progression in indian population, Indian Journal of Medical Research 139 (2014) 737–745.

[69] B. Gazzard, B. Portmann, I. M. Mureay-Lyon, R. Williams, Causes of death in fulminant hepatic failure and relationship to quantitative histological assessment of parenchymal damage, QJM: An International Journal of Medicine 44 (1975) 615–626.

[70] J.-L. Palgen, A. Perrillat-Mercerot, N. Ceres, E. Peyronnet, M. Coudron, E. Tixier, B. M. Illigens, J. Bosley, A. L’Hostis, C. Monteiro, Integration of heterogeneous biological data in multiscale mechanistic model calibration: application to lung adenocarcinoma, Acta biotheoretica 70 (2022) 19.

[71] L. Wilkinson, ggplot2: elegant graphics for data analysis by wickham, h., 2011.

[72] J.-H. Yan, M. Bifano, S. Olsen, R. A. Smith, D. Zhang, D. M. Grasela, F. LaCreta, Entecavir pharmacokinetics, safety, and tolerability after multiple ascending doses in healthy subjects, The Journal of Clinical Pharmacology 46 (2006) 1250–1258.

[73] M. B. Costa, P. D. Picon, G. B. Sander, H. N. Cuni, C. V. Silva, R. P. Meireles, A. C. M. A. Goés, N. M. Batoreu, M. d. L. d. S. Maia, E. M. Albuquerque, et al., Pharmacokinetics comparison of two pegylated interferon alfa formulations in healthy volunteers, BMC Pharmacology and Toxicology 19 (2018) 1–8.

[74] R. Allen, T. R. Rieger, C. J. Musante, Efficient generation and selection of virtual populations in quantitative systems pharmacology models, CPT: pharmacometrics & systems pharmacology 5 (2016) 140–146.

[75] A. Saltelli, K. Aleksankina, W. Becker, P. Fennell, F. Ferretti, N. Holst, S. Li, Q. Wu, Why so many published sensitivity analyses are false: A systematic review of sensitivity analysis practices, Environmental modelling & misc 114 (2019) 29–39.

[76] M. Rodriguez-Fernandez, J. R. Banga, F. J. Doyle III, Novel global sensitivity analysis methodology accounting for the crucial role of the distribution of input parameters: application to systems biology models, International Journal of Robust and Nonlinear Control 22 (2012) 1082–1102.

[77] C. Friedrich, A model qualification method for mechanistic physiological qsp models to support model-informed drug development, CPT: pharmacometrics & systems pharmacology 5 (2016) 43–53.

[78] V. G. Eck, W. P. Donders, J. Sturdy, J. Feinberg, T. Delhaas, L. R. Hellevik, W. Huberts, A guide to uncertainty quantification and sensitivity analysis for cardiovascular applications, International journal for numerical methods in biomedical engineering 32 (2016) e02755.

[79] C. Bidot, H. Monod, M.-L. Taupin, A quick guide to multisensi, an r package for multivariate sensitivity analyses, 2018.

[80] R package - Cran, Multisensi, 2021. URL: https://rdrr.io/cran/multisensi/man/multisensi.html.

[81] A. Saltelli, P. Annoni, I. Azzini, F. Campolongo, M. Ratto, S. Tarantola, Variance based sensitivity analysis of model output. design and estimator for the total sensitivity index, Computer physics communications 181 (2010) 259–270.

[82] I. M. Sobol, Global sensitivity indices for nonlinear mathematical models and their monte carlo estimates, Mathematics and computers in simulation 55 (2001) 271–280.

[83] H. Monod, C. Naud, D. Makowski, Uncertainty and sensitivity analysis for crop models, Working with dynamic crop models: Evaluation, analysis, parameterization, and applications 4 (2006) 55–100.

[84] T. Homma, A. Saltelli, Importance measures in global sensitivity analysis of nonlinear models, Reliability Engineering & System Safety 52 (1996) 1–17.

[85] A. Puy, W. Becker, S. L. Piano, A. Saltelli, The battle of total-order sensitivity estimators, arXiv preprint 2009.01147 (2020).

[86] T. Zhang, J. J. Tyson, Understanding virtual patients efficiently and rigorously by combining machine learning with dynamical modelling, Journal of Pharmacokinetics and Pharmacodynamics 49 (2022) 117–131.

[87] L. Breiman, Random forests, Machine learning 45 (2001) 5–32.

[88] R. Parikh, A. Mathai, S. Parikh, G. C. Sekhar, R. Thomas, Understanding and using sensitivity, specificity and predictive values, Indian journal of ophthalmology 56 (2008) 45–50.

[89] M. Dandri, J. M. Murray, M. Lutgehetmann, T. Volz, A. W. Lohse, J. Petersen, Virion half-life in chronic hepatitis b infection is strongly correlated with levels of viremia, Hepatology 48 (2008) 1079–1086.

[90] C. R. A. Lesmana, K. Jackson, S. G. Lim, A. Sulaiman, L. S. Pakasi, R. A. Gani, I. Hasan, A. S. Sulaiman, L. A. Lesmana, R. Hammond, et al., Clinical significance of hepatitis b virion and svp productivity: relationships between intrahepatic and serum markers in chronic hepatitis b patients, United European Gastroenterology Journal 2 (2014) 99–107.

[91] A. Laras, J. Koskinas, E. Dimou, A. Kostamena, S. J. Hadziyannis, Intrahepatic levels and replicative activity of covalently closed circular hepatitis b virus dna in chronically infected patients, Hepatology 44 (2006) 694–702.

[92] T. Volz, M. Lutgehetmann, P. Wachtler, A. Jacob, A. Quaas, J. M. Murray, M. Dandri, J. Petersen, Impaired intrahepatic hepatitis b virus productivity contributes to low viremia in most hbeag-negative patients, Gastroenterology 133 (2007) 843–852.

[93] J. Wang, T. Shen, X. Huang, G. R. Kumar, X. Chen, Z. Zeng, R. Zhang, R. Chen, T. Li, T. Zhang, et al., Serum hepatitis b virus rna is encapsidated pregenome rna that may be associated with persistence of viral infection and rebound, Journal of hepatology 65 (2016) 700–710.

[94] X. Wang, X. Chi, R. Wu, H. Xu, X. Gao, L. Yu, L. Liu, M. Zhang, Y. Tan, J. Niu, et al., Serum hbv rna correlated with intrahepatic cccdna more strongly than other hbv markers during peg-interferon treatment, Virology Journal 18 (2021) 1–10.

[95] K. Falasca, C. Ucciferri, M. Dalessandro, P. Zingariello, P. Mancino, C. Petrarca, E. Pizzigallo, P. Conti, J. Vecchiet, Cytokine patterns correlate with liver damage in patients with chronic hepatitis b and c, Annals of Clinical & Laboratory Science 36 (2006) 144–150.

[96] M. K. Arababadi, A. A. Pourfathollah, A. Jafarzadeh, G. Hassanshahi, Serum levels of il-10 and il-17a in occult hbv-infected south-east iranian patients, Hepatitis monthly 10 (2010) 31.

[97] H.-K. Son, A.-Y. Lim, D.-H. Lee, S.-B. Jang, Y.-J. Lee, J.-Y. Chung, K.-S. Park, The safety and the pharmacokinetics and pharmacodynamics of a pegylated interferon alpha-2a formulation, dong-a’s da-3021, Journal of Korean Society for Clinical Pharmacology and Therapeutics 18 (2010) 117–126.

[98] M. Sunbul, Hepatitis b virus genotypes: global distribution and clinical importance, World journal of gastroenterology: WJG 20 (2014) 5427.

[99] S. Tong, P. Revill, Overview of hepatitis b viral replication and genetic variability, Journal of hepatology 64 (2016) S4–S16.

[100] M. Campos-Valdez, H.C. Monroy-Ramírez, J. Armendáriz-Borunda, L.V. Sánchez-Orozco, Molecular mechanisms during hepatitis b infection and the effects of the virus variability, Viruses 13 (2021) 1167.

[101] C. J. Baldick, B. J. Eggers, J. Fang, S. M. Levine, K. A. Pokornowski, R. E. Rose, C.-F. Yu, D. J. Tenney, R. J. Colonno, Hepatitis b virus quasispecies susceptibility to entecavir confirms the relationship between genotypic resistance and patient virologic response, Journal of hepatology 48 (2008) 895–902.

[102] D. K.-H. Wong, M. Kopaniszen, K. Omagari, Y. Tanaka, D. Y.-T. Fong, W.-K. Seto, J. Fung, F.Y. Huang, A.-y. Zhang, I. F.-N. Hung, et al., Effect of hepatitis b virus reverse transcriptase variations on entecavir treatment response, The Journal of infectious diseases 210 (2014) 701–707.

[103] M. Khatun, K. Kumar, A. Baidya, R. K. Mondal, O. Baszczynški, F. Kalčic, S. Banerjee, G. K. Dhali, K. Das, A. Chowdhury, et al., Variability in the responses of hepatitis b virus d-subgenotypes to antiviral therapy: Designing pan-d-subgenotypic reverse transcriptase inhibitors, Journal of Virology 96 (2022) e01800–21.

[104] C. Boni, P. Fisicaro, C. Valdatta, B. Amadei, P. Di Vincenzo, T. Giuberti, D. Laccabue, A. Zerbini, A. Cavalli, G. Missale, et al., Characterization of hepatitis b virus (hbv)-specific t-cell dysfunction in chronic hbv infection, Journal of virology 81 (2007) 4215–4225.

[105] S. Ezzikouri, M. E. Hoque Kayesh, S. Benjelloun, M. Kohara, K. Tsukiyama-Kohara, Targeting host innate and adaptive immunity to achieve the functional cure of chronic hepatitis b, Vaccines 8 (2020) 216.

[106] S. Yang, W. Zeng, J. Zhang, F. Lu, J. Chang, J.-T. Guo, Restoration of a functional antiviral immune response to chronic hbv infection by reducing viral antigen load: if not sufficient, is it necessary?, Emerging microbes & infections 10 (2021) 1545–1554.

[107] J. Chang, F. Guo, X. Zhao, J.-T. Guo, Therapeutic strategies for a functional cure of chronic hepatitis b virus infection, Acta pharmaceutica Sinica B 4 (2014) 248–257.

[108] H. L.-Y. Chan, V. W.-S. Wong, A. M.-L. Tse, C.-H. Tse, A. M.-L. Chim, H.-Y. Chan, G. L.-H. Wong, J. J.Y. Sung, Serum hepatitis b surface antigen quantitation can reflect hepatitis b virus in the liver and predict treatment response, Clinical Gastroenterology and Hepatology 5 (2007) 1462–1468.

[109] S. Zeuzem, E. Gane, Y.-F. Liaw, S. G. Lim, A. DiBisceglie, M. Buti, A. Chutaputti, J. Rasenack, J. Hou, C. O’Brien, et al., Baseline characteristics and early on-treatment response predict the outcomes of 2 years of telbivudine treatment of chronic hepatitis b, Journal of hepatology 51 (2009) 11–20.

[110] P. Sandhu, M. Haque, T. Humphries-Bickley, S. Ravi, J. Song, Hepatitis b virus immunopathology, model systems, and current therapies, Frontiers in immunology 8 (2017) 436.

[111] G. C. Fanning, F. Zoulim, J. Hou, A. Bertoletti, Therapeutic strategies for hepatitis b virus infection: towards a cure, Nature reviews Drug discovery 18 (2019) 827–844.

[112] F. Liu, Z. R. Liu, T. Li, Y. D. Liu, M. Zhang, Y. Xue, L. X. Zhang, Q. Ye, X. P. Fan, L. Wang, Varying 10-year off-treatment responses to nucleos (t) ide analogues in patients with chronic hepatitis b according to their pretreatment hepatitis b e antigen status, Journal of digestive diseases 19 (2018) 561–571.

[113] J. H. Na, J. H. Kim, W. H. Choe, S. Y. Kwon, B. C. Yoo, Changes in the hepatitis b surface antigen level according to the hbeag status and drug used in longterm nucleos (t) ide analog-treated chronic hepatitis b patients, The Korean Journal of Gastroenterology 77 (2021) 285–293.

[114] C. A. Biron, K. B. Nguyen, G. C. Pien, L. P. Cousens, T. P. Salazar-Mather, Natural killer cells in antiviral defense: function and regulation by innate cytokines, Annual review of immunology 17 (1999) 189–220.

[115] H.-T. Tsai, T.-H. Tsai, T.-M. Lu, C.-C. Yang, Immunopathology of hepatitis b virus infection, International Reviews of Immunology 27 (2008) 427–446.

[116] A. Das, M. K. Maini, Innate and adaptive immune responses in hepatitis b virus infection, Digestive Diseases 28 (2010) 126–132.

[117] D. Peppa, L. Micco, A. Javaid, P. T. Kennedy, Schurich, C. Dunn, C. Pallant, G. Ellis, P. Khanna, G. Dusheiko, et al., Blockade of immunosuppressive cytokines restores nk cell antiviral function in chronic hepatitis b virus infection, PLoS pathogens 6 (2010) e1001227.

[118] P. Revill, Z. Yuan, New insights into how hbv manipulates the innate immune response to establish acute and persistent infection, Antiviral therapy 18 (2013) 1–15.

[119] J. N. Stoop, R. G. van der Molen, C. C. Baan, L. J. van der Laan, E. J. Kuipers, J. G. Kusters, H. L. Janssen, Regulatory t cells contribute to the impaired immune response in patients with chronic hepatitis b virus infection, Hepatology 41 (2005) 771–778.

[120] P. Fisicaro, V. Barili, M. Rossi, I. Montali, A. Vecchi, G. Acerbi, D. Laccabue, A. Zecca, A. Penna, G. Missale, et al., Pathogenetic mechanisms of t cell dysfunction in chronic hbv infection and related therapeutic approaches, Frontiers in Immunology 11 (2020) 849.

[121] Z. Ma, E. Zhang, S. Gao, Y. Xiong, M. Lu, Toward a functional cure for hepatitis b: the rationale and challenges for therapeutic targeting of the b cell immune response, Frontiers in immunology 10 (2019) 2308.

[122] T. Pollicino, G. Caminiti, Hbv-integration studies in the clinic: role in the natural history of infection, Viruses 13 (2021) 368.

[123] C. I. Wooddell, M.-F. Yuen, H. L.-Y. Chan, R. G. Gish, S. A. Locarnini, D. Chavez, C. Ferrari, B. D. Given, J. Hamilton, S. B. Kanner, et al., Rnai-based treatment of chronically infected patients and chimpanzees reveals that integrated hepatitis b virus dna is a source of hbsag, Science translational medicine 9 (2017) eaan0241.

[124] N. Freitas, T. Lukash, S. Gunewardena, B. Chappell, L. Slagle, S. O. Gudima, Relative abundance of integrant-derived viral rnas in infected tissues harvested from chronic hepatitis b virus carriers, Journal of Virology 92 (2018) 10–1128.

[125] T. Tu, H. Zhang, S. Urban, Hepatitis b virus dna integration: in vitro models for investigating viral pathogenesis and persistence, Viruses 13 (2021) 180.

[126] S. El Messaoudi, A. Lemenuel-Diot, A. Gonçalves, J. Guedj, A semi-mechanistic model to characterize the long-term dynamic of hbv markers during treatment with lamivudine and peg-ifn., Clinical Pharmacology and Therapeutics (2022).

[127] C. Ko, T. Michler, U. Protzer, Novel viral and host targets to cure hepatitis b, Current opinion in virology 24 (2017) 38–45.

[128] S. Urban, A. Schulze, M. Dandri, J. Petersen, The replication cycle of hepatitis b virus, Journal of hepatology 52 (2010) 282–284.

[129] R. J. Lamontagne, S. Bagga, M. J. Bouchard, Hepatitis b virus molecular biology and pathogenesis, Hepatoma research 2 (2016) 163.

[130] T. J. Liang, T. M. Block, B. J. McMahon, M. G. Ghany, S. Urban, J.-T. Guo, S. Locarnini, F. Zoulim, K.-M. Chang, A. S. Lok, Present and future therapies of hepatitis b: from discovery to cure, Hepatology 62 (2015) 1893–1908.

[131] F. Zoulim, D. Durantel, Antiviral therapies and prospects for a cure of chronic hepatitis b, Cold Spring Harbor perspectives in medicine 5 (2015) a021501.

[132] E. Asin-Prieto, Z. P. Parra-Guillen, J. D. G. Mantilla, J. Vandenbossche, K. Stuyckens, X. W. de Trixhe, J. J. Perez-Ruixo, I. F. Troconiz, Immune network for viral hepatitis b: Topological representation, European Journal of Pharmaceutical Sciences 136 (2019) 104939.

[133] L. Tang, C. Chen, X. Gao, W. Zhang, X. Yan, Y. Zhou, L. Guo, X. Zheng, W. Wang, F. Yang, et al., Interleukin 21 reinvigorates the antiviral activity of hepatitis b virus (hbv)–specific cd8+ t cells in chronic hbv infection, The Journal of Infectious Diseases 219 (2019) 750–759.

[134] L. Allweiss, T. Volz, K. Giersch, J. Kah, G. Raffa, J. Petersen, A. W. Lohse, C. Beninati, T. Pollicino, S. Urban, et al., Proliferation of primary human hepatocytes and prevention of hepatitis b virus reinfection efficiently deplete nuclear cccdna in vivo, Gut 67 (2018) 542–552.

[135] M. Lutgehetmann, T. Volz, A. Köpke, T. Broja, E. Tigges, A. W. Lohse, E. Fuchs, J. M. Murray, J. Petersen, M. Dandri, In vivo proliferation of hepadnavirus-infected hepatocytes induces loss of covalently closed circular dna in mice, Hepatology 52 (2010) 16–24.

[136] T. Tu, S. Urban, Virus entry and its inhibition to prevent and treat hepatitis b and hepatitis d virus infections, Current opinion in virology 30 (2018) 68–79.

[137] T. Garcia, J. Li, C. Sureau, K. Ito, Y. Qin, J. Wands, S. Tong, Drastic reduction in the production of subviral particles does not impair hepatitis b virus virion secretion, Journal of virology 83 (2009) 11152–11165.

[138] C. Ferrari, Hbv and the immune response, Liver international 35 (2015) 121–128.

[139] A. Tan, S. Koh, A. Bertoletti, Immune response in hepatitis b virus infection, Cold Spring Harbor perspectives in medicine 5 (2015) a021428.

[140] A. Busca, A. Kumar, Innate immune responses in hepatitis b virus (hbv) infection, Virology journal 11 (2014) 1–8.

[141] S. Wieland, R. Thimme, R. H. Purcell, F. V. Chisari, Genomic analysis of the host response to hepatitis b virus infection, Proceedings of the National Academy of Sciences 101 (2004) 6669–6674.

[142] S. P. Fletcher, D. J. Chin, Y. Ji, A. L. Iniguez, B. Taillon, D. C. Swinney, P. Ravindran, D. T. Cheng, H. Bitter, U. Lopatin, et al., Transcriptomic analysis of the woodchuck model of chronic hepatitis b, Hepatology 56 (2012) 820–830.

[143] S. F. Wieland, F. V. Chisari, Stealth and cunning: hepatitis b and hepatitis c viruses, Journal of virology 79 (2005) 9369–9380.

[144] C. Dunn, D. Peppa, P. Khanna, G. Nebbia, M. Jones, N. Brendish, R. M. Lascar, D. Brown, R. J. Gilson, R. J. Tedder, et al., Temporal analysis of early immune responses in patients with acute hepatitis b virus infection, Gastroenterology 137 (2009) 1289–1300.

[145] Y. Chen, H. Wei, B. Gao, Z. Hu, S. Zheng, Z. Tian, Activation and function of hepatic nk cells in hepatitis b infection: an underinvestigated innate immune response, Journal of viral hepatitis 12 (2005) 38–45.

[146] K. Kakimi, L. G. Guidotti, Y. Koezuka, F. V. Chisari, Natural killer t cell activation inhibits hepatitis b virus replication in vivo, The Journal of experimental medicine 192 (2000) 921–930.

[147] A. Bertoletti, C. Ferrari, Kinetics of the immune response during hbv and hcv infection, Hepatology 38 (2003) 4–13.

[148] H.-J. Li, N.-C. Zhai, H.-X. Song, Y. Yang, A. Cui, T.Y. Li, Z.-K. Tu, The role of immune cells in chronic hbv infection, Journal of clinical and translational hepatology 3 (2015) 277.

[149] L. G. Guidotti, F. V. Chisari, To kill or to cure: options in host defense against viral infection, Current opinion in immunology 8 (1996) 478–483.

[150] K. Ando, L. G. Guidotti, S. Wirth, T. Ishikawa, G. Missale, T. Moriyama, R. D. Schreiber, H.-J. Schlicht, S. N. Huang, F. V. Chisari, Class i-restricted cytotoxic t lymphocytes are directly cytopathic for their target cells in vivo., Journal of immunology (Baltimore, Md.: 1950) 152 (1994) 3245–3253.

[151] L. V. Tsui, L. G. Guidotti, T. Ishikawa, F. V. Chisari, Posttranscriptional clearance of hepatitis b virus rna by cytotoxic t lymphocyte-activated hepatocytes., Proceedings of the National Academy of Sciences 92 (1995) 12398–12402.

[152] B. Rehermann, M. Nascimbeni, Immunology of hepatitis b virus and hepatitis c virus infection, Nature Reviews Immunology 5 (2005) 215–229.

[153] C.-F. Huang, S.-S. Lin, Y.-C. Ho, F.-L. Chen, C.-C. Yang, et al., The immune response induced by hepatitis b virus principal antigens, Cell Mol Immunol 3 (2006) 97–106.

[154] R. P. Beasley, Rocks along the road to the control of hbv and hcc, Annals of epidemiology 19 (2009) 231– 234.

[155] Z. Ma, E. Zhang, D. Yang, M. Lu, Contribution of toll-like receptors to the control of hepatitis b virus infection by initiating antiviral innate responses and promoting specific adaptive immune responses, Cellular & molecular immunology 12 (2015) 273–282.

[156] J. Wu, Z. Meng, M. Jiang, R. Pei, M. Trippler, R. Broering, A. Bucchi, J.-P. Sowa, U. Dittmer, D. Yang, et al., Hepatitis b virus suppresses toll-like receptor–mediated innate immune responses in murine parenchymal and nonparenchymal liver cells, Hepatology 49 (2009) 1132–1140.

[157] K. Visvanathan, N. A. Skinner, A. J. Thompson, S. M. Riordan, V. Sozzi, R. Edwards, S. Rodgers, J. Kurtovic, J. Chang, S. Lewin, et al., Regulation of tolllike receptor-2 expression in chronic hepatitis b by the precore protein, Hepatology 45 (2007) 102–110.

[158] E. Loggi, N. Gamal, F. Bihl, M. Bernardi, P. Andreone, Adaptive response in hepatitis b virus infection, Journal of viral hepatitis 21 (2014) 305–313.

[159] M. de Martino, M. E. Rossi, A. T. Muccioli, M. Resti, A. Vierucci, Interference of hepatitis b virus surface antigen with natural killer cell function., Clinical and experimental immunology 61 (1985) 90.

[160] Y. Sobao, H. Tomiyama, K. Sugi, M. Tokunaga, T. Ueno, S. Saito, S. Fujiyama, M. Morimoto, K. Tanaka, M. Takiguchi, The role of hepatitis b virus-specific memory cd8 t cells in the control of viral replication, Journal of hepatology 36 (2002) 105–115.

[161] G. J. Webster, S. Reignat, D. Brown, G. S. Ogg, L. Jones, S. L. Seneviratne, R. Williams, G. Dusheiko, A. Bertoletti, Longitudinal analysis of cd8+ t cells specific for structural and nonstructural hepatitis b virus proteins in patients with chronic hepatitis b: implications for immunotherapy, Journal of virology 78 (2004) 5707–5719.

[162] J. J. Chang, F. Wightman, A. Bartholomeusz, A. Ayres, S. J. Kent, J. Sasadeusz, S. R. Lewin, Reduced hepatitis b virus (hbv)-specific cd4+ t-cell responses in human immunodeficiency virus type 1-hbv-coinfected individuals receiving hbv-active antiretroviral therapy, Journal of virology 79 (2005) 3038–3051.

[163] D. P. Bogdanos, B. Gao, M. E. Gershwin, Liver immunology, Comprehensive Physiology 3 (2013) 567.

[164] H. M. Jones, I. B. Gardner, K. J. Watson, Modelling and pbpk simulation in drug discovery, The AAPS journal 11 (2009) 155–166.

[165] C.-H. Hsueh, V. Hsu, P. Zhao, L. Zhang, K. Giacomini, S.-M. Huang, Pbpk modeling of the effect of reduced kidney function on the pharmacokinetics of drugs excreted renally by organic anion transporters, Clinical Pharmacology & Therapeutics 103 (2018) 485–492.

[166] T. Boortalary, B. Shinn, D. Halegoua-DeMarzio, H.W. Hann, Achieving a cure: the next frontier in hepatitis b treatment, Exon Publications (2021) 109–125.

[167] E. C. Borden, G. C. Sen, G. Uze, R. H. Silverman, R. M. Ransohoff, G. R. Foster, G. R. Stark, Interferons at age 50: past, current and future impact on biomedicine, Nature reviews Drug discovery 6 (2007) 975–990.

[168] A. J. Sadler, B. R. Williams, Interferon-inducible antiviral effectors, Nature reviews immunology 8 (2008) 559–568.

[169] L. Micco, D. Peppa, E. Loggi, A. Schurich, L. Jefferson, C. Cursaro, A. M. Panno, M. Bernardi, C. Brander, F. Bihl, et al., Differential boosting of innate and adaptive antiviral responses during pegylated-interferon-alpha therapy of chronic hepatitis b, Journal of hepatology 58 (2013) 225–233.

[170] C. Boni, D. Laccabue, P. Lampertico, T. Giuberti, M. Viganó, S. Schivazappa, A. Alfieri, M. Pesci, G. B. Gaeta, G. Brancaccio, et al., Restored function of hbv-specific t cells after long-term effective therapy with nucleos (t) ide analogues, Gastroenterology 143 (2012) 963–973.

[171] S. F. Wieland, L. G. Guidotti, F. V. Chisari, Intrahepatic induction of alpha/beta interferon eliminates viral rna-containing capsids in hepatitis b virus transgenic mice, Journal of virology 74 (2000) 4165–4173.

[172] C. Xu, H. Guo, X.-B. Pan, R. Mao, W. Yu, X. Xu, L. Wei, J. Chang, T. M. Block, J.-T. Guo, Interferons accelerate decay of replication-competent nucleocapsids of hepatitis b virus, Journal of virology 84 (2010) 9332–9340.

[173] J. Li, S. Lin, Q. Chen, L. Peng, J. Zhai, Y. Liu, Z. Yuan, Inhibition of hepatitis b virus replication by myd88 involves accelerated degradation of pregenomic rna and nuclear retention of pre-s/s rnas, Journal of virology 84 (2010) 6387–6399.

[174] L. Belloni, L. Allweiss, F. Guerrieri, N. Pediconi, T. Volz, T. Pollicino, J. Petersen, G. Raimondo, M. Dandri, M. Levrero, et al., Ifn-α inhibits hbv transcription and replication in cell culture and in humanized mice by targeting the epigenetic regulation of the nuclear cccdna minichromosome, The Journal of clinical investigation 122 (2012) 529–537.

[175] N. Chuaypen, M. Sriprapun, K. Praianantathavorn, S. Payungporn, N. Wisedopas, Y. Poovorawan, P. Tangkijvanich, Kinetics of serum hbsag and intrahepatic cccdna during pegylated interferon therapy in patients with hbeag-positive and hbeag-negative chronic hepatitis b, Journal of medical virology 89 (2017) 130–138.

[176] D. Mu, F.-C. Yuan, Y. Chen, X.-Y. Jiang, L. Yan, L.Y. Jiang, J.-P. Gong, D.-Z. Zhang, H. Ren, Y. Liao, Baseline value of intrahepatic hbv dna over cccdna predicts patient’s response to interferon therapy, Scientific Reports 7 (2017) 5937.

[177] C. Niu, L. Li, S. Daffis, J. Lucifora, M. Bonnin, S. Maadadi, E. Salas, R. Chu, H. Ramos, C. M. Livingston, et al., Toll-like receptor 7 agonist gs9620 induces prolonged inhibition of hbv via a type i interferon-dependent mechanism, Journal of hepatology 68 (2018) 922–931.

[178] Y. Shen, N. L. Li, J. Wang, B. Liu, S. Lester, K. Li, Trim56 is an essential component of the tlr3 antiviral signaling pathway, Journal of Biological Chemistry 287 (2012) 36404–36413.

[179] P. Mutz, P. Metz, F. A. Lempp, S. Bender, B. Qu, K. Schöneweis, S. Seitz, T. Tu, A. Restuccia, J. Frankish, et al., Hbv bypasses the innate immune response and does not protect hcv from antiviral activity of interferon, Gastroenterology 154 (2018) 1791–1804.

[180] Rohatgi A., Webplotdigitizer, 2020. URL: https://automeris.io/WebPlotDigitizer.

[181] D. Ganem, A. M. Prince, Hepatitis b virus infection—natural history and clinical consequences, New England Journal of Medicine 350 (2004) 1118–1129.

[182] C. Trépo, H. L. Chan, A. Lok, Hepatitis b virus infection, The Lancet 384 (2014) 2053–2063.

[183] L. Luckenbaugh, K. Kitrinos, W. Delaney Iv, J. Hu, Genome-free hepatitis b virion levels in patient sera as a potential marker to monitor response to antiviral therapy, Journal of viral hepatitis 22 (2015) 561–570.

[184] Y. Gao, Y. Li, Q. Meng, Z. Zhang, P. Zhao, Q. Shang, Y. Li, M. Su, T. Li, X. Liu, et al., Serum hepatitis b virus dna, rna, and hbsag: which correlated better with intrahepatic covalently closed circular dna before and after nucleos (t) ide analogue treatment?, Journal of clinical microbiology 55 (2017) 2972–2982.

[185] M. Wang, N. Qiu, S. Lu, D. Xiu, J. Yu, X. T. Wang, F. Lu, T. Li, X. Liu, H. Zhuang, Serum hepatitis b surface antigen is correlated with intrahepatic total hbv dna and cccdna in treatment-naïve patients with chronic hepatitis b but not in patients with hbv related hepatocellular carcinoma, Journal of medical virology 85 (2013) 219–227.

[186] Q. Chen, H. Chen, W. Wang, J. Liu, W. Liu, P. Ni, G. Sang, G. Wang, F. Zhou, J. Zhang, Glycyrrhetic acid, but not glycyrrhizic acid, strengthened entecavir activity by promoting its subcellular distribution in the liver via efflux inhibition, European Journal of Pharmaceutical Sciences 106 (2017) 313–327.

[187] R package Cran, Algdesign, 2022. URL: https://cran.r-project.org/web/packages/AlgDesign/index.html.

[188] N. Chai, H. E. Chang, E. Nicolas, Z. Han, M. Jarnik, J. Taylor, Properties of subviral particles of hepatitis b virus, Journal of virology 82 (2008) 7812–7817.

[189] C. Seeger, W. S. Mason, Molecular biology of hepatitis b virus infection, Virology 479 (2015) 672–686.

[190] U. Mahlknecht, S. Kaiser, Age-related changes in peripheral blood counts in humans, ExpErimEntal and thErapEutic mEdicinE 1 (2010) 1019–1025.

[191] S. Halle, K. A. Keyser, F. R. Stahl, A. Busche, A. Marquardt, X. Zheng, M. Galla, V. Heissmeyer, K. Heller, J. Boelter, et al., In vivo killing capacity of cytotoxic t cells is limited and involves dynamic interactions and t cell cooperativity, Immunity 44 (2016) 233–245.

[192] W. D. Wick, O. O. Yang, L. Corey, S. G. Self, How many human immunodeficiency virus type 1-infected target cells can a cytotoxic t-lymphocyte kill?, Journal of virology 79 (2005) 13579–13586.

[193] M. Djaldetti, H. Bessler, High temperature affects the phagocytic activity of human peripheral blood mononuclear cells, Scandinavian Journal of Clinical and Laboratory Investigation 75 (2015) 482–486.

[194] A. M. Nikiforuk, M. E. Karim, D. M. Patrick, A. N. Jassem, Influence of chronic hepatitis c infection on the monocyte-to-platelet ratio: data analysis from the national health and nutrition examination survey (2009–2016), BMC Public Health 21 (2021) 1–11.

[195] DrugBank, Entecavir, 2021. URL: https://pubchem.ncbi.nlm.nih.gov/compound/Entecavir.

[196] Prospecbio, M-hbsag, 2021. URL: https://www.prospecbio.com/hbsag_pre_s2.

[197] FDA, Peginterferon alfa 2a, 2008. URL: https://www.accessdata.fda.gov/drugsatfda_docs/label/2008/103964s5154lbl.pdf.

[198] F. Messageot, S. Salhi, P. Eon, J.-M. Rossignol, Proteolytic processing of the hepatitis b virus e antigen precursor: cleavage at two furin consensus sequences, Journal of Biological Chemistry 278 (2003) 891–895.

[199] PubMed, S-hbsag, 2021. URL: https://go.drugbank.com/polypeptides/Q69600.

[200] M. Bruns, S. Miska, S. Chassot, H. Will, Enhancement of hepatitis b virus infection by noninfectious subviral particles, Journal of virology 72 (1998) 1462–1468.

[201] M. J. Koziel, Cytokines in viral hepatitis, Seminars in liver disease 19 (1999) 157–169.

[202] M. Ferguson, N. Lelie, M. Nubling, S. Nick, W. Gerlich, R. Decker, A. Padilla, B. Unit, W. H. Organization, W. E. C. on Biological Standardization, et al., WHO Working Group on Hepatitis and HIV Diagnostic Kits: report of a collaborative study to 1) assess the suitability of candidate replacement international standard for HBsAG and a reference panel for HBsAG and 2) to calibrate the candidate standard in IU, Technical Report, World Health Organization, 2003.

[203] Z. E. Barter, M. K. Bayliss, P. H. Beaune, A. R. Boobis, D. J. Carlile, R. J. Edwards, J. Brian Houston, B. G. Lake, J. C. Lipscomb, O. R. Pelkonen, et al., Scaling factors for the extrapolation of in vivo metabolic drug clearance from in vitro data: reaching a consensus on values of human micro-somal protein and hepatocellularity per gram of liver, Current drug metabolism 8 (2007) 33–45.

[204] J. M. Murray, R. H. Purcell, S. F. Wieland, The halflife of hepatitis b virions, Hepatology 44 (2006) 1117– 1121.

[205] L. Shekhtman, S. J. Cotler, L. Hershkovich, S. L. Uprichard, M. Bazinet, V. Pantea, V. Cebotarescu, L. Cojuhari, P. Jimbei, A. Krawczyk, et al., Modelling hepatitis d virus rna and hbsag dynamics during nucleic acid polymer monotherapy suggest rapid turnover of hbsag, Scientific Reports 10 (2020) 7837.

[206] S. M. Ciupe, S. Hews, Mathematical models of eantigen mediated immune tolerance and activation following prenatal hbv infection, PLoS One 7 (2012) e39591.

[207] S. Kadelka, S. M Ciupe, Mathematical investigation of hbeag seroclearance, Mathematical Biosciences and Engineering 16 (2019).

[208] T. Le, L. Leung, W. L. Carroll, K. R. Schibler, Regulation of interleukin-10 gene expression: possible mechanisms accounting for its upregulation and for maturational differences in its expression by blood mononuclear cells, Blood, The Journal of the American Society of Hematology 89 (1997) 4112–4119.

[209] M. McDonagh, E. Bell, The survival and turnover of mature and immature cd8 t cells., Immunology 84 (1995) 514.

[210] S. Nayar, P. Dasgupta, C. Galustian, Extending the lifespan and efficacies of immune cells used in adoptive transfer for cancer immunotherapies–a review, Oncoimmunology 4 (2015) e1002720.

[211] R. Bartenschlager, M. Junker-Niepmann, H. Schaller, The p gene product of hepatitis b virus is required as a structural component for genomic rna encapsidation, Journal of virology 64 (1990) 5324–5332.

[212] C. Seeger, W. S. Mason, Hepatitis b virus biology, Microbiology and molecular biology reviews 64 (2000) 51–68.

[213] H. Imam, M. Khan, N. S. Gokhale, A. B. McIntyre, G.-W. Kim, J. Y. Jang, S.-J. Kim, C. E. Mason, S. M. Horner, A. Siddiqui, N6-methyladenosine modification of hepatitis b virus rna differentially regulates the viral life cycle, Proceedings of the National Academy of Sciences 115 (2018) 8829–8834.

